## Supplementary Information for "The relative contribution of close-proximity contacts, shared classroom exposure and indoor air quality to respiratory virus transmission in schools"

### A Genomic analysis

#### A.1 Supplementary methods

Viral genomes from specimens testing positive for Influenza A (IAV), Influenza B (IBV), or respiratory syncytial virus (RSV) were sequenced using a tiled PCR amplicon-based Illumina™ sequencing-by-synthesis (SBS) workflow. Complementary DNA (cDNA) was generated from viral RNA and subsequently amplified using the xGen™ Respiratory Virus Amplicon Panel (IDT), which employs targeted primers for comprehensive genome coverage. The resulting amplicon libraries were prepared following standard Illumina library preparation protocols and sequenced on an Illumina NovaSeq X system. Resulting reads were aligned using BLAST to reference genomes for RSV A, RSV B, and IAV H1N1 and H3N2 and IBV victoria; accession numbers are detailed below. Consensus sequences were established for all specimens that had at least 20% coverage with depth of one read. For each specimen we generated three consensus sequences, masking sites with depth lower than 5, 10 and 20 reads.

We analysed the consensus sequences to establish likely transmission pairs and to evaluate the likely number of imported cases to best explain the diversity of IAV/IBV and RSV virus sampled from the school population. After sequencing it was clear that the general coverage of the consensus sequences was generally poor, but markedly better in the NA region of the IAV genome and the G region of the RSV genome Figure S1. For this reason, we used these sections exclusively for our analysis.

To establish likely transmission events, we compared every pair of sequences by quantifying the number of single nucleotide polymorphisms (SNPs) between sequences as a proportion of comparable sites (sites covered by the consensus in both sequences). Further, we calculated the likelihood that the viral RNA sequenced from each pair of hosts was consistent with a transmission event between them using Jukes-Cantor 1969 (JC69) model [1], assuming an evolution time of the number of days between test dates plus a perceived maximum of 4 days (2 days of within-host evolution per sample), and using the closed form [2]

$$\log \mathcal{L}(\mu, t \mid s_i, s_j) = l(s_i, s_j) \log\left(\frac{1}{4} + \frac{3}{4}e^{-\frac{4}{3}\mu t}\right) + d(s_i, s_j) \log\left(\frac{1}{4} - \frac{1}{4}e^{-\frac{4}{3}\mu t}\right),$$

where $\mu$ is the substitution rate, $t$ is the time between samples in evolutionary time. For both, we assumed a nucleotide substitution rate of $1.5 \times 10^{-3}$ mutations per site per year, which is close to the estimates in literature [3,4].

Further, to evaluate the relationship between sequences in a more holistic sense, we constructed maximum likelihood phylogenetic trees using a coalescent model using the *Nextstrain* framework [5]. Alongside the sequences acquired through the school study, we also included sequences from clinical samples across Switzerland [6] and publicly available sequences from GenBank (see the list of accession codes below).

#### A.2 Supplementary results

Consensus sequences were successfully defined for 10/13 samples testing positive for IAV, 3/6 for IBV (and 8/11 for RSV. The coverage of the consensus sequences was variable with 16–93% for IAV, 7–96% for IV and 10–86% for RSV at a read depth threshold of 10 (Figure S1). Due to the low number and relatively poor quality of IBV sequences, we chose not to analyse them in detail. Because IAV has a segmented genome, recombination of the segments makes comparison for the whole genomes challenging. Typically, the HA and NA genes are used for phylogenetic comparisons. Since in our case the HA gene was poorly covered, we focussed on the NA gene. For RSV, we focused on the G gene, which had consistently high coverage (site 4637 to 5631). Comparing the pairs of IAV consensus sequences, we found between 0 and 25 substitutions corresponding up to 0.025 substitutions per comparable pair of bases; the generation-based and JC69 log-likelihood values agreed well and resulted in 15 possible transmission pairs. For RSV, between 0 and 233 substitutions were present between sequences corresponding to up to 0.42 substitutions per comparable site. The JC69 and generation-based likelihoods were sufficiently comparable to identify the same 9 possible transition pairs. Pairwise comparisons for IAV and RSV sequences at read depths 5, 10, and 20 are shown in Figures S2–S7.

The IAV NA sequences were clustered on the maximum likelihood phylogenetic tree (Figure S8), suggesting that the viruses are all relatively closely related compared to the global diversity. The cluster was in a part of the tree amongst many other sequences from Switzerland; however, the location of this cluster is likely to be highly uncertain due to the relative genetic divergence from the rest of the tree. Moreover, the genetic divergences within the sub-clade are relatively large and mostly inconsistent with direct transmission between hosts. This structure partly arises because although five sequences are identical in overlapping sections (S-7, K-32, K-39, K-95 and K-75), they diverge on the tree as a subset (K-32 and K-95) are identical to S-7, where the others diverge. This introduces uncertainty into the precise local structure of the tree. The RSV G gene tree reveals four samples clustered together are genetically identical when overlapping sections are compared on a pairwise basis, and two that are distantly related. Of the four that are closely related, three (K-10, K-38 and P-67) are clustered together on the tree, with no divergence separating them. Sample P-79 has been placed in a separate clade on the tree despite being identical in the pairwise comparison, due to its relative proximity to sample K-75. The additional insights provided by the tree serve to highlight some dependency when assigning transmission pairs, however, precise groupings of samples is not possible with the information available.

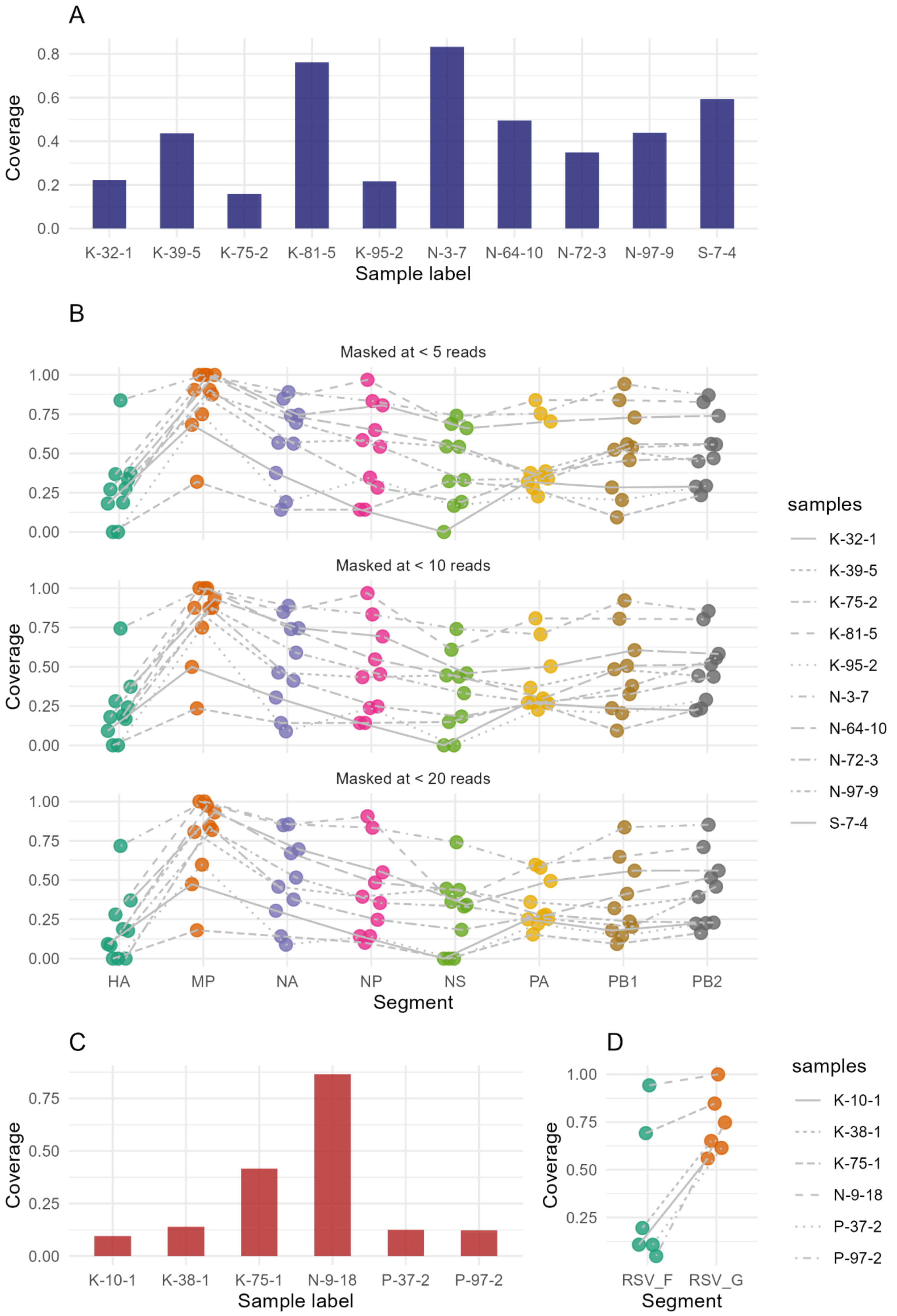

A
Coverage
0.0
0.2
0.4
0.6
0.8
K-32-1
K-39-5
K-75-2
K-81-5
K-95-2
N-3-7
N-64-10
N-72-3
N-97-9
S-7-4
Sample label
B
Masked at < 5 reads
Masked at < 10 reads
Masked at < 20 reads
Coverage
0.00
0.25
0.50
0.75
1.00
HA
MP
NA
NP
NS
PA
PB1
PB2
Segment
samples
K-32-1
K-39-5
K-75-2
K-81-5
K-95-2
N-3-7
N-64-10
N-72-3
N-97-9
S-7-4
C
Coverage
0.00
0.25
0.50
0.75
K-10-1
K-38-1
K-75-1
N-9-18
P-37-2
P-97-2
Sample label
D
Coverage
0.25
0.50
0.75
1.00
RSV_F
RSV_G
Segment
samples
K-10-1
K-38-1
K-75-1
N-9-18
P-37-2
P-97-2

**Figure S1**: Coverage of the consensus sequences. (A) Coverage of the whole influenza A H1N1 genome for each sample masked at read depth of 10. (B) Coverage for each segment of the influenza A H1N1 masked at different read depth thresholds (5, 10 and 20 reads). (C) Coverage of the whole genome of respiratory syncytial virus A for each sample masked at read depth of 10. D) Coverage for the F and G genes of RSV A masked a read depth threshold of 10.

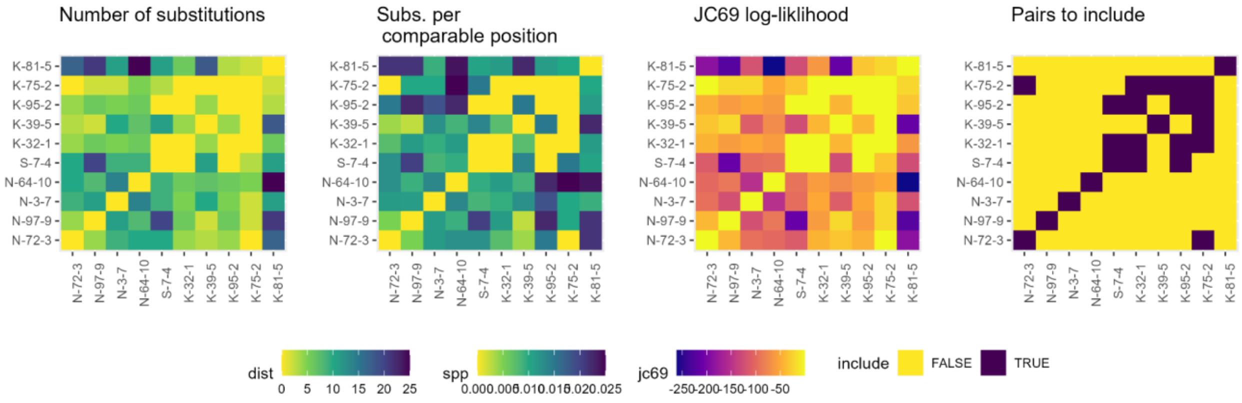

**Figure S2:** Pairwise comparison of the consensus sequences for the NA gene of influenza A virus H1N1, with masking at read depth of 5. The panels show the number of single nucleotide polymorphisms (SNPs), SNPs per nucleotide shared between sequences, the likelihood that pairs are consistent with a transmission event based on the Jukes-Cantor 1969 model and the pairs that were selected to be included or removed from the pairwise analysis.

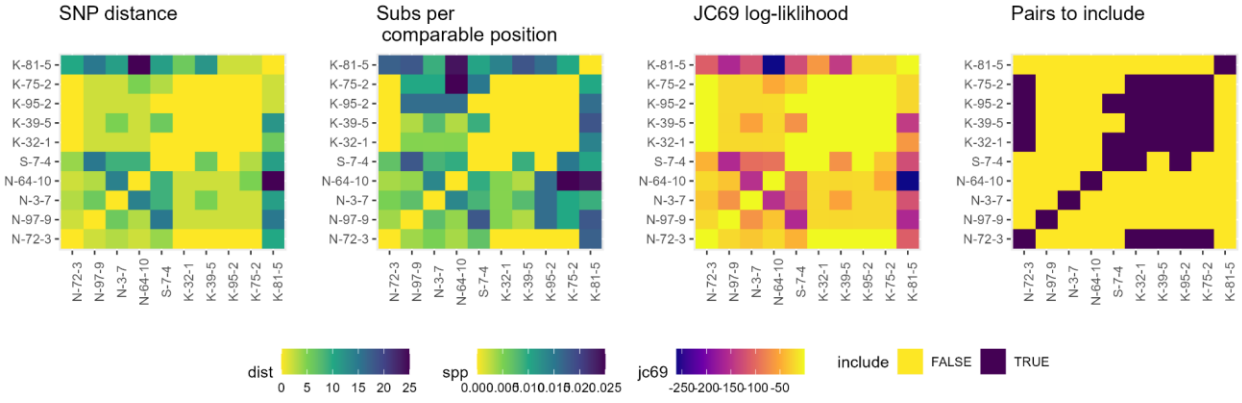

**Figure S3:** Pairwise comparison of the consensus sequences for the NA gene of influenza A virus H1N1, with masking at read depth of 10. The panels show the number of single nucleotide polymorphisms (SNPs), SNPs per nucleotide shared between sequences, the likelihood that pairs are consistent with a transmission event based on the Jukes-Cantor 1969 model and the pairs that were selected to be included or removed from the pairwise analysis.

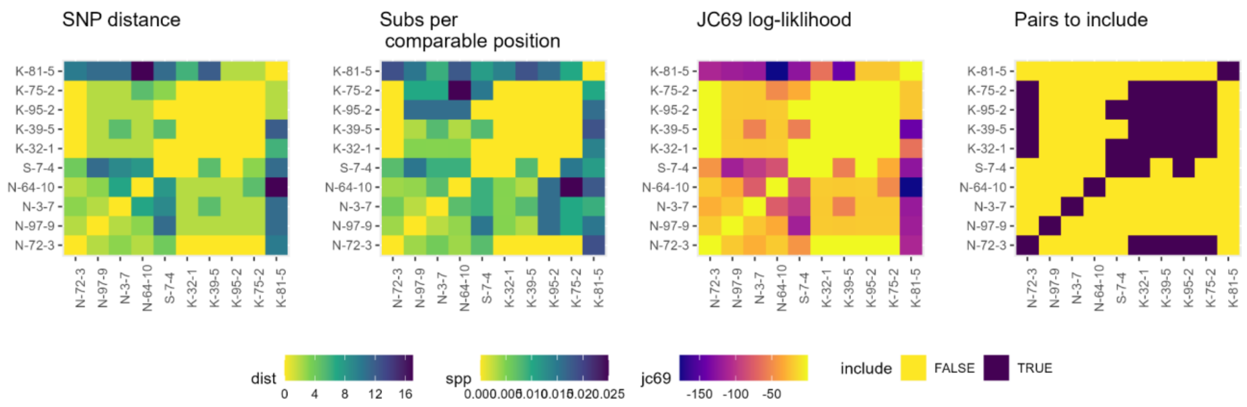

**Figure S4:** Pairwise comparison of the consensus sequences for the NA gene of influenza A virus H1N1, with masking at read depth of 20. The panels show the number of single nucleotide polymorphisms (SNPs), SNPs per nucleotide shared between sequences, the likelihood that pairs are consistent with a transmission event based on the Jukes-Cantor 1969 model and the pairs that were selected to be included or removed from the pairwise analysis.

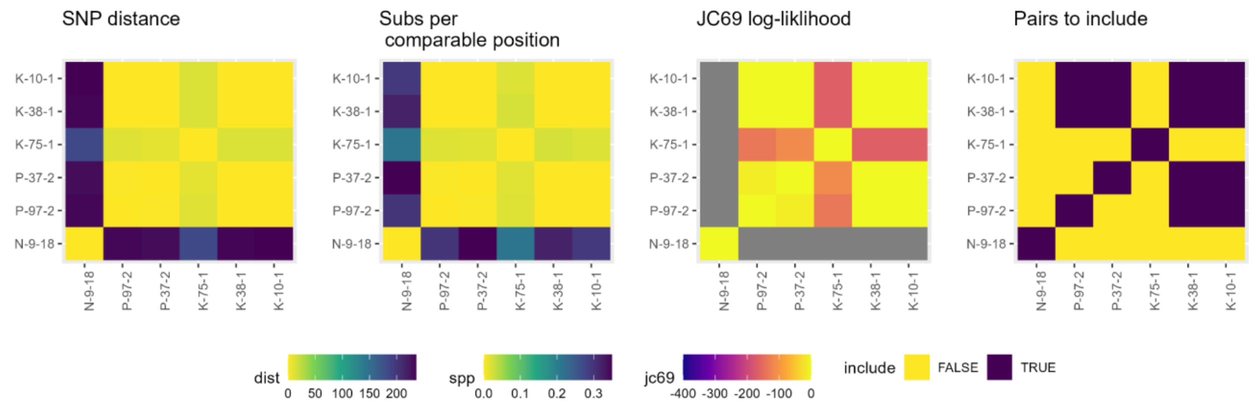

**Figure S5:** Pairwise comparison of the consensus sequences for the G gene of gene respiratory syncytial virus, with masking at read depth of 5. The panels show the number of single nucleotide polymorphisms (SNPs), SNPs per nucleotide shared between sequences, the likelihood that pairs are consistent with a transmission event based on the Jukes-Cantor 1969 model and the pairs that were selected to be included or removed from the pairwise analysis.

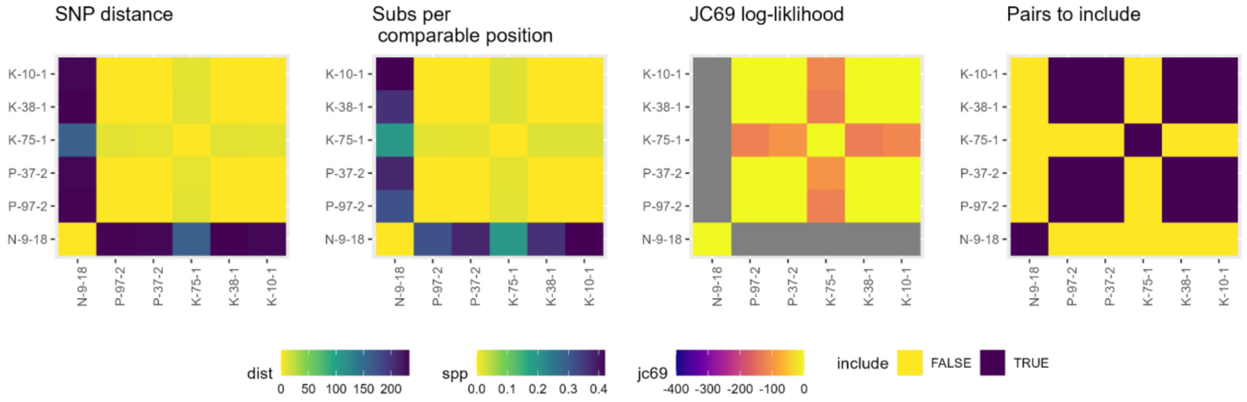

**Figure S6:** Pairwise comparison of the consensus sequences for the G gene of respiratory syncytial virus, with masking at read depth of 10. The panels show the number of single nucleotide polymorphisms (SNPs), SNPs per nucleotide shared between sequences, the likelihood that pairs are consistent with a transmission event based on the Jukes-Cantor 1969 model and the pairs that were selected to be included or removed from the pairwise analysis.

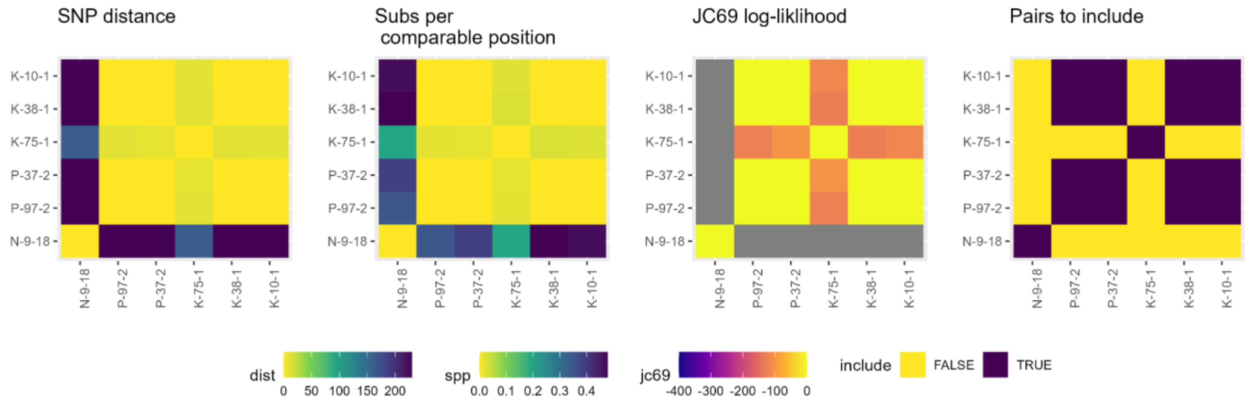

**Figure S7:** Pairwise comparison of the consensus sequences for the G of gene respiratory syncytial virus, with masking at read depth of 20. The panels show the number of single nucleotide polymorphisms (SNPs), SNPs per nucleotide shared between sequences, the likelihood that pairs are consistent with a transmission event based on the Jukes-Cantor 1969 model and the pairs that were selected to be included or removed from the pairwise analysis.

### B Pairwise survival analysis

#### B.1 Supplementary data

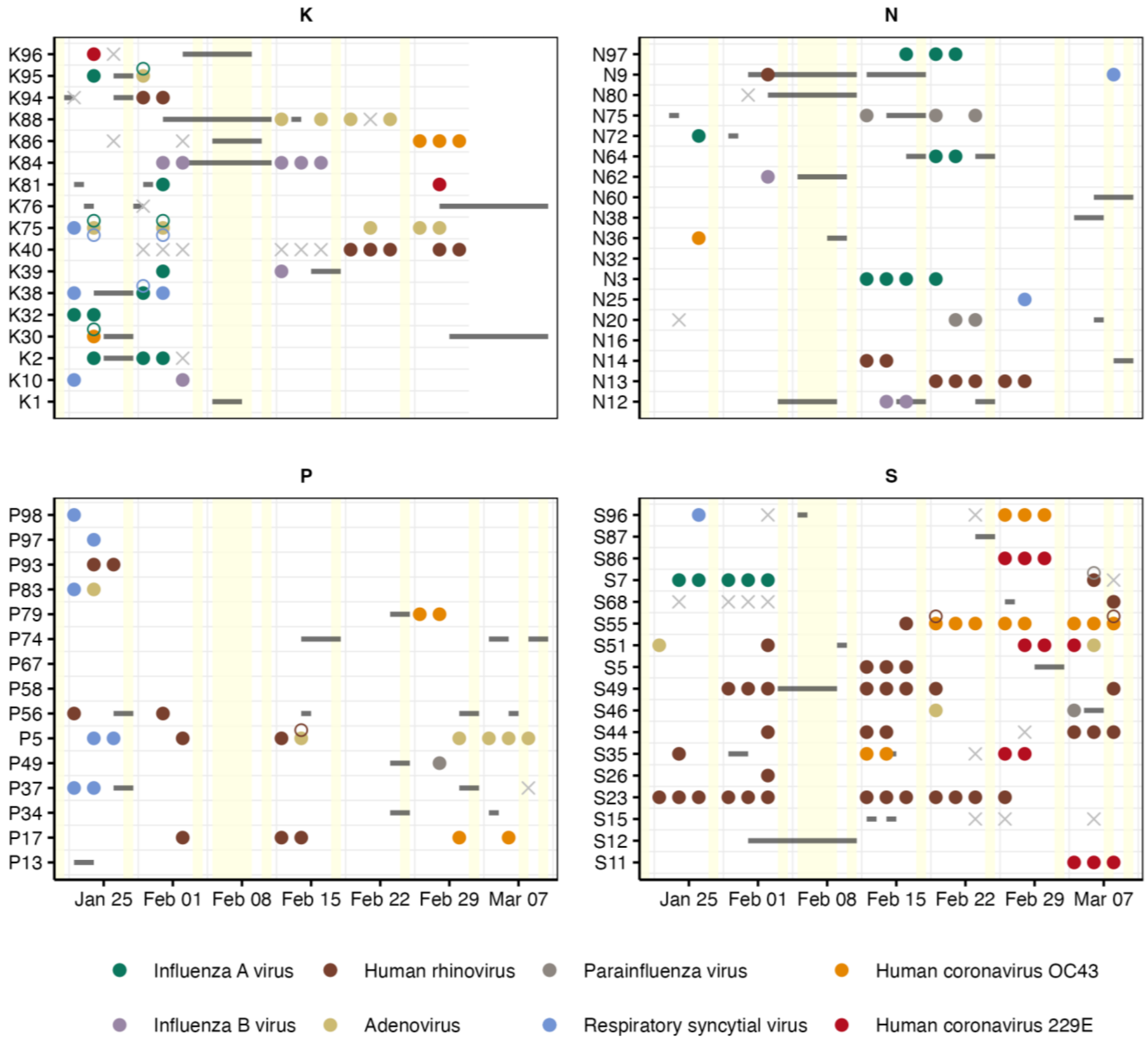

**Figure S8:** Dates of positive saliva test results (coloured dots) and absences from school (grey lines). Crosses show dates when students did not participate in saliva sampling. Yellow areas mark school-free days.

#### B.2 Supplementary results

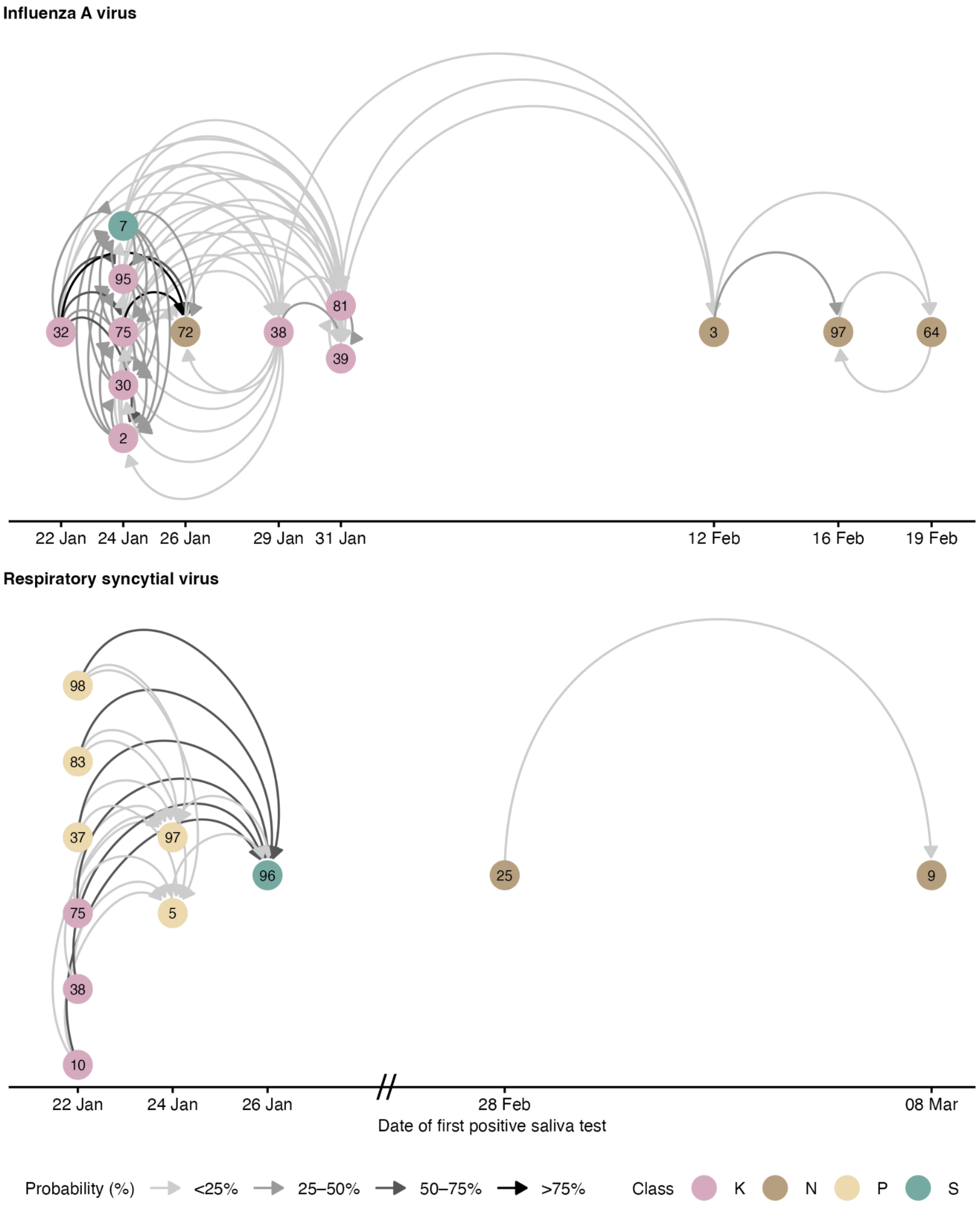

**Figure S9:** Transmission networks of influenza A and respiratory syncytial virus without excluding implausible links through genomic analysis. The probability is the proportion of paired datasets in which the link was present, considering uncertainty in pathogen-specific incubation and infectious periods. The temporal axis shows the date of the first positive saliva sample, which can be several days after the date of infection, so that circular links are possible.

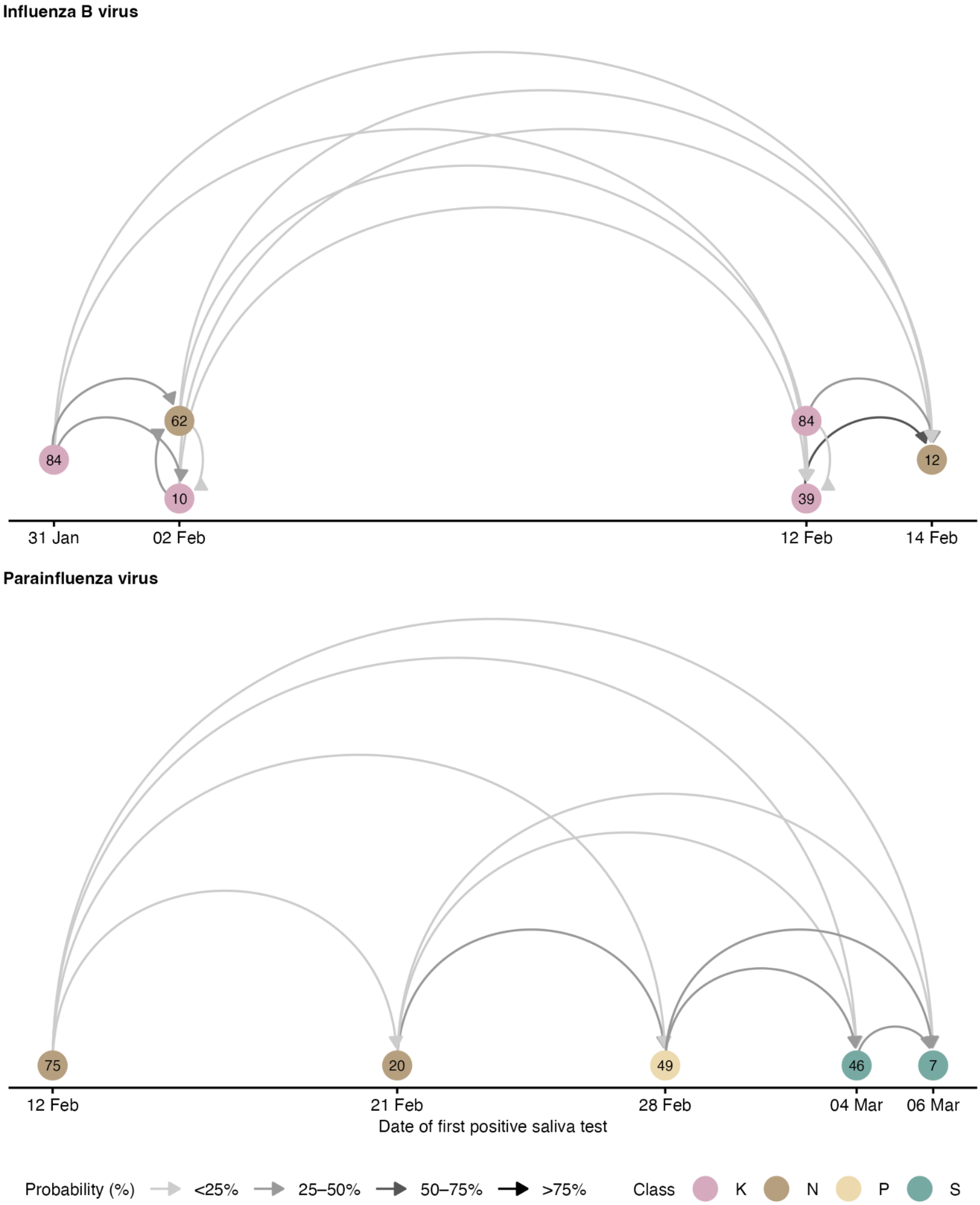

**Figure S10:** Transmission networks of influenza B and parainfluenza virus. The probability is the proportion of paired datasets in which the link was present, considering uncertainty in pathogen-specific incubation and infectious periods. The temporal axis shows the date of the first positive saliva sample, which can be several days after the date of infection, so that circular links are possible.

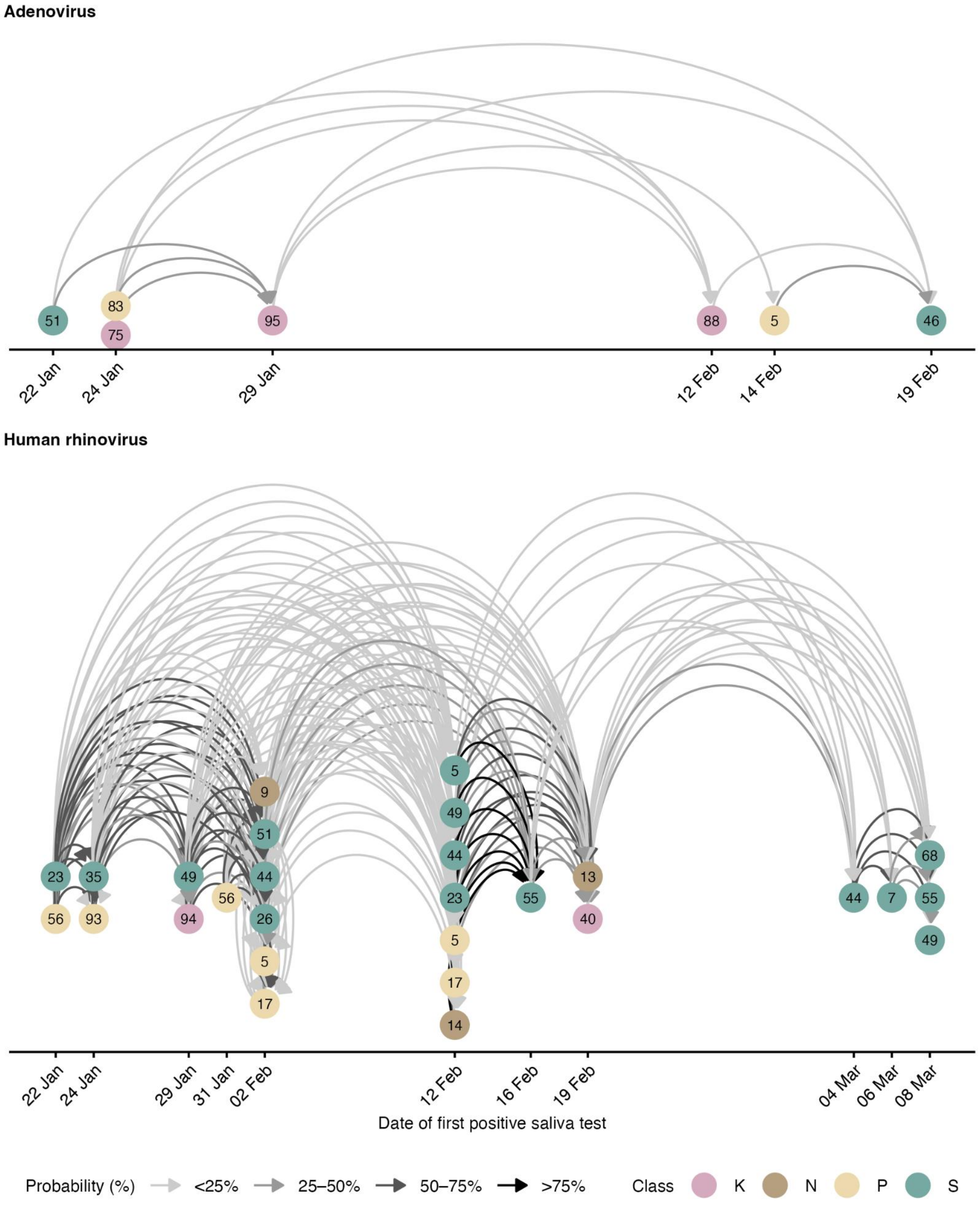

**Figure S11:** Transmission networks of human rhino and adenovirus. The probability is the proportion of paired datasets in which the link was present, considering uncertainty in pathogen-specific incubation and infectious periods. The temporal axis shows the date of the first positive saliva sample, which can be several days after the date of infection, so that circular links are possible.

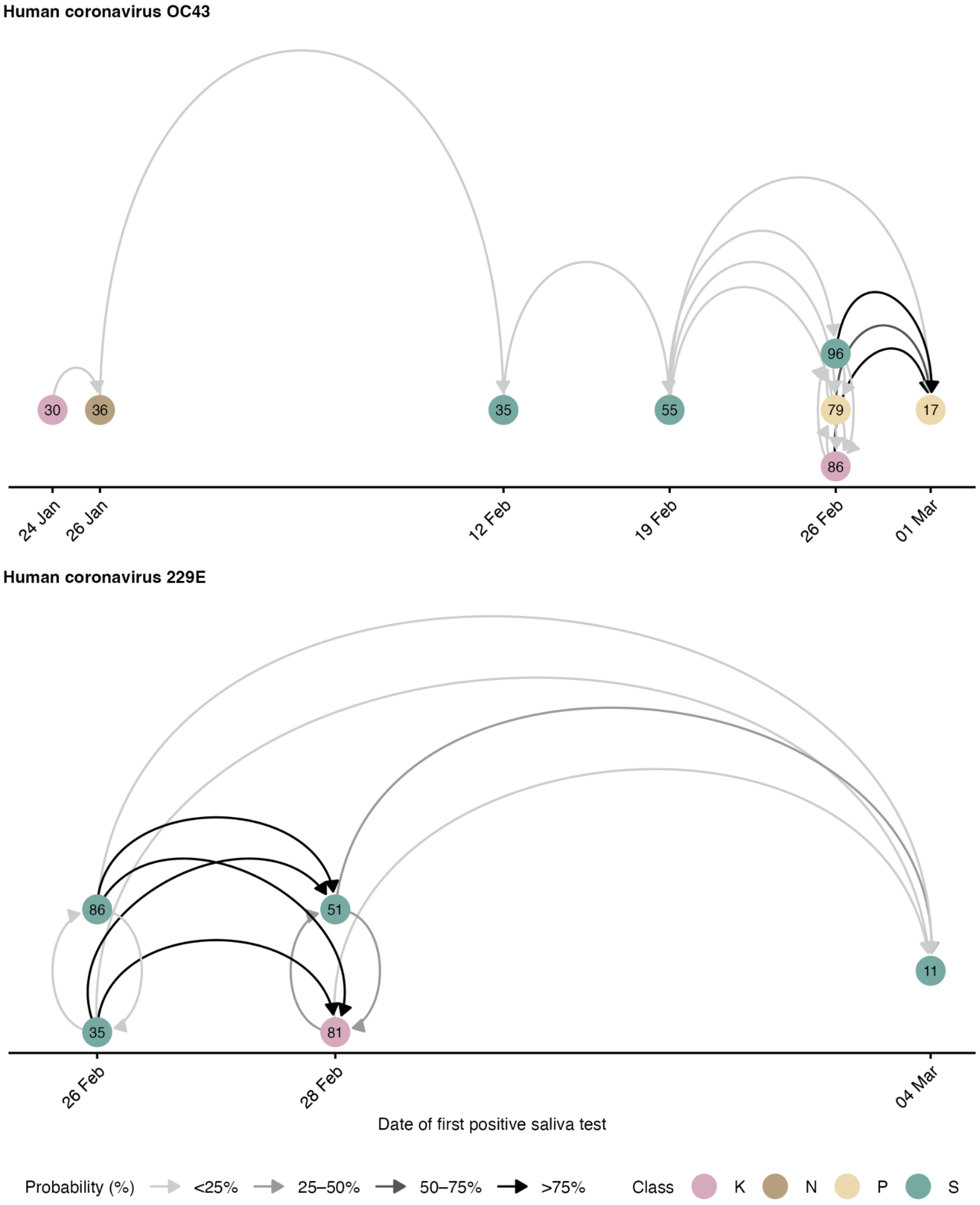

**Figure S12:** Transmission networks of human coronaviruses HCoV-OC43 and HCoV-229E. The probability is the proportion of paired datasets in which the link was present, considering uncertainty in pathogen-specific incubation and infectious periods. The temporal axis shows the date of the first positive saliva sample, which can be several days after the date of infection, so that circular links are possible.

**Table S1:** Rate ratios (median of the mean and 95%-confidence interval [CI] across paired datasets) of transmission risk factors from univariable models considering all transmissions within and between classes and only transmissions within classes. For time in close proximity, time in the shared classroom, and time with low indoor air quality, the rate ratio refers to a doubling in the time. The effects of time in the same classroom and time with low indoor air quality could not be assessed (n.a.) for all transmissions. The effect of being in the same class could not be assessed for transmissions only within classes. For social factors, we estimated the effects on internal and external transmission rates.

| **Variable** | **All transmissions** | **Only within classes** |
|---|---|---|
| | Mean (95%-CI) | Mean (95%-CI) |
| **Physical proximity** | | – |
| Daily time in close proximity (log sec) | | |
| Inf.-weighted average during exposure | 1.16 (1.01–1.33) | 1.09 (0.84–1.41) |
| Unweighted average during exposure | 1.07 (0.90–1.27) | 0.85 (0.69–1.06) |
| Overall median during exposure | 1.12 (0.94–1.34) | 0.95 (0.79–1.14) |
| **Shared classroom** | | |
| Students in the same class | 4.02 (1.69–9.64) | n.a. |
| Time in the same classroom (log min) | n.a. | 8.96 (4.85–16.88) |
| **Indoor air quality** | | |
| Daily time with low indoor air quality (log min) | | |
| $CO_2$ >1,000 ppm in classroom | n.a. | 5.59 (3.25 – 9.83) |
| $CO_2$ >1,400 ppm in classroom | n.a. | 2.62 (1.53 – 4.46) |
| $PM_{2.5}$ > 5 µg/m³ in classroom | n.a. | 2.83 (1.49 – 5.24) |
| **Social factors** | | |
| Susceptible has siblings (internal) | 1.81 (0.38–7.57) | 1.15 (0.22–4.94) |
| Susceptible has extracurricular activities (internal) | 1.69 (0.73–3.89) | 1.03 (0.38–2.83) |
| Susceptible has siblings (external) | 0.36 (0.06–2.44) | 0.80 (0.14–5.51) |
| Susceptible has extracurricular activities (external) | 0.55 (0.19–1.61) | 1.05 (0.32–3.41) |

*Notes: Time in close proximity in the main analysis was measured by the daily infectiousness-weighted average time during the exposure period and indoor air quality by the daily time $CO_2$ > 1,000 ppm in the classroom. Other measures were considered as part of a sensitivity analysis. Further note that $PM_{2.5}$ concentration (µg/m³) was only measured in the rooms of classes K and S.*

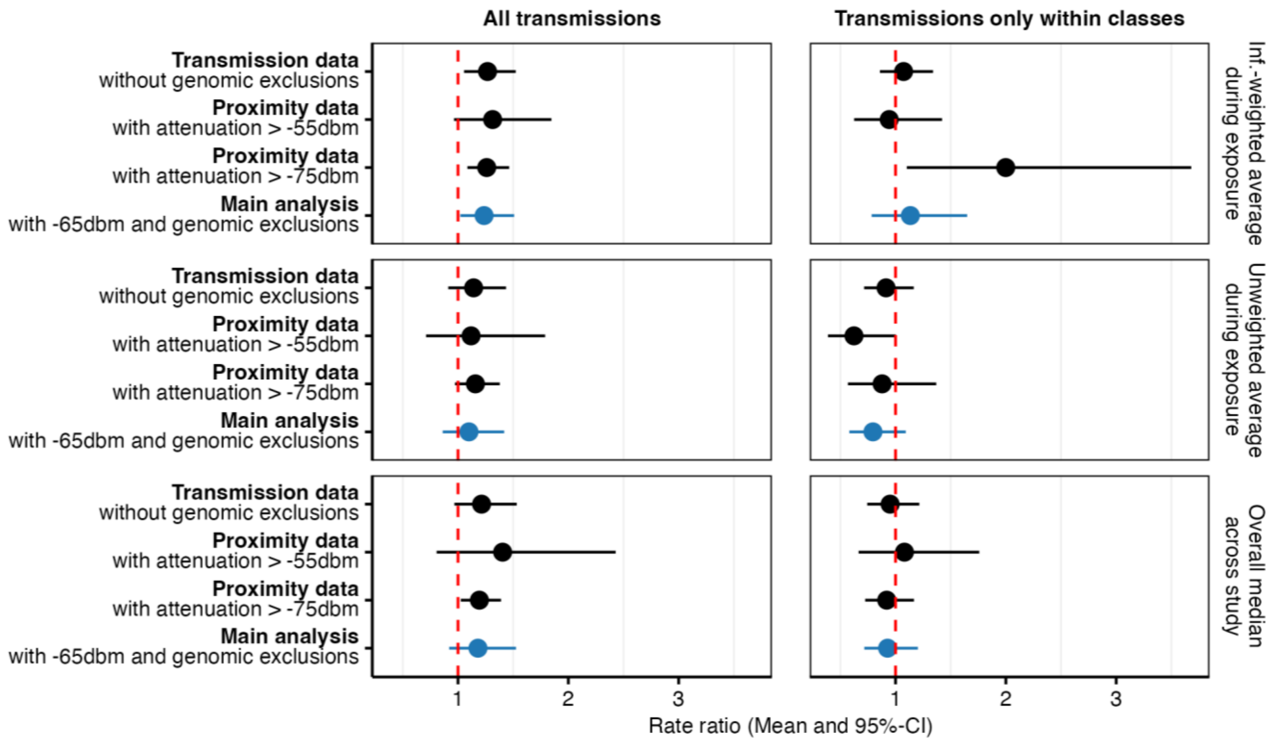

**Figure S13:** Sensitivity analysis showing the rate ratio (means as dots and 95%-confidence intervals [CIs] as lines) for the effects of close-proximity time on transmission using data without genomic exclusions for influenza A and respiratory syncytial virus and with different attenuation thresholds for detecting close-proximity contacts with the wearable sensors. Main analysis highlighted in blue on data with genomic exclusions and an attenuation threshold of -65dbm.

#### B.3 Supplementary illustrations for methods

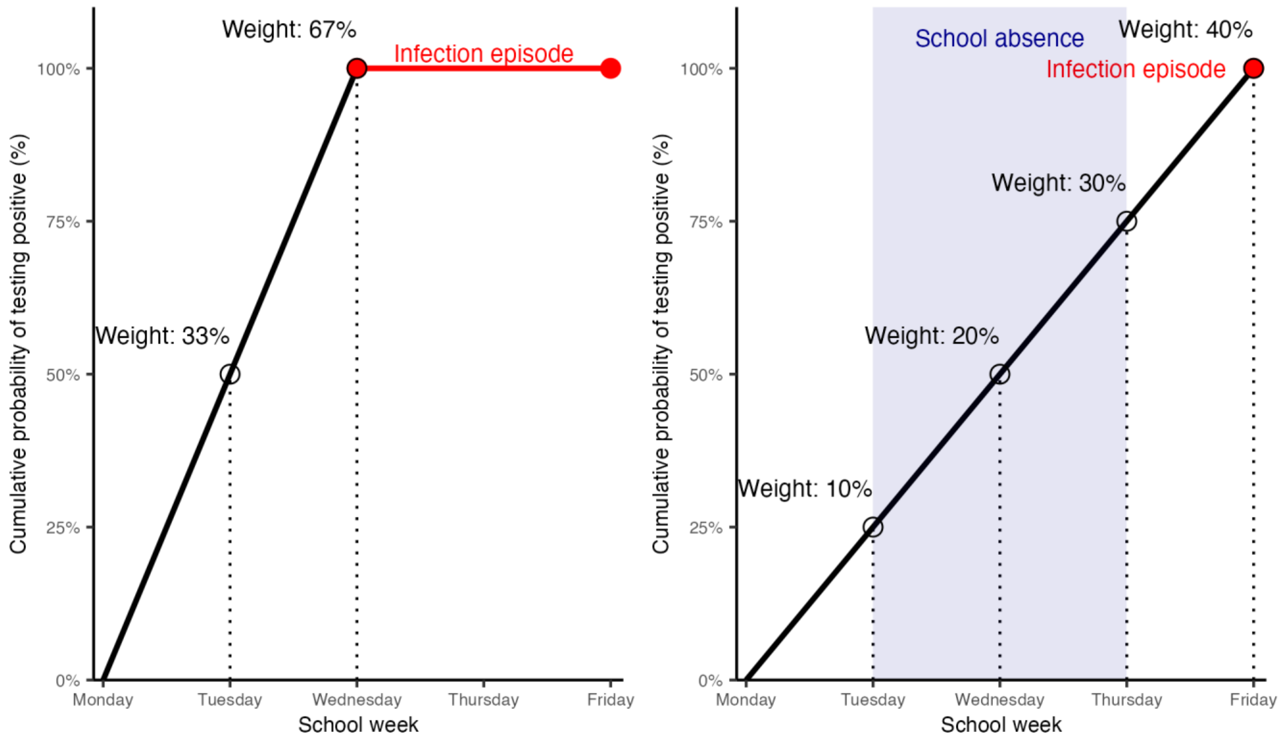

**Figure S14:** Illustration of probabilistic sampling for the first possible positive saliva test result (start of infectiousness). Left: Student first tested positive on Wednesday with first possible date on Tuesday, corresponding to sampling weights of 33% for Tuesday and 67% for Wednesday. Right: Student first tested positive on Friday but was absent from school from Tuesday to Thursday, therefore the weights are 10%, 20%, 30%, and 40% for Tuesday to Friday, respectively.

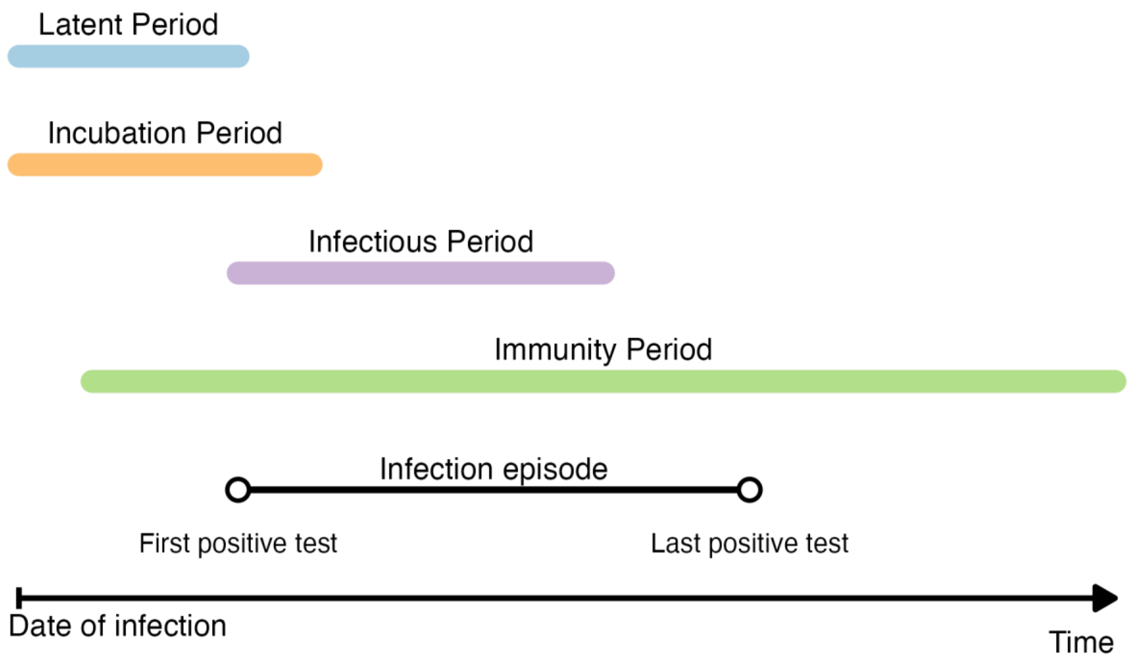

**Figure S15:** Illustration of the incubation, latent, infectious, and immunity period as derived from infection episodes defined by a period of consecutive positive saliva test results.

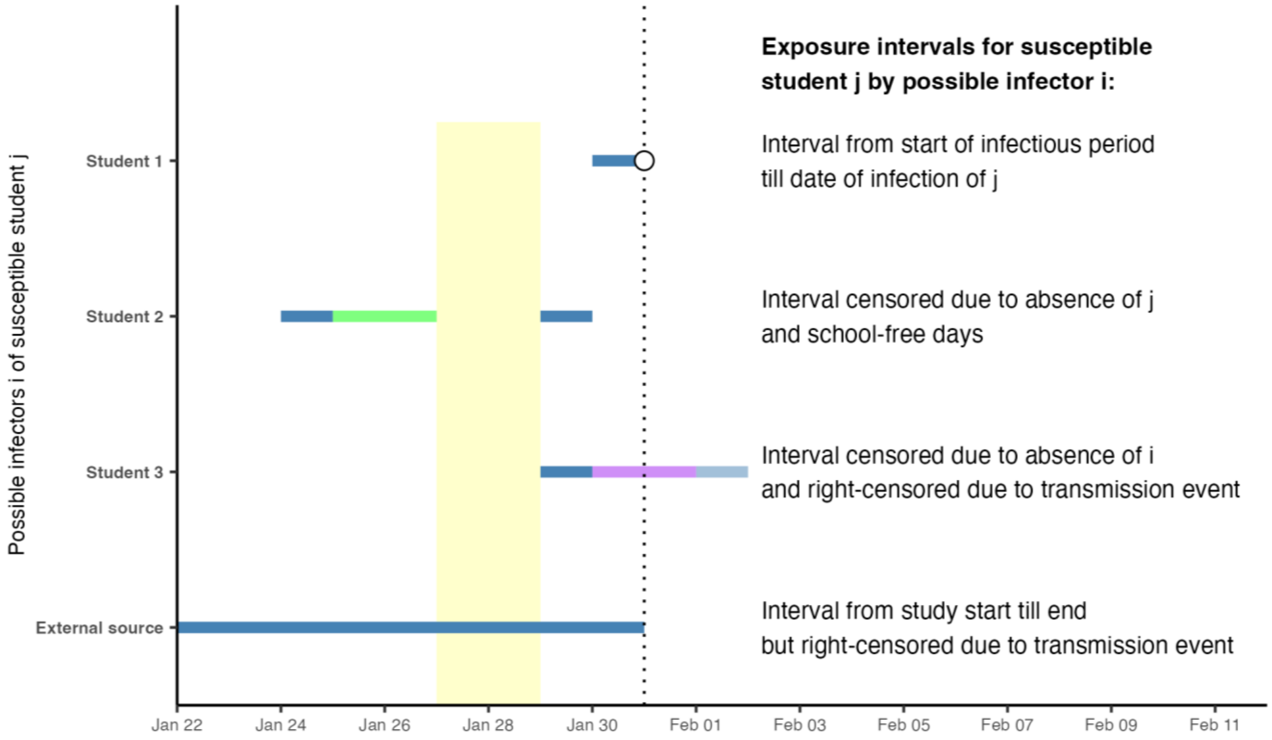

**Figure S16:** Illustration of different types of exposure intervals (blue lines) for a susceptible student j with varying infectors i. Date of infection shown as dot, weekend as yellow area, absence period of susceptible student j as green line and of infectious student i as purple line, right-censored exposure period as more transparent blue-coloured line. Note that the exposure interval is generally defined as the time the susceptible student is in school together with the infectious student. School-free days and absences can shorten the exposure interval, except for external sources to which the susceptible student is always considered exposed. Right-censoring can occur due prior infection by another student.

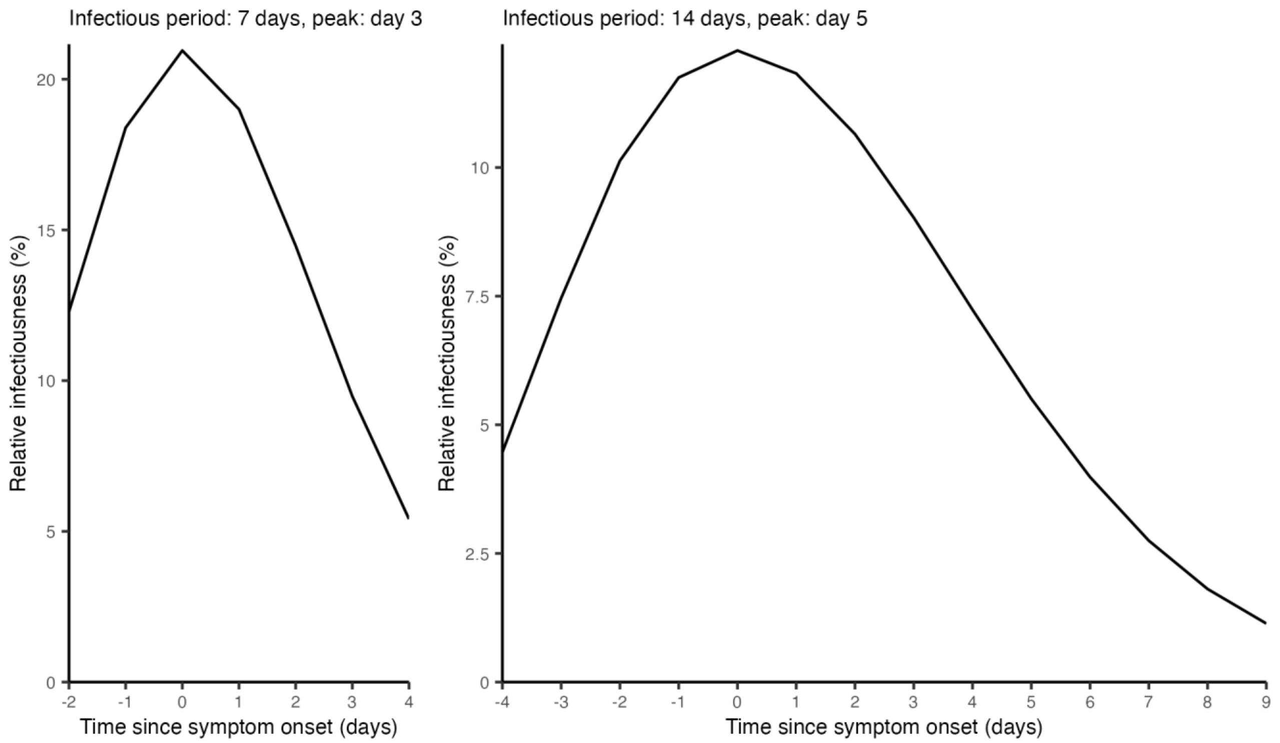

**Figure S17:** Illustration of the relative infectiousness for two infectious periods of seven and 14 days with a peak of infectiousness at days three and five, the days of symptom onset, respectively.

**Table S2:** Illustration of the impact of weighting time in close proximity by relative infectiousness. Shown are the daily average infectiousness-weighted times during the exposure period for three susceptible individuals A, B, and C. Each susceptible has a cumulative time of five minutes during the exposure period. Susceptible A has one minute on every day, B was not exposed (n.e.) on the first three days, while C was not exposed during the last three days. The infectious individual had an infectious period of five days with symptom onset (peak of infectiousness) on the second day.

| | **Close contact times during infectious period** | | | | | **Total and average time during exposure period** | | |
|---|---|---|---|---|---|---|---|---|
| **Susceptible** | Day 1 | Day 2 (peak) | Day 3 | Day 4 | Day 5 | Total | Unweighted average | Infectiousness-weighted average |
| **A** | 1 | 1 | 1 | 1 | 1 | 5 | 1 | 1 |
| **B** | n.e. | n.e. | n.e. | 2 | 3 | 5 | 2.5 | 1.14 |
| **C** | 2 | 3 | n.e. | n.e. | n.e. | 5 | 2.5 | 3.53 |

### C Genomic sequence accession codes

#### C.1 Accession Codes for sequences used as targets in sequencing protocol

IAV:

NC_007373.1-start: 1

NC_007373.1-end: 2341

NC_007372.1-start: 2342

NC_007372.1-end: 4682

NC_007371.1-start: 4683

NC_007371.1-end: 6915

NC_007366.1-start: 6916

NC_007366.1-end: 8677

NC_007369.1-start: 8678

NC_007369.1-end: 10243

NC_007368.1-start: 10244

NC_007368.1-end: 11710

NC_007367.1-start: 11711

NC_007367.1-end: 12737

NC_007372.1-start: 12738

NC_007372.1-end: 15078

IBV:

CY040455.1-start: 1

CY040455.1-end: 2315

CY040456.1-start: 2316

CY040456.1-end: 4675

CY040454.1-start: 4676

CY040454.1-end: 6927

CY040449.1-start: 6928

CY040449.1-end: 8754

CY040452.1-start: 8755

CY040452.1-end: 10530

CY040451.1-start: 10531

CY040451.1-end: 12015

CY040450.1-start: 12016

CY040450.1-end: 13159

CY040453.1-start: 13160

CY040453.1-end: 14196

<u>H1N1:</u>

NC_026431.1-start: 1

NC_026431.1-end: 982

NC_026432.1-start: 983

NC_026432.1-end: 1845

NC_026433.1-start: 1846

NC_026433.1-end: 3546

NC_026434.1-start: 3547

NC_026434.1-end: 4956

NC_026435.1-start: 4957

NC_026435.1-end: 7230

NC_026436.1-start: 7231

NC_026436.1-end: 8727

NC_026437.1-start: 8728

NC_026437.1-end: 10878

NC_026438.1-start: 10879

NC_026438.1-end: 13158

<u>SARS-CoV_2:</u>

NC_045512.2-start: 1

NC_045512.2-end: 29903

<u>RSV A:</u>

OP890336.1-start: 1

OP890336.1-end: 15261

<u>RSV B:</u>

OP965707.1-start: 1

OP965707.1-end: 15278

##### C.2 Accession codes for sequences from the Swiss Respiratory Virus Sequencing study

**Table S3:** Accession codes for the BioProject PRJEB83635, of genomic sequences from the Swiss Respiratory Virus Sequencing study used for the Influenza A NA phylogenetic tree

| accession | date | country |
|---|---|---|
| xwkVbK | 27.12.2023 | Switzerland |
| kwbwf8 | 26.12.2023 | Switzerland |
| wVBQbT | 29.05.2024 | Switzerland |
| 3qDrEd | 19.04.2023 | Switzerland |
| h2ZJd5 | 31.01.2024 | Switzerland |
| ydSpNQ | 16.01.2024 | Switzerland |
| qtZxkz | 15.01.2024 | Switzerland |
| Sm9GYA | 12.01.2024 | Switzerland |
| 32WNFL | 07.12.2023 | Switzerland |
| g33df2 | 09.01.2024 | Switzerland |
| gkcF5C | 31.01.2024 | Switzerland |
| xrRPFj | 18.01.2024 | Switzerland |
| PZQaiq | 20.02.2024 | Switzerland |
| 3edUBd | 30.11.2023 | Switzerland |
| b99ooj | 27.02.2024 | Switzerland |
| Rtjhba | 24.01.2024 | Switzerland |
| VA9Yi5 | 10.01.2024 | Switzerland |
| qeHUBn | 11.04.2024 | Switzerland |
| SQZYHn | 17.01.2024 | Switzerland |
| XEuJCy | 22.12.2023 | Switzerland |
| PGoRgN | 20.02.2024 | Switzerland |
| V5iihT | 18.01.2024 | Switzerland |
| QqAEGg | 22.02.2024 | Switzerland |
| gF3E5u | 22.02.2024 | Switzerland |
| SKJgwA | 18.01.2024 | Switzerland |
| FXFt6J | 04.03.2024 | Switzerland |
| gaQkyT | 06.03.2024 | Switzerland |
| YowhiU | 16.02.2024 | Switzerland |
| 4SnyCD | 10.04.2023 | Switzerland |
| K2udH5 | 30.01.2024 | Switzerland |
| xHGR9u | 17.01.2024 | Switzerland |
| jtjCjd | 27.12.2023 | Switzerland |
| iDqyV9 | 25.01.2024 | Switzerland |
| HtiJwT | 19.02.2024 | Switzerland |

| | | |
|---|---|---|
| 3H4uLZ | 01.02.2024 | Switzerland |
| 375EUk | 08.12.2023 | Switzerland |
| mc3GSV | 28.12.2023 | Switzerland |
| C983Zw | 19.02.2024 | Switzerland |
| 4UF9K6 | 28.12.2023 | Switzerland |
| f8rfBe | 04.03.2024 | Switzerland |
| qWgDtq | 04.06.2024 | Switzerland |
| 4L3pXZ | 27.01.2024 | Switzerland |
| WrGEQp | 27.02.2024 | Switzerland |
| YYRzxL | 21.02.2024 | Switzerland |
| 8QXJTp | 28.01.2024 | Switzerland |
| UqhsDL | 23.12.2023 | Switzerland |
| hozdkG | 15.02.2024 | Switzerland |
| sFFUfJ | 14.03.2024 | Switzerland |
| itJLqU | 29.12.2023 | Switzerland |
| w9v3Qv | 12.01.2024 | Switzerland |
| oLXZ9a | 17.01.2024 | Switzerland |
| m6243p | 17.01.2024 | Switzerland |
| q2oXnd | 09.01.2024 | Switzerland |
| URVR7h | 19.01.2024 | Switzerland |
| MHWQwo | 11.01.2024 | Switzerland |
| opfXjQ | 11.01.2024 | Switzerland |
| 9HhvMh | 18.01.2024 | Switzerland |
| x6NRYc | 16.01.2024 | Switzerland |
| LosGHn | 28.01.2024 | Switzerland |
| ZAyjrq | 18.12.2023 | Switzerland |
| 39kc9U | 19.01.2024 | Switzerland |
| 2b8YpQ | 16.02.2024 | Switzerland |
| 4t7Ncr | 31.01.2024 | Switzerland |
| RyXauM | 27.12.2023 | Switzerland |
| 9eyPAv | 30.03.2024 | Switzerland |
| ztFGoy | 02.04.2024 | Switzerland |
| juVwzj | 01.02.2024 | Switzerland |
| kBgpZ4 | 16.01.2024 | Switzerland |
| SpXNmN | 29.01.2024 | Switzerland |
| 3VC6ft | 30.01.2024 | Switzerland |
| jHbrsp | 20.02.2024 | Switzerland |
| yhE88h | 01.03.2024 | Switzerland |
| bpDXvz | 26.04.2024 | Switzerland |

| | | |
|---|---|---|
| DwiwwU | 19.01.2024 | Switzerland |
| JFKsEJ | 20.02.2024 | Switzerland |
| kNU7js | 15.02.2024 | Switzerland |
| 4rvG7f | 19.01.2024 | Switzerland |
| BnbHLr | 26.01.2024 | Switzerland |
| LCBdav | 31.01.2024 | Switzerland |
| sr4bvM | 02.02.2024 | Switzerland |
| 6z2wf2 | 07.03.2024 | Switzerland |
| uTpHe5 | 24.01.2024 | Switzerland |
| AcEG8S | 18.02.2024 | Switzerland |
| uSTYZ2 | 04.03.2024 | Switzerland |
| DgRuyg | 14.02.2024 | Switzerland |
| YBrMoY | 16.02.2024 | Switzerland |
| DMLKnU | 15.02.2024 | Switzerland |
| gyUNUP | 19.02.2024 | Switzerland |
| sAyJww | 27.02.2024 | Switzerland |
| 2hKeW2 | 27.02.2024 | Switzerland |
| j3ZXpL | 29.02.2024 | Switzerland |
| TPgies | 11.03.2024 | Switzerland |
| 5cJWyf | 13.03.2024 | Switzerland |
| xYnaUy | 16.01.2024 | Switzerland |
| F2HifQ | 18.01.2024 | Switzerland |
| 3p7s5a | 20.03.2024 | Switzerland |
| TPsF65 | 01.03.2024 | Switzerland |
| CLwScw | 28.12.2023 | Switzerland |
| eAr6Lo | 15.01.2024 | Switzerland |
| ss4Fh8 | 19.01.2024 | Switzerland |
| mmYEfJ | 01.02.2024 | Switzerland |
| fhoncL | 02.02.2024 | Switzerland |
| gsuEGK | 16.02.2024 | Switzerland |
| 8ochGa | 10.06.2024 | Switzerland |
| FHPnrx | 17.01.2024 | Switzerland |
| hzNQRG | 01.02.2024 | Switzerland |

**Table S4:** Accession codes for the BioProject PRJEB83635, of the genomic sequences from the Swiss Respiratory Virus Sequencing study used for the RSV G gene phylogenetic tree

| accession | date | country |
|---|---|---|
| wF4k2T | 07.03.2024 | Switzerland |
| zGrf4n | 20.11.2023 | Switzerland |
| nf9ieP | 24.01.2024 | Switzerland |
| MT4r8p | 20.12.2023 | Switzerland |
| HXGr8V | 28.12.2023 | Switzerland |
| 2MHu4K | 13.03.2024 | Switzerland |
| ahT4kF | 29.01.2024 | Switzerland |
| v3KDUF | 27.12.2023 | Switzerland |
| 3oZJ5x | 24.11.2023 | Switzerland |
| NDEjHn | 17.02.2024 | Switzerland |
| iZ7TLc | 12.03.2024 | Switzerland |
| M5iqYC | 30.01.2024 | Switzerland |
| sVxWai | 17.02.2024 | Switzerland |
| CgiSAY | 20.03.2024 | Switzerland |
| 9eyPAv | 30.03.2024 | Switzerland |
| FcN3wC | 05.03.2024 | Switzerland |
| Di2qm7 | 27.02.2024 | Switzerland |
| xPAa2d | 22.12.2023 | Switzerland |
| XisgNp | 27.12.2023 | Switzerland |
| 33eCP4 | 06.12.2023 | Switzerland |
| afGSAT | 04.03.2024 | Switzerland |
| RCdbJH | 21.02.2024 | Switzerland |
| 6RPWyT | 06.03.2024 | Switzerland |
| 8YYVHP | 24.11.2023 | Switzerland |
| TCsq7r | 02.02.2024 | Switzerland |
| 3fLBSP | 24.11.2023 | Switzerland |
| zwqknf | 23.11.2023 | Switzerland |
| Ki8Aa3 | 12.03.2024 | Switzerland |
| GpvELC | 19.01.2024 | Switzerland |
| 4SRYNi | 21.11.2023 | Switzerland |
| 3eGjGj | 07.12.2023 | Switzerland |
| hHKDyB | 15.02.2024 | Switzerland |
| Kr6366 | 10.04.2024 | Switzerland |
| qFQ9kJ | 10.04.2024 | Switzerland |
| Q8Ba9A | 30.01.2024 | Switzerland |
| gnDemF | 27.12.2023 | Switzerland |

| | | |
|---|---|---|
| Zu3u7N | 29.12.2023 | Switzerland |
| yebiMQ | 23.02.2024 | Switzerland |
| mzwDVC | 10.01.2024 | Switzerland |
| AnXy9n | 16.02.2024 | Switzerland |
| GbMzJy | 16.02.2024 | Switzerland |
| 5fSMj3 | 07.03.2024 | Switzerland |
| DBKZHX | 04.03.2024 | Switzerland |
| Qf8cLC | 22.02.2024 | Switzerland |
| T5VXtD | 30.01.2024 | Switzerland |
| pzbDPq | 29.02.2024 | Switzerland |
| zxhtR4 | 29.01.2024 | Switzerland |
| HAAUgG | 27.12.2023 | Switzerland |
| 4JSa55 | 29.11.2023 | Switzerland |
| 4i2tZF | 29.12.2023 | Switzerland |
| k9vgiw | 31.01.2024 | Switzerland |
| HXHQAT | 20.12.2023 | Switzerland |
| G86emu | 21.02.2024 | Switzerland |
| 8ERNNJ | 07.12.2023 | Switzerland |
| 3fGJE8 | 30.11.2023 | Switzerland |
| G8vc2N | 02.02.2024 | Switzerland |
| NmHGDa | 29.12.2023 | Switzerland |
| ND22rx | 04.03.2024 | Switzerland |
| SrvHnQ | 22.02.2024 | Switzerland |

##### C.3 Accession codes for sequences from the Swiss Respiratory Virus Sequencing study

**Table S5:** Accession codes for GenBank, of the global genomic sequences used for the Influenza A NA phylogenetic tree

| accession | date | country |
|---|---|---|
| GS_002EHP8.1 | 01.01.2009 | USA |
| GS_00286TP.1 | 23.03.2010 | Nicaragua |
| GS_0022ULT.1 | 12.01.2010 | USA |
| GS_001YQFG.1 | 22.05.2009 | Taiwan |
| GS_0022RTG.1 | 01.11.2009 | South Korea |
| GS_0026P8F.1 | 01.10.2009 | USA |
| GS_0040EYK.1 | 12.03.2010 | USA |
| GS_003U2RP.1 | 28.01.2011 | China |
| GS_003RQ88.1 | 01.08.2010 | Taiwan |
| GS_0028PFX.1 | 26.01.2011 | USA |
| GS_00255SY.1 | 07.12.2009 | Poland |
| GS_003VFDZ.1 | 24.03.2010 | Singapore |
| GS_001ZB87.1 | 23.06.2009 | Argentina |
| GS_002EZRN.1 | 26.06.2009 | Singapore |
| GS_002ES9U.1 | 07.07.2009 | Singapore |
| GS_002CU0E.1 | 05.08.2009 | Hong Kong |
| GS_00233VY.1 | 18.01.2010 | USA |
| GS_0021EDQ.1 | 18.10.2009 | USA |
| GS_00AAQFJ.1 | 01.01.2017 | Colombia |
| GS_002F67G.1 | 11.01.2011 | Singapore |
| GS_006S8LU.1 | 07.09.2011 | Uganda |
| GS_0041B49.1 | 25.12.2010 | USA |
| GS_002CQDS.1 | 27.01.2011 | USA |
| GS_00585CJ.1 | 30.11.2010 | United Kingdom |
| GS_0041VV8.1 | 20.03.2012 | USA |
| GS_0054YPA.1 | 08.07.2012 | Brazil |
| GS_002PNE3.1 | 21.09.2012 | Thailand |
| GS_0041SH1.1 | 02.07.2012 | USA |
| GS_003SBJY.1 | 25.01.2011 | Russia |
| GS_002G9DZ.1 | 26.01.2011 | Estonia |
| GS_0090MH3.1 | 01.02.2011 | USA |
| GS_002G9HQ.1 | 15.02.2011 | China |
| GS_0029GPK.1 | 27.08.2010 | Thailand |
| GS_0041SLV.1 | 18.01.2012 | USA |

| | | |
|---|---|---|
| GS_0041298.1 | 07.03.2011 | Iran |
| GS_004QZWP.1 | 28.09.2010 | Singapore |
| GS_0041SW9.1 | 07.02.2012 | USA |
| GS_0041VNN.1 | 09.04.2012 | USA |
| GS_0041WSD.1 | 12.03.2012 | USA |
| GS_0041SX7.1 | 09.02.2012 | USA |
| GS_0041VQJ.1 | 02.03.2012 | USA |
| GS_0045VWY.1 | 19.02.2013 | Russia |
| GS_0045FVE.1 | 10.04.2013 | USA |
| GS_00454XM.1 | 05.02.2013 | Finland |
| GS_002RKRD.1 | 30.06.2013 | Nicaragua |
| GS_005510J.1 | 21.08.2013 | Brazil |
| GS_002RMD2.1 | 25.12.2013 | USA |
| GS_004AXCP.1 | 20.11.2013 | USA |
| GS_0040P5X.1 | 04.02.2014 | USA |
| GS_0045H9K.1 | 08.08.2013 | USA |
| GS_004RH5L.1 | 19.12.2013 | USA |
| GS_004AQUX.1 | 04.02.2014 | Finland |
| GS_004AR75.1 | 17.02.2014 | Finland |
| GS_002RNZU.1 | 10.03.2014 | USA |
| GS_002RPA6.1 | 09.04.2014 | USA |
| GS_00484NZ.1 | 30.12.2013 | USA |
| GS_0055JH0.1 | 08.03.2014 | Japan |
| GS_0055JJY.1 | 29.01.2014 | Japan |
| GS_002Z03T.1 | 25.11.2015 | Nicaragua |
| GS_004SG5K.1 | 07.01.2015 | USA |
| GS_0051CAV.1 | 19.03.2015 | State of Palestine |
| GS_004UT0E.1 | 05.11.2015 | USA |
| GS_004X4NL.1 | 04.10.2015 | Kenya |
| GS_004USN4.1 | 07.09.2015 | USA |
| GS_004ZVCC.1 | 11.04.2016 | USA |
| GS_004WAS8.1 | 30.12.2015 | USA |
| GS_002WYJ4.1 | 12.05.2017 | USA |
| GS_002V6AF.1 | 05.01.2017 | USA |
| GS_004W6TA.1 | 17.12.2015 | USA |
| GS_002WXPV.1 | 26.02.2017 | USA |
| GS_002VVTR.1 | 09.02.2017 | USA |
| GS_002WJDV.1 | 27.02.2017 | USA |

| | | |
|---|---|---|
| GS_004ZVVB.1 | 29.04.2016 | USA |
| GS_00513FU.1 | 20.01.2016 | USA |
| GS_004XV6U.1 | 28.02.2016 | USA |
| GS_004USZG.1 | 31.10.2015 | USA |
| GS_004W7D4.1 | 25.11.2015 | USA |
| GS_004XR50.1 | 18.01.2016 | USA |
| GS_00515MD.1 | 08.02.2016 | USA |
| GS_004XN61.1 | 12.02.2016 | USA |
| GS_0054366.1 | 21.03.2016 | USA |
| GS_004Y3HV.1 | 15.04.2016 | USA |
| GS_002V64T.1 | 26.12.2016 | USA |
| GS_002VWCM.1 | 24.02.2017 | USA |
| GS_006CH93.1 | 01.02.2019 | USA |
| GS_006FTLY.1 | 11.03.2019 | USA |
| GS_007B8ZY.1 | 14.02.2020 | USA |
| GS_002XGFR.1 | 16.07.2017 | USA |
| GS_002YZH1.1 | 03.09.2017 | USA |
| GS_006Z28B.1 | 30.09.2018 | USA |
| GS_006LS9B.1 | 14.02.2019 | USA |
| GS_00976K0.1 | 14.12.2018 | USA |
| GS_0066QS7.1 | 07.01.2019 | USA |
| GS_0069HHS.1 | 29.12.2018 | USA |
| GS_006BM75.1 | 11.01.2019 | USA |
| GS_005HXW4.1 | 21.02.2018 | USA |
| GS_005P1DP.1 | 11.04.2018 | USA |
| GS_005YL60.1 | 03.10.2018 | USA |
| GS_00602T5.1 | 06.08.2018 | USA |
| GS_005HWTB.1 | 23.02.2018 | USA |
| GS_005LHG7.1 | 09.03.2018 | USA |
| GS_002ZQK4.1 | 08.12.2017 | USA |
| GS_00615VW.1 | 14.11.2018 | USA |
| GS_006Y55G.1 | 15.06.2019 | USA |
| GS_006BJZP.1 | 03.01.2019 | South Korea |
| GS_0079B9E.1 | 21.03.2019 | China |
| GS_007B7E4.1 | 25.01.2020 | USA |
| GS_007EA95.1 | 05.10.2019 | USA |
| GS_00AND4S.1 | 01.01.2020 | USA |
| GS_007AXQV.1 | 03.02.2020 | USA |
| GS_0077BE8.1 | 29.10.2019 | USA |

| | | |
|---|---|---|
| GS_0079PR3.1 | 08.01.2020 | USA |
| GS_0087SJH.1 | 24.02.2021 | USA |
| GS_00ANJW2.1 | 01.02.2020 | USA |
| GS_007B6RG.1 | 23.01.2020 | USA |
| GS_00ANE9F.1 | 12.01.2020 | USA |
| GS_007AXCL.1 | 03.02.2020 | USA |
| GS_007BNN6.1 | 03.03.2020 | USA |
| GS_007NYSZ.1 | 18.09.2021 | USA |
| GS_0089SL9.1 | 09.07.2021 | USA |
| GS_007NYR1.1 | 22.09.2021 | USA |
| GS_007SFT6.1 | 24.10.2021 | USA |
| GS_008VQ35.1 | 21.08.2022 | USA |
| GS_009V71R.1 | 26.10.2022 | United Kingdom |
| GS_007P9LY.1 | 17.09.2021 | USA |
| GS_007P9SL.1 | 17.09.2021 | USA |
| GS_007R422.1 | 19.11.2021 | USA |
| GS_007P9QQ.1 | 19.11.2021 | USA |
| GS_007R430.1 | 19.11.2021 | USA |
| GS_009Y9VU.1 | 10.10.2023 | USA |
| GS_009XNEE.1 | 06.12.2023 | USA |
| GS_009WL28.1 | 26.10.2023 | USA |
| GS_009Z7KE.1 | 30.11.2023 | USA |
| GS_009WTK0.1 | 30.10.2023 | USA |
| GS_009Z2EW.1 | 08.11.2023 | USA |
| GS_00A0BJA.1 | 06.09.2023 | Thailand |
| GS_009XA2G.1 | 26.12.2023 | USA |
| GS_00A389S.1 | 16.02.2024 | USA |
| GS_00A528W.1 | 28.12.2023 | USA |
| GS_009WMCM.1 | 02.12.2023 | USA |
| GS_00A54AQ.1 | 21.12.2023 | USA |
| GS_009ZPWA.1 | 29.12.2023 | USA |
| GS_009ZADQ.1 | 24.10.2023 | USA |
| GS_009Z6Q5.1 | 25.10.2023 | USA |
| GS_009YPCF.1 | 26.12.2023 | Germany |
| GS_00AH8KA.1 | 26.02.2024 | USA |
| GS_008V9MH.1 | 21.11.2022 | USA |
| GS_009ABD2.1 | 27.02.2023 | USA |
| GS_009270E.1 | 16.12.2022 | USA |

| | | |
|---|---|---|
| GS_008ZHCL.1 | 19.12.2022 | Germany |
| GS_00AVXBE.1 | 13.02.2023 | USA |
| GS_00A9VPY.1 | 08.12.2023 | USA |
| GS_008V001.1 | 11.10.2022 | USA |
| GS_0091MZ3.1 | 09.01.2023 | USA |
| GS_009SPP4.1 | 25.09.2023 | Japan |
| GS_009SPQ2.1 | 01.10.2023 | Japan |
| GS_009A67K.1 | 28.01.2023 | USA |
| GS_008VSYC.1 | 31.10.2022 | USA |
| GS_009ABHT.1 | 28.02.2023 | USA |
| GS_009HYS7.1 | 05.08.2023 | USA |
| GS_009P17X.1 | 03.08.2023 | USA |
| GS_009P18V.1 | 03.08.2023 | USA |
| GS_009QKT4.1 | 13.12.2022 | USA |
| GS_009J3GM.1 | 24.06.2023 | USA |
| GS_009ZBFK.1 | 19.10.2023 | USA |
| GS_009VEES.1 | 27.06.2023 | Australia |
| GS_009ZKDF.1 | 19.09.2023 | USA |
| GS_009WUWB.1 | 26.11.2023 | USA |
| GS_009X9R4.1 | 24.12.2023 | USA |
| GS_009Z72F.1 | 03.10.2023 | USA |
| GS_009ZCVP.1 | 23.11.2023 | USA |
| GS_009Y694.1 | 14.12.2023 | USA |
| GS_009J2FQ.1 | 30.07.2023 | USA |
| GS_009EHM5.1 | 24.07.2023 | USA |
| GS_009A4Y4.1 | 08.02.2023 | USA |
| GS_009A96J.1 | 15.02.2023 | USA |
| GS_00A0VJS.1 | 10.08.2023 | USA |
| GS_00A103G.1 | 30.01.2024 | United Kingdom |
| GS_009YAGL.1 | 08.11.2023 | USA |
| GS_00A5145.1 | 28.12.2023 | USA |
| GS_009DK54.1 | 02.06.2023 | USA |
| GS_009J3VU.1 | 30.06.2023 | USA |
| GS_00A3FKY.1 | 31.12.2023 | USA |
| GS_0096ESE.1 | 07.12.2022 | USA |
| GS_009HJNV.1 | 04.01.2023 | USA |
| GS_0096FW5.1 | 08.12.2022 | USA |
| GS_009QLKK.1 | 25.01.2023 | USA |

| | | |
|---|---|---|
| GS_009VFWS.1 | 01.06.2023 | United Kingdom |
| GS_009VFQ4.1 | 01.06.2023 | United Kingdom |
| GS_009VLM5.1 | 01.06.2023 | United Kingdom |
| GS_00A5CTF.1 | 17.11.2023 | USA |
| GS_009WKRW.1 | 08.11.2023 | USA |
| GS_00A5B9K.1 | 04.02.2024 | USA |
| GS_009WUBG.1 | 05.11.2023 | Japan |
| GS_009W58B.1 | 28.11.2023 | United Kingdom |
| GS_009J581.1 | 16.07.2023 | USA |
| GS_009ZHH8.1 | 01.11.2023 | USA |
| GS_009ZKZ8.1 | 05.10.2023 | USA |
| GS_00A4WCW.1 | 26.12.2023 | USA |
| GS_00A51XJ.1 | 03.01.2024 | USA |
| GS_009QNA2.1 | 03.09.2023 | USA |
| GS_009ZECN.1 | 19.10.2023 | USA |
| GS_009XN32.1 | 03.12.2023 | USA |
| GS_009ZBUS.1 | 08.10.2023 | USA |
| GS_00A45RU.1 | 19.12.2023 | USA |
| GS_00A656V.1 | 11.12.2023 | USA |
| GS_009Z7UW.1 | 27.09.2023 | USA |
| GS_009X894.1 | 29.12.2023 | USA |
| GS_00A3GDA.1 | 07.12.2023 | USA |
| GS_00A043E.1 | 17.12.2023 | USA |
| GS_009Z9AX.1 | 02.11.2023 | USA |
| GS_00A4RS6.1 | 25.12.2023 | USA |
| GS_009ZEUP.1 | 09.11.2023 | USA |
| GS_009WKP0.1 | 09.11.2023 | USA |
| GS_009XA76.1 | 26.12.2023 | USA |
| GS_009X9ET.1 | 22.12.2023 | USA |
| GS_009WHL8.1 | 21.11.2023 | USA |
| GS_009ZPLW.1 | 29.12.2023 | USA |
| GS_009ZK5X.1 | 14.11.2023 | USA |
| GS_00A4XER.1 | 18.11.2023 | USA |
| GS_00A4UZP.1 | 04.12.2023 | USA |
| GS_009Z3T2.1 | 06.12.2023 | USA |
| GS_00A5E2W.1 | 10.11.2023 | USA |
| GS_00A5BMU.1 | 08.01.2024 | USA |

| | | |
|---|---|---|
| GS_00A5E5Q.1 | 06.12.2023 | USA |
| GS_009J4M9.1 | 05.08.2023 | USA |
| GS_009ZN8N.1 | 20.12.2023 | USA |
| GS_00A58AL.1 | 30.11.2023 | USA |
| GS_009ZBYJ.1 | 29.11.2023 | USA |
| GS_00AKXR5.1 | 02.10.2023 | USA |
| GS_009NP97.1 | 09.08.2023 | USA |
| GS_00ABD2K.1 | 09.03.2024 | USA |
| GS_009XQNV.1 | 11.12.2023 | USA |
| GS_00A0SQH.1 | 23.01.2024 | USA |
| GS_009ZBVQ.1 | 09.10.2023 | USA |
| GS_00A9UK7.1 | 05.12.2023 | USA |
| GS_009ZE44.1 | 24.10.2023 | USA |
| GS_009ZE52.1 | 25.10.2023 | USA |
| GS_00A25XL.1 | 12.02.2024 | United Kingdom |
| GS_009Z8CU.1 | 01.12.2023 | USA |
| GS_00A4ZSY.1 | 26.11.2023 | USA |
| GS_009XUCC.1 | 23.12.2023 | USA |
| GS_009WLST.1 | 27.11.2023 | USA |
| GS_009Y8ZM.1 | 28.11.2023 | USA |
| GS_009XT2Z.1 | 15.12.2023 | USA |
| GS_00A310H.1 | 17.11.2023 | Hong Kong |
| GS_00A4W4C.1 | 01.01.2024 | USA |
| GS_00A4W3E.1 | 01.01.2024 | USA |
| GS_009Z1U2.1 | 12.10.2023 | USA |
| GS_00A597R.1 | 25.11.2023 | USA |
| GS_00AFWLR.1 | 22.02.2024 | USA |
| GS_009Z3ZQ.1 | 02.11.2023 | USA |
| GS_009WU6S.1 | 08.11.2023 | USA |
| GS_00A9UL5.1 | 03.12.2023 | USA |
| GS_009Y6JK.1 | 06.12.2023 | USA |
| GS_00A5E7L.1 | 06.12.2023 | USA |
| GS_00A47TN.1 | 02.01.2024 | USA |
| GS_009Z7MA.1 | 24.09.2023 | USA |
| GS_009Z7YN.1 | 04.10.2023 | USA |
| GS_009Y6EU.1 | 10.10.2023 | Puerto Rico |
| GS_009Y6DW.1 | 11.10.2023 | Puerto Rico |
| GS_009ZKK2.1 | 16.10.2023 | USA |

| | | |
|---|---|---|
| GS_009ZG18.1 | 21.10.2023 | USA |
| GS_009ZGZB.1 | 30.10.2023 | USA |
| GS_009ZAFL.1 | 01.11.2023 | USA |
| GS_009ZBP2.1 | 02.12.2023 | USA |
| GS_00A9WL3.1 | 12.12.2023 | USA |
| GS_00A546Y.1 | 16.12.2023 | USA |
| GS_009ZNAJ.1 | 18.12.2023 | USA |
| GS_00ABEFS.1 | 01.01.2024 | USA |
| GS_00A5F4R.1 | 03.01.2024 | USA |
| GS_00A5EY3.1 | 18.12.2023 | USA |
| GS_009Z6YP.1 | 21.09.2023 | USA |
| GS_009Z288.1 | 29.10.2023 | USA |
| GS_009WGN5.1 | 13.11.2023 | USA |
| GS_009WLGE.1 | 24.11.2023 | USA |
| GS_009W4QD.1 | 30.01.2024 | Switzerland |
| GS_009X8VX.1 | 18.12.2023 | USA |
| GS_00A4X83.1 | 23.12.2023 | USA |
| GS_00A4WR3.1 | 14.12.2023 | USA |
| GS_009YR11.1 | 25.12.2023 | USA |
| GS_009ZG50.1 | 24.10.2023 | USA |
| GS_009Z42J.1 | 08.11.2023 | USA |
| GS_009ZLDE.1 | 04.12.2023 | USA |
| GS_009WP6X.1 | 08.12.2023 | USA |
| GS_00AM0BT.1 | 03.02.2024 | USA |
| GS_00ABEA2.1 | 20.11.2023 | USA |
| GS_009WULX.1 | 25.10.2023 | Japan |
| GS_00A4TDX.1 | 14.11.2023 | USA |
| GS_00A6341.1 | 05.02.2024 | USA |
| GS_00AGT9F.1 | 12.12.2023 | USA |
| GS_009ZG6Y.1 | 14.11.2023 | USA |
| GS_009XN24.1 | 02.12.2023 | USA |
| GS_00A0EVK.1 | 04.01.2024 | USA |
| GS_009ZN12.1 | 15.12.2023 | USA |
| GS_009YNK1.1 | 03.01.2024 | United Kingdom |
| GS_00A3GH1.1 | 22.12.2023 | USA |
| GS_0098FKP.1 | 10.02.2023 | China |
| GS_009YNJ3.1 | 19.12.2023 | United Kingdom |
| GS_00AWU9K.1 | 05.01.2025 | USA |

| | | |
|---|---|---|
| GS_009PTYN.1 | 02.05.2023 | China |
| GS_009PTXQ.1 | 27.04.2023 | China |
| GS_009W41S.1 | 29.01.2024 | Switzerland |
| GS_00A4642.1 | 19.12.2023 | USA |
| GS_009J5FM.1 | 18.07.2023 | USA |
| GS_00A0Z5F.1 | 30.01.2024 | Germany |
| GS_00A91CF.1 | 06.03.2024 | Germany |
| GS_009WTEB.1 | 24.10.2023 | USA |
| GS_009EGQ0.1 | 02.06.2023 | Japan |
| GS_009YATX.1 | 16.11.2023 | USA |
| GS_009YAJF.1 | 13.11.2023 | USA |
| GS_009Y8UX.1 | 07.12.2023 | USA |
| GS_00A4H8H.1 | 29.02.2024 | USA |
| GS_009YPG7.1 | 03.01.2024 | Japan |
| GS_00A1QQH.1 | 18.01.2024 | USA |
| GS_00A1RE3.1 | 24.01.2024 | USA |
| GS_00A2KUC.1 | 06.02.2024 | USA |
| GS_00A916T.1 | 19.03.2024 | South Korea |
| GS_00A3ENT.1 | 15.12.2023 | USA |
| GS_009XBJG.1 | 13.12.2023 | USA |
| GS_009XBYN.1 | 17.12.2023 | USA |
| GS_009ZQ3W.1 | 31.12.2023 | USA |
| GS_00A3FSJ.1 | 08.01.2024 | USA |
| GS_00A0S0Y.1 | 15.01.2024 | USA |
| GS_00ACVF9.1 | 13.05.2024 | Germany |
| GS_00ABPSU.1 | 06.05.2024 | Germany |
| GS_00ABPRW.1 | 06.05.2024 | Germany |
| GS_00ABP9V.1 | 06.05.2024 | Germany |
| GS_009YQ9L.1 | 15.12.2023 | United Kingdom |
| GS_00A0XV2.1 | 28.12.2023 | Germany |
| GS_009W4FY.1 | 25.01.2024 | Switzerland |
| GS_009VR5Z.1 | 12.02.2024 | United Kingdom |
| GS_00A3GRK.1 | 19.01.2024 | USA |
| GS_009YQ12.1 | 14.12.2023 | USA |
| GS_00A5T4D.1 | 01.03.2024 | USA |
| GS_00A5QR7.1 | 04.03.2024 | USA |
| GS_00A2D23.1 | 21.02.2024 | USA |
| GS_00A331D.1 | 28.01.2024 | USA |

| | | |
|---|---|---|
| GS_00A0XRA.1 | 29.01.2024 | Germany |
| GS_00A0XU4.1 | 31.01.2024 | Germany |
| GS_00A2XU0.1 | 01.01.2024 | Peru |
| GS_00A2XT2.1 | 01.01.2024 | Peru |
| GS_00A27TS.1 | 29.12.2023 | Peru |
| GS_00A27P0.1 | 28.12.2023 | Peru |
| GS_00A27XJ.1 | 12.01.2024 | Peru |
| GS_009X9WU.1 | 26.12.2023 | USA |
| GS_00A2183.1 | 29.12.2023 | USA |
| GS_00A5US1.1 | 05.02.2024 | USA |
| GS_00A91MW.1 | 08.03.2024 | USA |
| GS_00A9P57.1 | 15.02.2024 | USA |
| GS_00A2G9L.1 | 15.02.2024 | USA |
| GS_009Z53F.1 | 18.10.2023 | USA |
| GS_00AM18Y.1 | 29.01.2024 | USA |
| GS_00A0EJ7.1 | 01.01.2024 | USA |
| GS_00A2MHY.1 | 26.02.2024 | USA |
| GS_00AGY9A.1 | 01.04.2024 | USA |
| GS_009YS10.1 | 27.12.2023 | USA |
| GS_00AGT2V.1 | 01.04.2024 | USA |
| GS_00AJ4M8.1 | 01.07.2024 | Japan |
| GS_009VEHK.1 | 02.11.2023 | United Kingdom |
| GS_00A9U1A.1 | 12.02.2024 | USA |
| GS_00A0YNF.1 | 01.02.2024 | United Kingdom |
| GS_00A0K21.1 | 08.01.2024 | USA |
| GS_00A0XS8.1 | 30.01.2024 | Germany |
| GS_009W45J.1 | 05.02.2024 | Switzerland |
| GS_00A9RVQ.1 | 20.01.2024 | USA |
| GS_00A5AUF.1 | 11.12.2023 | USA |
| GS_00A61SS.1 | 15.02.2024 | USA |
| GS_009W3D3.1 | 04.03.2024 | Switzerland |
| GS_00A4HX4.1 | 21.02.2024 | United Kingdom |
| GS_00AH24D.1 | 20.03.2024 | USA |
| GS_009WHXL.1 | 29.11.2023 | USA |
| GS_009WPPW.1 | 10.12.2023 | USA |
| GS_009J4P5.1 | 18.07.2023 | USA |
| GS_00A2XR6.1 | 01.01.2024 | Peru |
| GS_00A5A4W.1 | 16.11.2023 | USA |

| | | |
|---|---|---|
| GS_009SR63.1 | 29.08.2023 | USA |
| GS_009ZA0G.1 | 10.10.2023 | USA |
| GS_009Z5ZN.1 | 23.10.2023 | USA |
| GS_009WU04.1 | 01.11.2023 | USA |
| GS_00A4TQ8.1 | 21.12.2023 | USA |
| GS_00A60UP.1 | 02.01.2024 | USA |
| GS_009Z5WU.1 | 16.10.2023 | USA |
| GS_009Z63E.1 | 30.10.2023 | USA |
| GS_00A4XAZ.1 | 10.11.2023 | USA |
| GS_00A9U36.1 | 12.02.2024 | USA |
| GS_009ZJ08.1 | 12.11.2023 | USA |
| GS_00A2W0Q.1 | 13.02.2024 | USA |
| GS_00A2XJL.1 | 01.01.2024 | Peru |
| GS_00A2Y5C.1 | 01.01.2024 | Peru |
| GS_00A2Y86.1 | 01.01.2024 | Peru |
| GS_00AH3UX.1 | 04.03.2024 | USA |
| GS_00A2PQG.1 | 02.03.2024 | USA |
| GS_00A7T0H.1 | 13.03.2024 | USA |
| GS_00AFWC8.1 | 28.12.2023 | USA |
| GS_00A1WKM.1 | 19.01.2024 | USA |
| GS_00A1RHW.1 | 25.01.2024 | USA |
| GS_00AM055.1 | 04.02.2024 | USA |
| GS_00A2J8L.1 | 13.02.2024 | USA |
| GS_00A2WU1.1 | 22.02.2024 | USA |
| GS_00A38EG.1 | 16.02.2024 | USA |
| GS_00A5EAE.1 | 14.12.2023 | USA |
| GS_00A9U9U.1 | 20.02.2024 | USA |
| GS_00AH673.1 | 20.02.2024 | USA |
| GS_00A1US9.1 | 12.02.2024 | USA |
| GS_00ABGYP.1 | 15.04.2024 | USA |
| GS_009Y6KH.1 | 07.12.2023 | Germany |
| GS_00AHB8W.1 | 06.03.2024 | USA |
| GS_00A55GB.1 | 18.12.2023 | USA |
| GS_009YPXA.1 | 22.12.2023 | USA |
| GS_009X80N.1 | 28.12.2023 | USA |
| GS_00A0KXA.1 | 13.01.2024 | USA |
| GS_00A0SBA.1 | 19.01.2024 | USA |
| GS_00A0WRB.1 | 07.02.2024 | USA |
| GS_00A2WDY.1 | 22.02.2024 | USA |

| | | |
|---|---|---|
| GS_00AH7Q1.1 | 02.03.2024 | USA |
| GS_00AGZ2P.1 | 15.03.2024 | USA |
| GS_00AF0HT.1 | 15.04.2024 | USA |
| GS_00ABMRY.1 | 28.03.2024 | USA |
| GS_00A03HL.1 | 29.11.2023 | USA |
| GS_00A60PZ.1 | 12.12.2023 | USA |
| GS_00A0GPV.1 | 16.01.2024 | USA |
| GS_00A927Q.1 | 11.03.2024 | USA |
| GS_00ALW1K.1 | 29.01.2024 | USA |
| GS_00A6250.1 | 26.01.2024 | USA |
| GS_00ALZUV.1 | 30.01.2024 | USA |
| GS_00A4XUW.1 | 10.12.2023 | USA |
| GS_00A2WR7.1 | 22.02.2024 | USA |
| GS_009XNSP.1 | 11.12.2023 | USA |
| GS_00A38K5.1 | 16.02.2024 | USA |
| GS_009W3FZ.1 | 06.05.2024 | Switzerland |
| GS_00AWSCF.1 | 02.01.2025 | USA |
| GS_00A9UWK.1 | 07.12.2023 | USA |
| GS_00AGSQK.1 | 14.03.2024 | USA |
| GS_00AGUBA.1 | 10.04.2024 | USA |
| GS_009XA3E.1 | 26.12.2023 | USA |
| GS_00A5FNN.1 | 30.01.2024 | USA |
| GS_00A5B7P.1 | 31.01.2024 | USA |
| GS_00AGZ6F.1 | 04.03.2024 | USA |
| GS_00A2M0Z.1 | 19.02.2024 | USA |
| GS_009EGN4.1 | 20.06.2023 | Japan |
| GS_00A0TV6.1 | 27.01.2024 | USA |
| GS_009N2L5.1 | 22.08.2023 | USA |
| GS_009W4S9.1 | 14.03.2024 | Switzerland |
| GS_00ABNAU.1 | 25.03.2024 | USA |
| GS_009Y6UZ.1 | 04.12.2023 | Germany |
| GS_009ZNMV.1 | 28.12.2023 | USA |
| GS_00A2UZT.1 | 02.02.2024 | USA |
| GS_00AKX1L.1 | 18.02.2024 | USA |
| GS_00A2464.1 | 27.02.2024 | Germany |
| GS_00A26AU.1 | 27.02.2024 | Germany |
| GS_00A26BS.1 | 27.02.2024 | Germany |
| GS_00A26CQ.1 | 27.02.2024 | Germany |
| GS_00A24ZH.1 | 27.02.2024 | Germany |

| | | |
|---|---|---|
| GS_00A26EL.1 | 27.02.2024 | Germany |
| GS_00A24YK.1 | 27.02.2024 | Germany |
| GS_00A26FJ.1 | 28.02.2024 | Germany |
| GS_00A250F.1 | 28.02.2024 | Germany |
| GS_00AETMU.1 | 08.12.2023 | USA |
| GS_00ALYYN.1 | 29.07.2024 | USA |
| GS_009ZJEE.1 | 09.10.2023 | USA |
| GS_00AKTJP.1 | 27.03.2024 | USA |
| GS_009ZF19.1 | 30.10.2023 | USA |
| GS_009ZF0B.1 | 30.10.2023 | USA |
| GS_00A5R2K.1 | 06.03.2024 | USA |
| GS_009WNRT.1 | 28.11.2023 | USA |
| GS_009YA74.1 | 30.10.2023 | USA |
| GS_00A1UA8.1 | 08.01.2024 | USA |
| GS_00A47QU.1 | 29.12.2023 | USA |
| GS_009XTWA.1 | 19.12.2023 | USA |
| GS_00A54CL.1 | 23.12.2023 | USA |
| GS_00A9W26.1 | 04.02.2024 | USA |
| GS_00A1TFZ.1 | 29.01.2024 | USA |
| GS_00A2XKJ.1 | 01.01.2024 | Peru |
| GS_00A9YFC.1 | 20.12.2023 | USA |
| GS_00A3GF6.1 | 07.12.2023 | USA |
| GS_009XSX9.1 | 14.12.2023 | USA |
| GS_00A5BNS.1 | 14.01.2024 | USA |
| GS_00A6614.1 | 01.02.2024 | USA |
| GS_00A2FMW.1 | 11.02.2024 | USA |
| GS_00A2KSG.1 | 05.02.2024 | USA |
| GS_009WHWN.1 | 28.11.2023 | USA |
| GS_00ALW77.1 | 28.01.2024 | USA |
| GS_00AETJ0.1 | 16.03.2024 | USA |
| GS_00AWU8M.1 | 05.01.2025 | USA |
| GS_00AWWW8.1 | 03.01.2025 | USA |
| GS_00AWUKY.1 | 06.01.2025 | USA |
| GS_00A9XZA.1 | 11.02.2024 | USA |
| GS_00A52N1.1 | 25.11.2023 | USA |
| GS_009WPGB.1 | 09.12.2023 | USA |
| GS_00AH6LA.1 | 26.02.2024 | USA |
| GS_00A0THW.1 | 25.01.2024 | USA |
| GS_009Z2R7.1 | 20.11.2023 | USA |

| | | |
|---|---|---|
| GS_00A62PX.1 | 16.01.2024 | USA |
| GS_009WP8T.1 | 08.12.2023 | USA |
| GS_009XB91.1 | 20.12.2023 | USA |
| GS_00ALW69.1 | 07.02.2024 | USA |
| GS_009ZA64.1 | 20.11.2023 | USA |
| GS_00A5DD9.1 | 18.11.2023 | USA |
| GS_00A9VSS.1 | 02.02.2024 | USA |
| GS_00A4JF2.1 | 01.03.2024 | USA |
| GS_00A7VUS.1 | 08.03.2024 | USA |
| GS_00A4UVX.1 | 03.12.2023 | USA |
| GS_009ZNKZ.1 | 16.12.2023 | USA |
| GS_00A5D1Z.1 | 31.12.2023 | USA |
| GS_009ZKL0.1 | 15.10.2023 | USA |
| GS_009YABW.1 | 06.11.2023 | USA |
| GS_00ABE5C.1 | 20.11.2023 | USA |
| GS_00A4YM9.1 | 02.12.2023 | USA |
| GS_009XU3W.1 | 21.12.2023 | USA |
| GS_00A4725.1 | 23.12.2023 | USA |
| GS_00A0CDL.1 | 08.01.2024 | USA |
| GS_00A2PE3.1 | 01.03.2024 | USA |
| GS_00A61CM.1 | 13.02.2024 | USA |
| GS_00A61EH.1 | 29.01.2024 | USA |
| GS_00AH459.1 | 12.03.2024 | USA |
| GS_00A61GD.1 | 07.02.2024 | USA |
| GS_00A2FQQ.1 | 11.02.2024 | USA |
| GS_009X95B.1 | 19.12.2023 | USA |
| GS_00A2CSP.1 | 19.02.2024 | USA |
| GS_00A2T7D.1 | 25.02.2024 | USA |
| GS_00A9QJD.1 | 24.02.2024 | USA |
| GS_00A3A9Q.1 | 04.12.2023 | USA |
| GS_00A3FE9.1 | 15.12.2023 | USA |
| GS_00AF16F.1 | 22.05.2024 | USA |
| GS_00A5UWT.1 | 14.02.2024 | USA |
| GS_00A51BR.1 | 14.01.2024 | USA |
| GS_00AGSW7.1 | 20.03.2024 | USA |
| GS_00A2NV7.1 | 28.02.2024 | USA |
| GS_00ABCR7.1 | 14.03.2024 | USA |
| GS_00AH5P5.1 | 04.04.2024 | USA |
| GS_00A3CLZ.1 | 27.01.2024 | USA |

| | | |
|---|---|---|
| GS_00AAEFU.1 | 01.02.2024 | USA |
| GS_00A61M2.1 | 04.02.2024 | USA |
| GS_00A9VL4.1 | 31.01.2024 | USA |
| GS_00ALC05.1 | 22.10.2024 | USA |
| GS_00A24P3.1 | 05.02.2024 | Germany |
| GS_009X7JL.1 | 20.12.2023 | USA |
| GS_009ZNCE.1 | 22.12.2023 | USA |
| GS_00A1U6G.1 | 16.12.2023 | USA |
| GS_00A66J3.1 | 09.02.2024 | USA |
| GS_00A2547.1 | 13.02.2024 | Italy |
| GS_00A26N3.1 | 13.02.2024 | Italy |
| GS_00AWUAH.1 | 05.01.2025 | USA |
| GS_00AGTLS.1 | 24.03.2024 | USA |
| GS_00AKP5L.1 | 09.07.2024 | USA |
| GS_00AUQXG.1 | 07.10.2024 | USA |
| GS_00AKRQD.1 | 22.04.2024 | USA |
| GS_00AEZGZ.1 | 05.05.2024 | USA |
| GS_00A5BJ0.1 | 03.01.2024 | USA |
| GS_00A5DY4.1 | 12.01.2024 | USA |
| GS_00A64L1.1 | 26.01.2024 | USA |
| GS_00AGVC7.1 | 11.04.2024 | USA |
| GS_00ABMUS.1 | 22.04.2024 | USA |
| GS_00AGEQX.1 | 15.07.2024 | USA |
| GS_00AGXA9.1 | 24.04.2024 | USA |
| GS_00ALY4B.1 | 30.08.2024 | USA |
| GS_00AKUMG.1 | 25.06.2024 | USA |
| GS_00AKRC4.1 | 02.07.2024 | USA |
| GS_00AKTV1.1 | 05.07.2024 | USA |
| GS_00AWS7R.1 | 02.01.2025 | USA |
| GS_00AHCXG.1 | 31.05.2024 | USA |
| GS_00AUR42.1 | 02.10.2024 | USA |
| GS_00AUQJ8.1 | 22.10.2024 | USA |
| GS_00ASLYN.1 | 16.11.2024 | USA |
| GS_00AUE3F.1 | 29.10.2024 | USA |
| GS_00AM1GG.1 | 26.08.2024 | USA |
| GS_009W3X0.1 | 30.01.2024 | Switzerland |
| GS_00A2WT3.1 | 20.02.2024 | USA |
| GS_00A2SW1.1 | 16.02.2024 | USA |
| GS_00A26HD.1 | 06.02.2024 | Germany |

| | | |
|---|---|---|
| GS_00A0KAK.1 | 08.01.2024 | USA |
| GS_009W39B.1 | 31.01.2024 | Switzerland |
| GS_00A0682.1 | 03.01.2024 | USA |
| GS_00A063C.1 | 29.12.2023 | USA |
| GS_009W2YZ.1 | 31.01.2024 | Switzerland |
| GS_009W4YX.1 | 19.02.2024 | Switzerland |
| GS_009YRG5.1 | 26.12.2023 | USA |
| GS_00A269W.1 | 20.02.2024 | Germany |
| GS_009W4C4.1 | 22.01.2024 | Switzerland |
| GS_009W43N.1 | 23.01.2024 | Switzerland |
| GS_00A2E06.1 | 06.01.2024 | USA |
| GS_00AEQ9N.1 | 20.03.2024 | USA |
| GS_00AGT1X.1 | 15.04.2024 | USA |
| GS_00AWQ23.1 | 09.08.2024 | USA |
| GS_00ALNJS.1 | 23.09.2024 | USA |
| GS_00AMLRC.1 | 21.10.2024 | Germany |
| GS_00AMLPG.1 | 21.10.2024 | Germany |
| GS_00AR5PP.1 | 27.10.2024 | USA |
| GS_00AWSAK.1 | 02.01.2025 | USA |
| GS_00AUF1J.1 | 28.10.2024 | USA |
| GS_00AWUE9.1 | 06.01.2025 | USA |
| GS_00ASB9A.1 | 14.11.2024 | USA |
| GS_009W4B6.1 | 11.03.2024 | Switzerland |
| GS_00AWQ15.1 | 02.08.2024 | USA |
| GS_00AETQN.1 | 02.05.2024 | USA |
| GS_009W3KQ.1 | 02.02.2024 | Switzerland |
| GS_00A4ZZJ.1 | 14.12.2023 | USA |
| GS_009W32R.1 | 16.02.2024 | Switzerland |
| GS_00AFXHW.1 | 16.07.2024 | USA |
| GS_00AKQZW.1 | 09.07.2024 | USA |
| GS_00ALYS0.1 | 18.08.2024 | USA |
| GS_00A2670.1 | 06.02.2024 | Germany |
| GS_00A2753.1 | 22.02.2024 | United Kingdom |
| GS_00AL8VJ.1 | 30.09.2024 | USA |
| GS_00AUEZN.1 | 04.11.2024 | USA |
| GS_00AW4QC.1 | 17.12.2024 | USA |
| GS_00AWUCD.1 | 05.01.2025 | USA |
| GS_00AW8CZ.1 | 25.11.2024 | USA |

| | | |
|---|---|---|
| GS_00AVGGK.1 | 23.11.2024 | USA |
| GS_00AURAQ.1 | 04.10.2024 | USA |
| GS_00AUQWJ.1 | 05.10.2024 | USA |
| GS_009W3ML.1 | 22.01.2024 | Switzerland |
| GS_009W4E0.1 | 24.01.2024 | Switzerland |
| GS_00ALYDT.1 | 26.02.2024 | USA |
| GS_00AM0L8.1 | 23.07.2024 | USA |
| GS_009YQBG.1 | 26.12.2023 | United Kingdom |
| GS_009W4HT.1 | 15.01.2024 | Switzerland |
| GS_009W4GW.1 | 15.01.2024 | Switzerland |
| GS_009W3ZW.1 | 26.02.2024 | Switzerland |
| GS_00AW5R9.1 | 16.12.2024 | USA |
| GS_00AWRPT.1 | 29.12.2024 | USA |
| GS_00AWSK0.1 | 04.01.2025 | USA |
| GS_00AWSJ2.1 | 04.01.2025 | USA |
| GS_00AWSDD.1 | 03.01.2025 | USA |
| GS_00AWSF9.1 | 03.01.2025 | USA |
| GS_00AWWUC.1 | 02.01.2025 | USA |
| GS_00AWU7P.1 | 05.01.2025 | USA |
| GS_00AWUPQ.1 | 08.01.2025 | USA |
| GS_00AMLX0.1 | 21.10.2024 | Japan |
| GS_00AKYDV.1 | 29.05.2024 | Hong Kong |
| GS_00AL263.1 | 27.09.2024 | Japan |
| GS_00AJL3V.1 | 28.08.2024 | Japan |
| GS_00AJL7M.1 | 27.08.2024 | Japan |
| GS_00AWSG7.1 | 03.01.2025 | USA |
| GS_00AMLYY.1 | 09.10.2024 | United Kingdom |
| GS_00AV897.1 | 05.11.2024 | USA |
| GS_00AWRLZ.1 | 24.12.2024 | USA |
| GS_00AWMH9.1 | 06.01.2025 | USA |
| GS_00AWSBH.1 | 02.01.2025 | USA |
| GS_00AWUG5.1 | 06.01.2025 | USA |
| GS_00AR4Y6.1 | 13.11.2024 | USA |
| GS_00AWW2X.1 | 20.11.2024 | USA |
| GS_00AV988.1 | 03.12.2024 | USA |
| GS_00AJV0S.1 | 24.08.2024 | USA |
| GS_00AWUMU.1 | 07.01.2025 | USA |
| GS_00AM1FJ.1 | 24.08.2024 | USA |

| | | |
|---|---|---|
| GS_00AR45T.1 | 24.10.2024 | USA |
| GS_00AWUQN.1 | 08.01.2025 | USA |
| GS_00AKTZT.1 | 02.07.2024 | USA |
| GS_00AWUNS.1 | 07.01.2025 | USA |
| GS_00AWWVA.1 | 02.01.2025 | USA |
| GS_00AWW9H.1 | 20.12.2024 | USA |
| GS_00ALP8C.1 | 22.10.2024 | USA |
| GS_00AW3T7.1 | 26.11.2024 | Slovakia |
| GS_00AUUQS.1 | 29.10.2024 | USA |
| GS_00AUUPU.1 | 29.10.2024 | USA |
| GS_00AWUBF.1 | 05.01.2025 | USA |
| GS_00AWUF7.1 | 06.01.2025 | USA |
| GS_00AWTNT.1 | 26.12.2024 | USA |
| GS_00AWS6T.1 | 01.01.2025 | USA |
| GS_00AWSEB.1 | 03.01.2025 | USA |
| GS_00AWS9M.1 | 02.01.2025 | USA |
| GS_00AUUCH.1 | 31.10.2024 | USA |
| GS_00AWWTE.1 | 02.01.2025 | USA |
| GS_00AWUJ0.1 | 06.01.2025 | USA |
| GS_00AWNM0.1 | 22.12.2024 | USA |
| GS_00AWS5V.1 | 01.01.2025 | USA |
| GS_00AVC0M.1 | 11.12.2024 | Germany |
| GS_00AVCDV.1 | 10.12.2024 | Germany |
| GS_00A0DHA.1 | 21.12.2023 | Italy |
| GS_00AWUDB.1 | 05.01.2025 | USA |
| GS_00A7TR0.1 | 29.03.2024 | USA |
| GS_009WJJB.1 | 25.10.2023 | USA |
| GS_00AL22B.1 | 01.10.2024 | USA |
| GS_009W49A.1 | 15.01.2024 | Switzerland |
| GS_009W4W1.1 | 21.02.2024 | Switzerland |
| GS_009W3C5.1 | 29.01.2024 | Switzerland |
| GS_00A104E.1 | 06.02.2024 | United Kingdom |
| GS_009YQDC.1 | 03.01.2024 | United Kingdom |
| GS_00A2XME.1 | 01.01.2024 | Peru |
| GS_00A2662.1 | 06.02.2024 | Germany |
| GS_009W3V4.1 | 02.02.2024 | Switzerland |
| GS_00A27UQ.1 | 04.01.2024 | Peru |
| GS_009W40U.1 | 16.01.2024 | Switzerland |

| | | |
|---|---|---|
| GS_009W4D2.1 | 15.01.2024 | Switzerland |
| GS_00A0YS7.1 | 01.02.2024 | United Kingdom |
| GS_009W3A9.1 | 06.02.2024 | Switzerland |
| GS_009VR33.1 | 06.02.2024 | United Kingdom |
| GS_009VR41.1 | 04.02.2024 | United Kingdom |
| GS_009VR25.1 | 06.02.2024 | United Kingdom |
| GS_009W4V3.1 | 26.01.2024 | Switzerland |
| GS_009W3GX.1 | 14.02.2024 | Switzerland |
| GS_00A0Z6D.1 | 24.01.2024 | Germany |
| GS_00A26K9.1 | 08.02.2024 | Germany |
| GS_009W3E1.1 | 02.04.2024 | Switzerland |
| GS_009XNPV.1 | 10.12.2023 | USA |
| GS_009W47E.1 | 13.02.2024 | Switzerland |
| GS_009W31T.1 | 19.01.2024 | Switzerland |
| GS_009W2X1.1 | 29.01.2024 | Switzerland |
| GS_00AERK0.1 | 23.01.2024 | USA |
| GS_00AWQNW.1 | 06.12.2024 | USA |
| GS_00A0WXZ.1 | 08.02.2024 | USA |
| GS_00A0U1U.1 | 28.01.2024 | USA |
| GS_00AJ4LA.1 | 01.07.2024 | Germany |
| GS_009W34M.1 | 01.03.2024 | Switzerland |
| GS_00ABH3D.1 | 01.04.2024 | USA |
| GS_00AKQX0.1 | 20.05.2024 | USA |
| GS_009WHP2.1 | 21.11.2023 | USA |
| GS_00AWUH2.1 | 06.01.2025 | USA |
| GS_009YQCE.1 | 28.12.2023 | United Kingdom |
| GS_009W4RB.1 | 30.01.2024 | Switzerland |
| GS_009W4MK.1 | 05.02.2024 | Switzerland |
| GS_00A24AW.1 | 06.02.2024 | United Kingdom |
| GS_00A249Y.1 | 06.02.2024 | United Kingdom |
| GS_009W3W2.1 | 23.02.2024 | Switzerland |
| GS_00A252B.1 | 21.02.2024 | Germany |

**Table S6:** Accession codes for GenBank, of the global genomic sequences used for the RSV G gene phylogenetic tree

| accession | date | country |
|---|---|---|
| MZ417571 | 21.09.2018 | Taiwan |
| PQ599899 | 24.01.2024 | Mexico |
| PQ599900 | 30.01.2024 | Mexico |
| PP376265 | 01.04.2019 | South Africa |
| PP376453 | 27.12.2018 | France |
| PP376578 | 11.12.2018 | Spain |
| PP376633 | 16.12.2019 | Spain |
| PP376549 | 09.12.2017 | Finland |
| PP376459 | 28.01.2018 | Finland |
| PP376546 | 08.04.2019 | Finland |
| PP376786 | 30.04.2019 | Finland |
| PP376349 | 17.08.2018 | Brazil |
| PP376278 | 10.07.2019 | Brazil |
| PP376680 | 02.06.2019 | Australia |
| PP376434 | 25.08.2019 | Australia |
| PP376682 | 02.09.2019 | Japan |
| PP376605 | 13.02.2020 | South Korea |
| PP376504 | 04.12.2019 | South Korea |
| PP376530 | 06.12.2019 | South Korea |
| PP376356 | 18.12.2019 | South Korea |
| PP376548 | 09.01.2020 | South Korea |
| KJ723474 | 08.01.1989 | USA |
| KJ723486 | 31.12.1979 | USA |
| DQ171792 | 01.01.1998 | New Zealand |
| DQ171800 | 31.12.1987 | New Zealand |
| DQ171797 | 01.01.1988 | New Zealand |
| MG642079 | 30.12.1982 | USA |
| HQ731719 | 31.12.1985 | United Kingdom |
| MG642083 | 31.12.1987 | USA |
| DQ171807 | 01.01.1994 | New Zealand |
| KU316133 | 17.01.1990 | USA |
| KU316176 | 05.04.1990 | USA |
| JX198115 | 10.01.1991 | USA |
| KP258728 | 31.12.1992 | USA |
| JX513382 | 31.08.2001 | Brazil |

| | | |
|---|---|---|
| KJ627725 | 01.01.2004 | USA |
| JX069803 | 04.03.2000 | USA |
| JX661572 | 10.02.2000 | Germany |
| PQ591393 | 28.09.2008 | United Kingdom |
| JX015485 | 04.01.2005 | Netherlands |
| JX015488 | 09.01.2006 | Netherlands |
| DQ171829 | 10.11.1997 | New Zealand |
| JX513310 | 15.01.2002 | Brazil |
| KP317949 | 13.02.2003 | Kenya |
| KT765815 | 02.04.2004 | Kenya |
| JQ901449 | 10.01.2001 | Netherlands |
| KF826827 | 26.05.2004 | Argentina |
| HQ731698 | 01.01.2006 | Thailand |
| KJ627662 | 31.12.2003 | USA |
| HQ731773 | 01.01.2007 | United Kingdom |
| HQ731774 | 01.01.2007 | United Kingdom |
| KF826828 | 04.06.2004 | Argentina |
| JX513298 | 31.12.2005 | Brazil |
| JX513309 | 31.12.2005 | Brazil |
| KF826826 | 24.12.2004 | Mexico |
| KF826841 | 02.05.2007 | Argentina |
| KF826850 | 01.01.2008 | USA |
| KU950561 | 05.01.2006 | USA |
| KF826836 | 04.01.2006 | Mexico |
| MG642077 | 17.07.1987 | USA |
| DQ171786 | 30.09.1986 | New Zealand |
| HQ731716 | 16.10.1981 | United Kingdom |
| DQ171761 | 01.01.1989 | New Zealand |
| DQ171743 | 14.08.1982 | New Zealand |
| HQ731720 | 31.12.1984 | United Kingdom |
| DQ171742 | 01.01.1988 | New Zealand |
| OR795296 | 20.03.2000 | Germany |
| KU316166 | 31.12.1977 | USA |
| KT765780 | 18.01.2000 | Kenya |
| OK649657 | 01.01.1995 | Canada |
| MG642050 | 01.01.1994 | USA |
| KU316118 | 01.01.1996 | USA |
| KC476743 | 01.01.2009 | South Africa |
| OR795314 | 06.01.2005 | Germany |

| | | |
|---|---|---|
| KF826848 | 01.01.2007 | Australia |
| JQ901452 | 22.12.2001 | Netherlands |
| JQ901447 | 03.01.2001 | Netherlands |
| JX661580 | 02.10.2004 | Germany |
| JX015486 | 05.01.2005 | Netherlands |
| OR795322 | 05.02.2007 | Germany |
| HQ731775 | 29.06.2007 | United Kingdom |
| KT765661 | 18.02.2000 | Kenya |
| KT765664 | 21.07.2000 | Kenya |
| KT765663 | 13.03.2000 | Kenya |
| MT854048 | 24.01.2013 | Zambia |
| JX131638 | 16.03.2008 | Saudi Arabia |
| JX182822 | 25.05.2007 | Brazil |
| JX182849 | 09.04.2010 | Brazil |
| KF826838 | 06.06.2006 | Argentina |
| JF920047 | 17.01.2008 | USA |
| JX513320 | 31.12.2007 | Brazil |
| JX015481 | 07.01.2009 | Netherlands |
| JX645814 | 10.12.2009 | Belgium |
| JX645812 | 11.12.2009 | Belgium |
| KU955051 | 08.04.2008 | China |
| KF300971 | 14.10.2008 | Panama |
| HQ731725 | 01.01.2010 | United Kingdom |
| JX645796 | 14.12.2008 | Belgium |
| JX015482 | 22.11.2006 | Belgium |
| JF714705 | 25.02.2008 | Saudi Arabia |
| JX661578 | 10.04.2008 | Germany |
| KF156409 | 09.12.2008 | Kenya |
| KU955056 | 31.12.2007 | China |
| KC342394 | 22.07.2010 | Thailand |
| HQ731730 | 01.01.2010 | United Kingdom |
| KM508818 | 04.07.2010 | Paraguay |
| KM508820 | 07.07.2011 | Paraguay |
| HQ731735 | 31.12.2009 | United Kingdom |
| KC297421 | 02.01.2009 | China |
| JF979153 | 11.03.2010 | Latvia |
| KR816586 | 11.12.2009 | Thailand |
| MK344683 | 21.09.2009 | Taiwan |
| KY327967 | 01.09.2012 | Thailand |

| | | |
|---|---|---|
| MK344715 | 31.12.2012 | Taiwan |
| KT765285 | 25.04.2011 | Kenya |
| KT765427 | 02.01.2011 | Kenya |
| KX510263 | 13.04.2010 | Kenya |
| KJ939949 | 26.03.2010 | Vietnam |
| JX976353 | 05.11.2011 | Cuba |
| MK109775 | 14.09.2011 | Jordan |
| KY327980 | 01.09.2012 | Thailand |
| JX661579 | 19.04.2008 | Germany |
| JX645831 | 13.12.2010 | Belgium |
| KC763335 | 03.10.2010 | Saudi Arabia |
| JX645830 | 18.01.2011 | Belgium |
| KC476658 | 15.04.2009 | South Africa |
| MN262219 | 14.03.2011 | Mozambique |
| KC476668 | 01.01.2010 | South Africa |
| KU955113 | 01.01.2007 | China |
| MN604236 | 01.03.2011 | Netherlands |
| JX645825 | 31.10.2008 | Belgium |
| KU955165 | 03.06.2007 | China |
| KU955083 | 01.01.2007 | China |
| KU955170 | 01.01.2011 | China |
| JX131639 | 16.03.2008 | Saudi Arabia |
| FJ157348 | 31.07.2005 | China |
| MK182714 | 01.03.2012 | Saudi Arabia |
| OR795330 | 24.04.2010 | Germany |
| MH760600 | 22.09.2011 | Australia |
| MH760593 | 17.06.2010 | Australia |
| JX513301 | 01.01.2011 | Brazil |
| KF915183 | 05.12.2011 | Spain |
| MG971423 | 29.01.2013 | Netherlands |
| MT853822 | 10.09.2013 | Mali |
| MT853882 | 05.07.2012 | South Africa |
| MN262215 | 16.12.2010 | Morocco |
| OR795335 | 08.02.2012 | Germany |
| MT853793 | 28.11.2012 | Mali |
| OR795333 | 10.01.2011 | Germany |
| JN257701 | 28.01.2011 | Canada |
| OQ261746 | 29.01.2019 | Austria |
| MK720784 | 01.01.2018 | Colombia |

| | | |
|---|---|---|
| MH181939 | 07.11.2012 | Kenya |
| ON237288 | 24.06.2016 | Argentina |
| MT853770 | 01.03.2013 | Gambia |
| MZ961918 | 31.01.2016 | China |
| KX453332 | 13.03.2013 | Kenya |
| KU350791 | 12.06.2012 | Argentina |
| KF915251 | 19.12.2012 | Spain |
| OR795341 | 18.01.2013 | Germany |
| KC677857 | 15.10.2012 | Cuba |
| KC677871 | 19.09.2012 | Cuba |
| MH142239 | 30.12.2016 | Russia |
| OR795347 | 18.03.2014 | Germany |
| KU350778 | 09.05.2012 | Argentina |
| MH760607 | 08.07.2013 | Australia |
| KY865207 | 01.01.2014 | Egypt |
| KU350811 | 13.06.2013 | Argentina |
| JN257694 | 19.01.2011 | Canada |
| JN257693 | 29.12.2010 | Canada |
| PP974146 | 08.01.2024 | China |
| MN031363 | 23.12.2016 | Cambodia |
| OP927977 | 13.03.2018 | Germany |
| OR288011 | 13.03.2019 | Zambia |
| MH750252 | 01.01.2015 | Brazil |
| KT781345 | 24.03.2014 | China |
| MG971424 | 14.02.2013 | Netherlands |
| OR162242 | 08.03.2022 | Kenya |
| OR795354 | 25.02.2016 | Germany |
| MF496477 | 01.01.2015 | Taiwan |
| MH388030 | 04.01.2016 | Saudi Arabia |
| PP529983 | 02.03.2020 | Hungary |
| MW678361 | 22.07.2019 | Thailand |
| KX765972 | 01.07.2013 | New Zealand |
| KX765919 | 09.06.2013 | New Zealand |
| KX765954 | 08.05.2013 | New Zealand |
| MF445799 | 21.11.2011 | China |
| KU681161 | 07.10.2013 | South Korea |
| MH828493 | 07.11.2013 | Vietnam |
| MG971433 | 03.10.2016 | Netherlands |
| MW160778 | 22.03.2017 | Australia |

| | | |
|---|---|---|
| MN434119 | 01.01.2017 | Saudi Arabia |
| MH828491 | 10.12.2013 | Vietnam |
| MH142232 | 25.03.2014 | Russia |
| MH142234 | 18.03.2013 | Russia |
| MF496378 | 25.10.2013 | Taiwan |
| PP525308 | 01.01.2017 | USA |
| MZ961932 | 30.11.2017 | China |
| OY757645 | 28.11.2022 | Australia |
| OP554443 | 01.05.2022 | Saudi Arabia |
| PP376653 | 18.12.2018 | Canada |
| PP958027 | 31.12.2017 | Ireland |
| MN007064 | 02.02.2018 | China |
| KY328023 | 08.10.2014 | Thailand |
| MK306343 | 01.01.2016 | Taiwan |
| KU681168 | 08.12.2014 | South Korea |
| PP352304 | 28.02.2018 | USA |
| MG971426 | 24.12.2014 | Netherlands |
| PP957980 | 01.01.2016 | Ireland |
| MN630100 | 01.02.2016 | USA |
| PP454564 | 01.01.2023 | Saudi Arabia |
| PP454566 | 01.01.2023 | Saudi Arabia |
| MH760610 | 19.11.2016 | Australia |
| MK306338 | 31.12.2016 | Taiwan |
| MW160761 | 31.03.2017 | Australia |
| MG793382 | 01.02.2015 | Lebanon |
| PP109421 | 30.10.2017 | United Kingdom |
| PQ349000 | 09.05.2022 | United Kingdom |
| PP969952 | 21.09.2022 | Ireland |
| OR287922 | 04.01.2020 | USA |
| OR522506 | 22.01.2023 | USA |
| PQ788245 | 17.08.2024 | USA |
| OR522487 | 28.11.2022 | USA |
| OR872596 | 14.10.2022 | USA |
| PP969998 | 15.11.2023 | Ireland |
| PP969999 | 20.11.2023 | Ireland |
| PP454567 | 31.12.2023 | Saudi Arabia |
| PQ066224 | 15.02.2024 | USA |
| PP270247 | 09.10.2023 | USA |
| OR522464 | 01.11.2022 | USA |

| | | |
|---|---|---|
| OP890337 | 30.11.2022 | USA |
| OR522510 | 15.01.2023 | USA |
| PQ762763 | 30.12.2023 | France |
| PP203259 | 31.12.2022 | USA |
| PQ638697 | 08.11.2022 | USA |
| OR522502 | 15.12.2022 | USA |
| OR522523 | 08.12.2022 | USA |
| PQ638681 | 25.12.2022 | USA |
| PP203263 | 31.12.2022 | USA |
| OP890316 | 01.10.2022 | USA |
| PQ638726 | 13.10.2022 | USA |
| PQ638680 | 15.12.2022 | USA |
| PQ638709 | 07.10.2022 | USA |
| PP770463 | 15.01.2024 | USA |
| PQ390738 | 31.12.2023 | USA |
| PQ390744 | 26.12.2023 | USA |
| PP342420 | 01.12.2023 | USA |
| PQ638788 | 15.11.2023 | USA |
| PQ638791 | 11.11.2023 | USA |
| PQ638792 | 03.11.2023 | USA |
| PQ638719 | 27.10.2023 | USA |
| PQ638714 | 17.10.2023 | USA |
| PQ638716 | 11.10.2023 | USA |
| PQ410479 | 31.12.2018 | Belgium |
| OQ171919 | 06.08.2022 | USA |
| PP681273 | 03.01.2023 | USA |
| OR601479 | 12.02.2023 | USA |
| OR975300 | 05.02.2023 | USA |
| OQ024137 | 06.11.2022 | USA |
| OQ024114 | 04.11.2022 | USA |
| OQ024116 | 03.11.2022 | USA |
| OQ171922 | 23.10.2022 | USA |
| OQ171920 | 17.10.2022 | USA |
| OQ024149 | 05.11.2022 | USA |
| PP352376 | 31.12.2022 | USA |
| OR975303 | 30.11.2022 | USA |
| OR915732 | 03.11.2022 | USA |
| OQ171916 | 23.10.2022 | USA |
| OQ171894 | 20.07.2022 | USA |

| | | |
|---|---|---|
| PP970055 | 31.10.2022 | Ireland |
| PP352342 | 30.11.2022 | USA |
| OQ171909 | 11.08.2022 | USA |
| PP352377 | 30.11.2022 | USA |
| OR601475 | 23.10.2022 | USA |
| OQ024120 | 03.11.2022 | USA |
| OQ171896 | 11.08.2022 | USA |
| OQ024139 | 04.11.2022 | USA |
| OQ171899 | 23.10.2022 | USA |
| MW026979 | 01.03.2018 | Brazil |
| OP927929 | 27.02.2020 | Germany |
| ON237343 | 29.06.2017 | Argentina |
| MT236290 | 29.06.2017 | Brazil |
| PP529974 | 28.12.2018 | Hungary |
| MZ515750 | 21.12.2019 | Spain |
| OR872589 | 31.10.2022 | USA |
| PQ834838 | 28.10.2022 | Mexico |
| PQ834839 | 09.11.2022 | Mexico |
| OR522503 | 18.12.2022 | USA |
| OR522501 | 18.12.2022 | USA |
| OR522518 | 04.12.2022 | USA |
| OR601477 | 19.12.2022 | USA |
| OR601476 | 17.12.2022 | USA |
| OR883002 | 25.05.2023 | USA |
| OR522521 | 11.12.2022 | USA |
| PP084056 | 10.12.2022 | USA |
| OR522499 | 18.12.2022 | USA |
| OR522504 | 26.12.2022 | USA |
| PP352326 | 01.10.2022 | USA |
| PP681267 | 13.12.2022 | USA |
| PP352375 | 30.11.2022 | USA |
| PP352350 | 31.10.2022 | USA |
| PP352356 | 01.10.2022 | USA |
| PQ618030 | 24.10.2022 | Panama |
| PQ618020 | 19.12.2022 | Panama |
| PQ618026 | 22.09.2022 | Panama |
| PQ618021 | 08.11.2022 | Panama |
| PQ618027 | 11.08.2022 | Panama |
| PQ618025 | 03.10.2022 | Panama |

| | | |
|---|---|---|
| PQ618022 | 08.11.2022 | Panama |
| PQ618023 | 27.10.2022 | Panama |
| PQ618029 | 10.10.2022 | Panama |
| PQ618028 | 10.10.2022 | Panama |
| PQ618024 | 20.10.2022 | Panama |
| PQ618032 | 20.10.2022 | Panama |
| PQ618031 | 01.01.2023 | Panama |
| PQ834849 | 22.12.2022 | Mexico |
| OR143142 | 12.12.2022 | USA |
| PQ834848 | 13.12.2022 | Mexico |
| OQ171901 | 18.10.2022 | USA |
| OR872628 | 21.09.2022 | USA |
| OR522491 | 06.12.2022 | USA |
| OR872640 | 09.11.2022 | USA |
| OR872622 | 27.04.2022 | USA |
| PQ638693 | 11.11.2022 | USA |
| PP352333 | 10.10.2022 | USA |
| PP352327 | 10.10.2022 | USA |
| OQ024109 | 08.11.2022 | USA |
| PP529981 | 17.01.2023 | Hungary |
| OR883007 | 27.11.2022 | USA |
| OQ024136 | 06.11.2022 | USA |
| PQ762819 | 09.11.2023 | France |
| PQ763132 | 27.12.2023 | France |
| PP352325 | 01.10.2022 | USA |
| PP352357 | 31.10.2022 | USA |
| PP681275 | 05.01.2023 | USA |
| OQ024121 | 05.11.2022 | USA |
| PP959043 | 04.01.2024 | USA |
| OR143222 | 30.11.2022 | USA |
| OR872602 | 26.10.2022 | USA |
| OR872625 | 15.10.2022 | USA |
| OR872632 | 18.08.2022 | USA |
| OR522507 | 23.01.2023 | USA |
| PQ638706 | 16.11.2022 | USA |
| OR601474 | 23.10.2022 | USA |
| PQ638704 | 16.11.2022 | USA |
| OR601472 | 16.10.2022 | USA |
| PQ638674 | 23.11.2022 | USA |

| | | |
|---|---|---|
| PQ638692 | 11.01.2023 | USA |
| PQ638730 | 15.10.2022 | USA |
| OR975305 | 22.12.2022 | USA |
| OR601473 | 16.10.2022 | USA |
| OR522462 | 30.10.2022 | USA |
| PQ638695 | 08.11.2022 | USA |
| OR522479 | 23.10.2022 | USA |
| PQ638668 | 25.11.2022 | USA |
| OR522520 | 30.01.2023 | USA |
| PQ638675 | 30.11.2022 | USA |
| PQ638688 | 11.11.2022 | USA |
| OR522512 | 10.01.2023 | USA |
| PP342434 | 17.12.2023 | USA |
| OR143189 | 29.01.2023 | USA |
| PP352351 | 01.10.2022 | USA |
| PP352368 | 01.11.2022 | USA |
| OR143184 | 20.02.2023 | USA |
| PP352352 | 01.10.2022 | USA |
| PP681243 | 06.12.2022 | USA |
| PP352354 | 31.10.2022 | USA |
| OR872644 | 24.10.2022 | USA |
| OR143154 | 09.12.2022 | USA |
| OR143168 | 02.12.2022 | USA |
| PP352341 | 01.11.2022 | USA |
| OR522517 | 05.12.2022 | USA |
| OR522498 | 18.10.2022 | USA |
| OR522497 | 18.10.2022 | USA |
| PP495848 | 09.01.2024 | USA |
| PP973778 | 16.02.2024 | USA |
| PP495851 | 15.01.2024 | USA |
| PP978499 | 16.02.2024 | USA |
| PQ849780 | 05.12.2024 | USA |
| PQ849777 | 06.12.2024 | USA |
| PQ849795 | 03.12.2024 | USA |
| PQ834845 | 25.10.2022 | Mexico |
| PQ834844 | 26.10.2022 | Mexico |
| PQ788250 | 14.01.2024 | USA |
| PP781407 | 27.01.2024 | USA |
| PQ834878 | 07.12.2022 | Mexico |

| | | |
|---|---|---|
| PQ834875 | 18.11.2022 | Mexico |
| PQ834869 | 16.11.2022 | Mexico |
| PQ834847 | 07.11.2022 | Mexico |
| OR872587 | 22.07.2022 | USA |
| OR143170 | 19.12.2022 | USA |
| OR143179 | 02.01.2023 | USA |
| OR872585 | 17.10.2022 | USA |
| PQ638673 | 25.11.2022 | USA |
| PP352369 | 01.11.2022 | USA |
| PQ066219 | 21.12.2023 | USA |
| PP352347 | 01.10.2022 | USA |
| PP594806 | 24.11.2023 | USA |
| PP973769 | 05.02.2024 | USA |
| PP910757 | 16.01.2024 | USA |
| PP910760 | 14.01.2024 | USA |
| PP973748 | 10.01.2024 | USA |
| PP973750 | 08.01.2024 | USA |
| PP709397 | 02.01.2024 | USA |
| PP709386 | 27.12.2023 | USA |
| PP709375 | 19.12.2023 | USA |
| PP709374 | 18.12.2023 | USA |
| PP973754 | 22.01.2024 | USA |
| PP910759 | 15.01.2024 | USA |
| PP973749 | 09.01.2024 | USA |
| PP934460 | 07.03.2024 | USA |
| PP973760 | 29.01.2024 | USA |
| PP973753 | 22.01.2024 | USA |
| PP973752 | 21.01.2024 | USA |
| PP973758 | 29.01.2024 | USA |
| PP973757 | 29.01.2024 | USA |
| PP352345 | 30.11.2022 | USA |
| OR143223 | 31.12.2022 | USA |
| OR143224 | 04.12.2022 | USA |
| PQ638703 | 22.03.2023 | USA |
| PP795161 | 06.01.2024 | USA |
| PP882667 | 21.11.2023 | USA |
| PP270250 | 13.11.2023 | USA |
| PP882659 | 17.10.2023 | USA |
| PP970062 | 27.10.2023 | Ireland |

| | | |
|---|---|---|
| PP970036 | 04.11.2023 | Ireland |
| PP969961 | 30.10.2023 | Ireland |
| PP970003 | 24.11.2023 | Ireland |
| PP970063 | 06.11.2023 | Ireland |
| PP970029 | 29.10.2023 | Ireland |
| PP970025 | 18.10.2023 | Ireland |
| PP970023 | 13.10.2023 | Ireland |
| PQ763136 | 17.01.2024 | France |
| PQ763005 | 29.12.2023 | France |
| PQ763126 | 24.11.2023 | France |
| OR883014 | 14.12.2022 | USA |
| PP781387 | 17.01.2024 | USA |
| PP934468 | 30.03.2024 | USA |
| PP934470 | 01.04.2024 | USA |
| PP957778 | 03.04.2024 | USA |
| PQ849789 | 28.11.2024 | USA |
| PP830408 | 21.12.2023 | USA |
| PP957771 | 28.03.2024 | USA |
| PP978527 | 03.03.2024 | USA |
| PQ763164 | 12.03.2024 | France |
| PQ762815 | 30.12.2023 | France |
| PQ762814 | 27.12.2023 | France |
| PQ763060 | 21.12.2023 | France |
| PQ763094 | 12.12.2023 | France |
| PQ763092 | 05.12.2023 | France |
| PQ763085 | 25.11.2023 | France |
| PQ763083 | 21.11.2023 | France |
| PQ763091 | 02.12.2023 | France |
| PQ763108 | 05.12.2023 | France |
| PQ762920 | 03.11.2023 | France |
| PQ763101 | 29.12.2023 | France |
| PQ763096 | 15.12.2023 | France |
| PQ763163 | 06.02.2024 | France |
| PQ763088 | 27.11.2023 | France |
| PQ763087 | 27.11.2023 | France |
| PQ763089 | 01.12.2023 | France |
| PQ763093 | 06.12.2023 | France |
| PQ763098 | 21.12.2023 | France |
| PQ763086 | 26.11.2023 | France |

| | | |
|---|---|---|
| PQ762810 | 27.11.2023 | France |
| PQ763084 | 23.11.2023 | France |
| PQ763097 | 18.12.2023 | France |
| PQ763090 | 01.12.2023 | France |
| PQ762843 | 08.01.2024 | France |
| PQ762986 | 29.12.2023 | France |
| PQ763011 | 27.11.2023 | France |
| PQ763010 | 26.12.2023 | France |
| PQ762981 | 24.12.2023 | France |
| PQ763000 | 22.12.2023 | France |
| PQ762996 | 11.02.2024 | France |
| PQ763001 | 25.11.2023 | France |
| PQ763059 | 20.12.2023 | France |
| PQ762653 | 19.12.2023 | France |
| PQ763012 | 28.11.2023 | France |
| PQ762997 | 14.12.2023 | France |
| PQ763006 | 20.12.2023 | France |
| PQ763017 | 17.12.2023 | France |
| PQ762706 | 17.12.2023 | France |
| PQ763003 | 05.01.2024 | France |
| PQ762982 | 13.12.2023 | France |
| PP781409 | 23.01.2024 | USA |
| PP847361 | 09.01.2024 | USA |
| PP781397 | 29.01.2024 | USA |
| PP278015 | 23.10.2023 | USA |
| PQ066216 | 01.01.2024 | USA |
| PQ066222 | 19.12.2023 | USA |
| PP795137 | 19.10.2023 | USA |
| PP847379 | 11.01.2024 | USA |
| PP668165 | 03.12.2023 | USA |
| PP386340 | 27.10.2023 | USA |
| PP135017 | 07.09.2023 | USA |
| PP770476 | 20.02.2024 | USA |
| PQ788206 | 25.11.2024 | USA |
| PQ788208 | 22.11.2024 | USA |
| PP847386 | 21.01.2024 | USA |
| PP709445 | 28.12.2023 | USA |
| PP386335 | 26.10.2023 | USA |
| PP504644 | 13.11.2023 | USA |

| | | |
|---|---|---|
| PP847370 | 04.01.2024 | USA |
| PP830389 | 16.12.2023 | USA |
| PP830395 | 14.12.2023 | USA |
| PP957776 | 06.04.2024 | USA |
| PP957772 | 30.03.2024 | USA |
| PQ762507 | 19.03.2024 | USA |
| PQ762505 | 18.03.2024 | USA |
| PP978523 | 26.02.2024 | USA |
| PP910783 | 12.02.2024 | USA |
| PP910776 | 29.01.2024 | USA |
| PP847387 | 18.01.2024 | USA |
| PP830410 | 23.12.2023 | USA |
| PP270262 | 04.12.2023 | USA |
| PP795149 | 15.11.2023 | USA |
| PP795142 | 28.10.2023 | USA |
| PP795140 | 26.10.2023 | USA |
| PP795136 | 16.10.2023 | USA |
| PP795160 | 02.01.2024 | USA |
| PQ762522 | 02.05.2024 | USA |
| PP957765 | 21.03.2024 | USA |
| PP978535 | 11.03.2024 | USA |
| PP978526 | 02.03.2024 | USA |
| PQ762517 | 28.04.2024 | USA |
| PP795145 | 01.11.2023 | USA |
| PP934452 | 02.03.2024 | USA |
| PP934456 | 05.03.2024 | USA |
| PP934472 | 02.04.2024 | USA |
| PP934469 | 01.04.2024 | USA |
| PP830398 | 17.12.2023 | USA |
| PP978529 | 05.03.2024 | USA |
| PP847393 | 18.01.2024 | USA |
| PP594796 | 17.11.2023 | USA |
| PP973747 | 12.01.2024 | USA |
| PP871355 | 03.10.2023 | USA |
| PP957760 | 07.02.2024 | USA |
| PQ762508 | 19.03.2024 | USA |
| PP847369 | 09.01.2024 | USA |
| PP957767 | 25.03.2024 | USA |
| PP934473 | 13.04.2024 | USA |

| | | |
|---|---|---|
| PP934474 | 13.04.2024 | USA |
| PP978498 | 16.02.2024 | USA |
| PP978525 | 02.03.2024 | USA |
| PP910771 | 27.01.2024 | USA |
| OQ261751 | 06.05.2022 | Austria |
| PP781389 | 14.02.2024 | USA |
| PP969942 | 28.12.2022 | Ireland |
| OR162259 | 13.04.2022 | Kenya |
| OR162265 | 19.05.2022 | Kenya |
| OR162267 | 24.05.2022 | Kenya |
| OR162247 | 10.03.2022 | Kenya |
| OR162238 | 02.03.2022 | Kenya |
| OR162234 | 01.03.2022 | Kenya |
| OR162253 | 19.03.2022 | Kenya |
| PQ763129 | 11.12.2023 | France |
| PQ763128 | 05.12.2023 | France |
| OR287842 | 29.10.2019 | USA |
| PQ762803 | 11.12.2023 | France |
| PQ117646 | 04.06.2024 | USA |
| PQ763074 | 06.01.2024 | France |
| OR162266 | 22.05.2022 | Kenya |
| PP830378 | 04.12.2023 | USA |
| OR162255 | 25.03.2022 | Kenya |
| OR162261 | 19.04.2022 | Kenya |
| OR162256 | 28.03.2022 | Kenya |
| OR162260 | 13.04.2022 | Kenya |
| OR162268 | 26.05.2022 | Kenya |
| OR162271 | 22.06.2022 | Kenya |
| OR162257 | 30.03.2022 | Kenya |
| OR162250 | 12.03.2022 | Kenya |
| OR162235 | 02.03.2022 | Kenya |
| OR162258 | 10.04.2022 | Kenya |
| OR162249 | 11.03.2022 | Kenya |
| OR143206 | 07.12.2022 | USA |
| PQ349026 | 13.04.2023 | United Kingdom |
| PQ763133 | 02.01.2024 | France |
| PQ762764 | 03.01.2024 | France |
| PQ763106 | 02.12.2023 | France |
| PQ762656 | 24.12.2023 | France |

| | | |
|---|---|---|
| PP970021 | 02.10.2023 | Ireland |
| PQ762659 | 12.01.2024 | France |
| PQ762851 | 05.12.2023 | France |
| PQ762725 | 21.11.2023 | France |
| PP970004 | 24.11.2023 | Ireland |
| PQ763161 | 12.12.2023 | France |
| PQ762752 | 15.11.2023 | France |
| OM857287 | 29.11.2020 | Australia |
| PP530274 | 08.12.2023 | USA |
| PQ762995 | 20.01.2024 | France |
| PQ763077 | 26.12.2023 | France |
| PQ763075 | 28.12.2023 | France |
| PQ763141 | 19.01.2024 | France |
| PQ762701 | 07.12.2023 | France |
| PQ762993 | 12.11.2023 | France |
| PQ763049 | 16.12.2023 | France |
| PQ410468 | 13.11.2022 | Belgium |
| PQ376587 | 25.10.2023 | Belgium |
| OR872610 | 07.10.2022 | USA |
| PQ763052 | 24.11.2023 | France |
| PQ763144 | 11.12.2023 | France |
| PQ762719 | 13.12.2023 | France |
| PQ763032 | 04.02.2024 | France |
| PQ762731 | 06.12.2023 | France |
| PQ762671 | 28.11.2023 | France |
| PQ763051 | 26.11.2023 | France |
| PQ762722 | 30.10.2023 | France |
| PQ762745 | 18.10.2023 | France |
| PQ763110 | 08.12.2023 | France |
| PQ763115 | 14.12.2023 | France |
| PQ763113 | 11.12.2023 | France |
| PQ763118 | 26.12.2023 | France |
| PQ763151 | 10.12.2023 | France |
| PQ762902 | 02.01.2024 | France |
| PQ763042 | 12.12.2023 | France |
| PQ763165 | 12.12.2023 | France |
| PQ762771 | 31.12.2023 | France |
| PQ762712 | 24.12.2023 | France |
| PQ762947 | 10.12.2023 | France |

| | | |
|---|---|---|
| PQ762944 | 08.12.2023 | France |
| PQ762931 | 04.12.2023 | France |
| PQ763080 | 04.12.2023 | France |
| PQ762922 | 18.11.2023 | France |
| PQ762928 | 28.11.2023 | France |
| PQ762932 | 06.12.2023 | France |
| PQ762924 | 21.11.2023 | France |
| PQ763160 | 30.11.2023 | France |
| PQ762943 | 07.12.2023 | France |
| PQ763008 | 26.12.2023 | France |
| PQ763099 | 25.12.2023 | France |
| PQ762934 | 11.12.2023 | France |
| PQ762919 | 01.11.2023 | France |
| PQ762936 | 28.11.2023 | France |
| PQ762933 | 09.12.2023 | France |
| PQ762939 | 04.12.2023 | France |
| PQ762941 | 05.12.2023 | France |
| PQ762935 | 12.12.2023 | France |
| PQ762923 | 19.11.2023 | France |
| PQ763111 | 19.12.2023 | France |
| PQ763079 | 21.12.2023 | France |
| PQ762809 | 09.12.2023 | France |
| PQ762794 | 28.11.2023 | France |
| PQ763078 | 13.11.2023 | France |
| PQ762795 | 08.11.2023 | France |
| PQ762693 | 21.11.2023 | France |
| PQ762930 | 02.12.2023 | France |
| PQ762925 | 24.11.2023 | France |
| PQ762921 | 08.11.2023 | France |
| PQ762899 | 09.01.2024 | France |
| PQ763138 | 04.12.2023 | France |
| PQ762696 | 05.12.2023 | France |
| PQ762945 | 10.12.2023 | France |
| PQ762950 | 03.12.2023 | France |
| PQ762948 | 11.12.2023 | France |
| PQ762927 | 26.11.2023 | France |
| PQ763114 | 13.12.2023 | France |
| PQ763015 | 26.03.2024 | France |
| PQ763058 | 20.12.2023 | France |

| | | |
|---|---|---|
| PQ762988 | 25.12.2023 | France |
| PQ762709 | 19.12.2023 | France |
| PQ762697 | 06.12.2023 | France |
| PQ763162 | 11.01.2024 | France |
| OR162251 | 14.03.2022 | Kenya |
| OR162246 | 10.03.2022 | Kenya |
| OR162252 | 16.03.2022 | Kenya |
| OR162237 | 01.03.2022 | Kenya |
| OR162269 | 04.06.2022 | Kenya |
| OR162278 | 07.09.2022 | Kenya |
| OR162233 | 28.02.2022 | Kenya |
| OR162272 | 23.06.2022 | Kenya |
| OR162232 | 27.02.2022 | Kenya |
| OR162245 | 09.03.2022 | Kenya |
| PP970034 | 04.11.2023 | Ireland |
| PP969949 | 18.07.2022 | Ireland |
| PP969941 | 03.12.2022 | Ireland |
| PP969982 | 08.12.2022 | Ireland |
| PP970013 | 30.11.2022 | Ireland |
| PP969995 | 13.12.2022 | Ireland |
| PP970014 | 19.12.2022 | Ireland |
| PP352361 | 30.11.2022 | USA |
| PP352362 | 01.11.2022 | USA |
| PP352330 | 01.10.2022 | USA |
| PP352340 | 01.11.2022 | USA |
| PP970020 | 04.11.2023 | Ireland |
| PP978522 | 26.02.2024 | USA |
| PQ788199 | 26.11.2024 | USA |
| PP969988 | 25.11.2023 | Ireland |
| PQ788216 | 19.11.2024 | USA |
| PP770469 | 27.02.2024 | USA |
| PQ066220 | 21.12.2023 | USA |
| PQ348860 | 29.07.2022 | United Kingdom |
| PP969997 | 14.11.2023 | Ireland |
| PQ349023 | 06.02.2023 | United Kingdom |
| PQ348886 | 05.11.2022 | United Kingdom |
| PQ763120 | 04.01.2024 | France |
| PQ763117 | 24.12.2023 | France |
| PQ763107 | 04.12.2023 | France |

| | | |
|---|---|---|
| PQ763135 | 05.01.2024 | France |
| PP781416 | 14.12.2023 | USA |
| PP970017 | 01.11.2023 | Ireland |
| PP970042 | 19.11.2023 | Ireland |
| PQ762807 | 03.12.2023 | France |
| PQ762624 | 08.11.2023 | France |
| PQ737347 | 08.11.2023 | France |
| PQ762834 | 24.11.2023 | France |
| PQ762870 | 23.12.2023 | France |
| PP781415 | 17.02.2024 | USA |
| PQ762979 | 21.12.2023 | France |
| PQ763140 | 18.01.2024 | France |
| PQ762787 | 18.12.2023 | France |
| PQ762946 | 10.12.2023 | France |
| PQ763076 | 04.01.2024 | France |
| PQ763041 | 03.12.2023 | France |
| PQ763007 | 31.03.2024 | France |
| PQ763149 | 24.12.2023 | France |
| PQ763014 | 22.12.2023 | France |
| PQ763130 | 18.12.2023 | France |
| PQ762910 | 18.12.2023 | France |
| PQ763150 | 15.12.2023 | France |
| PQ763153 | 12.12.2023 | France |
| PQ763152 | 10.12.2023 | France |
| PQ763127 | 02.12.2023 | France |
| PQ762994 | 28.11.2023 | France |
| PQ762980 | 22.11.2023 | France |
| PQ762820 | 09.11.2023 | France |
| PQ762816 | 08.11.2023 | France |
| PQ762655 | 24.12.2023 | France |
| PQ762866 | 24.11.2023 | France |
| PQ762718 | 12.12.2023 | France |
| PQ763123 | 23.12.2023 | France |
| PQ762742 | 11.12.2023 | France |
| PQ762677 | 31.10.2023 | France |
| PQ762644 | 08.12.2023 | France |
| PQ737367 | 08.12.2023 | France |
| PQ762914 | 23.11.2023 | France |
| PQ763156 | 13.11.2023 | France |

| | | |
|---|---|---|
| PQ762664 | 28.11.2023 | France |
| PQ762758 | 13.12.2023 | France |
| PQ763116 | 18.12.2023 | France |
| PQ762858 | 16.12.2023 | France |
| PQ763057 | 05.12.2023 | France |
| PQ762913 | 17.11.2023 | France |
| PQ763050 | 17.11.2023 | France |
| PQ763104 | 23.10.2023 | France |
| PQ763054 | 23.12.2023 | France |
| PQ762859 | 16.12.2023 | France |
| PQ762918 | 23.11.2023 | France |
| PQ762833 | 12.12.2023 | France |
| PQ763105 | 05.12.2023 | France |
| PQ762825 | 02.11.2023 | France |
| PQ762873 | 19.12.2023 | France |
| PQ763124 | 31.12.2023 | France |
| PQ763157 | 21.11.2023 | France |
| PQ763037 | 03.12.2023 | France |
| PQ762668 | 17.12.2023 | France |
| PQ763145 | 25.12.2023 | France |
| PQ849798 | 28.11.2024 | USA |
| PQ762883 | 08.01.2024 | France |
| PQ763102 | 04.01.2024 | France |
| PP959045 | 04.01.2024 | USA |
| PQ762658 | 03.01.2024 | France |
| PQ763002 | 27.12.2023 | France |
| PQ762911 | 27.12.2023 | France |
| PQ763016 | 27.12.2023 | France |
| PQ762813 | 25.12.2023 | France |
| PQ762942 | 06.12.2023 | France |
| PQ737366 | 06.12.2023 | France |
| PQ762872 | 26.11.2023 | France |
| PQ762620 | 02.11.2023 | France |
| PQ737353 | 30.10.2023 | France |
| PQ762892 | 25.10.2023 | France |
| PQ376586 | 19.10.2023 | Belgium |
| PQ763056 | 17.12.2023 | France |
| PQ737343 | 02.11.2023 | France |
| PQ763061 | 12.01.2024 | France |

| | | |
|---|---|---|
| PQ762663 | 26.12.2023 | France |
| PQ763109 | 14.12.2023 | France |
| PQ762828 | 17.12.2023 | France |
| PQ763095 | 12.12.2023 | France |
| PQ762630 | 30.10.2023 | France |
| PQ762643 | 06.12.2023 | France |
| PQ763009 | 27.12.2023 | France |
| PQ762735 | 06.12.2023 | France |
| PQ762740 | 30.11.2023 | France |
| PQ762733 | 27.11.2023 | France |
| PQ762730 | 26.11.2023 | France |
| PQ763069 | 02.01.2024 | France |
| PQ763070 | 13.12.2023 | France |
| PQ737345 | 29.10.2023 | France |
| PQ762622 | 29.10.2023 | France |
| PQ618042 | 01.12.2022 | Panama |
| PP270276 | 30.11.2023 | USA |
| PP270275 | 04.12.2023 | USA |
| PP495846 | 03.01.2024 | USA |
| PP957756 | 07.02.2024 | USA |
| OR143138 | 19.12.2022 | USA |
| OR143220 | 16.12.2022 | USA |
| OR882975 | 17.11.2022 | USA |
| OR872645 | 25.10.2022 | USA |
| OR872626 | 26.10.2022 | USA |
| OR143197 | 30.11.2022 | USA |
| OR882985 | 07.11.2022 | USA |
| OR882981 | 04.11.2022 | USA |
| OR915734 | 03.11.2022 | USA |
| OR915756 | 24.10.2022 | USA |
| PP903812 | 22.10.2022 | USA |
| OR882974 | 26.11.2022 | USA |
| OR882978 | 29.11.2022 | USA |
| PP970035 | 04.11.2023 | Ireland |
| PP970032 | 31.10.2023 | Ireland |
| MZ515640 | 31.12.2019 | Spain |
| PQ618033 | 15.01.2020 | Panama |
| OR915735 | 19.01.2023 | USA |
| OQ024155 | 04.11.2022 | USA |

| | | |
|---|---|---|
| PP681283 | 05.12.2022 | USA |
| PP681242 | 06.12.2022 | USA |
| OR143137 | 13.12.2022 | USA |
| OR522496 | 17.10.2022 | USA |
| OR143188 | 25.12.2022 | USA |
| PP760419 | 15.10.2022 | USA |
| OP890314 | 15.10.2022 | USA |
| PP352344 | 30.11.2022 | USA |
| OY757638 | 22.07.2022 | Australia |
| OY757684 | 22.07.2022 | Australia |
| OY757641 | 15.07.2022 | Australia |
| OY757595 | 15.07.2022 | Australia |
| PP770472 | 19.02.2024 | USA |
| OR143209 | 19.12.2022 | USA |
| PP969963 | 04.12.2023 | Ireland |
| PP969991 | 29.11.2023 | Ireland |
| PP969989 | 26.11.2023 | Ireland |
| PP970018 | 02.11.2023 | Ireland |
| PP970019 | 01.11.2023 | Ireland |
| PP970026 | 20.10.2023 | Ireland |
| PP970022 | 11.10.2023 | Ireland |
| PP970033 | 26.10.2023 | Ireland |
| PP969957 | 24.11.2023 | Ireland |
| PP970028 | 28.10.2023 | Ireland |
| PP970044 | 15.11.2023 | Ireland |
| PP970037 | 06.11.2023 | Ireland |
| PP970061 | 19.10.2023 | Ireland |
| PP970046 | 19.11.2023 | Ireland |
| PP969962 | 04.12.2023 | Ireland |
| PP970041 | 19.11.2023 | Ireland |
| PP969985 | 16.11.2023 | Ireland |
| PP969984 | 16.11.2023 | Ireland |
| PP970005 | 17.11.2023 | Ireland |
| PP969958 | 23.11.2023 | Ireland |
| PP970000 | 17.11.2023 | Ireland |
| PP969986 | 27.11.2023 | Ireland |
| PP969959 | 15.11.2023 | Ireland |
| PP969956 | 22.11.2023 | Ireland |
| PQ849787 | 03.12.2024 | USA |

| | | |
|---|---|---|
| PQ762801 | 20.11.2023 | France |
| PQ349019 | 27.09.2022 | United Kingdom |
| PQ348960 | 27.09.2022 | United Kingdom |
| PQ348963 | 27.09.2022 | United Kingdom |
| PQ348962 | 27.09.2022 | United Kingdom |
| PQ763064 | 20.12.2023 | France |
| PQ737349 | 05.12.2023 | France |
| PQ762626 | 05.12.2023 | France |
| PQ762769 | 19.12.2023 | France |
| PQ763112 | 22.12.2023 | France |
| PQ762726 | 24.11.2023 | France |
| PQ763071 | 06.01.2024 | France |
| PQ762721 | 31.10.2023 | France |
| PQ762749 | 30.10.2023 | France |
| OR522473 | 06.11.2022 | USA |
| OR872616 | 28.09.2022 | USA |
| PP910758 | 16.01.2024 | USA |
| PP352338 | 01.10.2022 | USA |
| PQ618041 | 20.09.2022 | Panama |
| PQ618040 | 31.10.2022 | Panama |
| OQ171931 | 01.07.2022 | USA |
| OQ171911 | 13.07.2022 | USA |
| OQ024154 | 14.11.2022 | USA |
| OQ171926 | 26.10.2022 | USA |
| OR143183 | 01.12.2022 | USA |
| OR143198 | 06.12.2022 | USA |
| OQ171897 | 23.10.2022 | USA |
| OQ024152 | 03.11.2022 | USA |
| PQ762661 | 15.01.2024 | France |
| PQ762634 | 13.11.2023 | France |
| PQ737357 | 13.11.2023 | France |
| PP681280 | 10.01.2023 | USA |
| PP203260 | 12.10.2022 | USA |
| PP681260 | 13.12.2022 | USA |
| PQ638672 | 24.11.2022 | USA |
| PP681270 | 20.12.2022 | USA |
| OR522526 | 30.01.2023 | USA |
| OR522527 | 01.02.2023 | USA |
| OQ024124 | 14.11.2022 | USA |

| | | |
|---|---|---|
| OQ024148 | 03.11.2022 | USA |
| PQ618039 | 23.12.2023 | Panama |
| OR601470 | 02.10.2022 | USA |
| OR872619 | 21.06.2022 | USA |
| PP903821 | 21.10.2022 | USA |
| PP903827 | 21.10.2022 | USA |
| PP970008 | 17.11.2022 | Ireland |
| PP970054 | 30.10.2022 | Ireland |
| PP970052 | 19.10.2022 | Ireland |
| PP970051 | 17.10.2022 | Ireland |
| PQ348882 | 16.10.2022 | United Kingdom |
| PQ348881 | 16.10.2022 | United Kingdom |
| PQ348877 | 08.10.2022 | United Kingdom |
| PQ348884 | 19.10.2022 | United Kingdom |
| PQ348883 | 17.10.2022 | United Kingdom |
| PQ348875 | 03.10.2022 | United Kingdom |
| PQ348862 | 27.08.2022 | United Kingdom |
| PQ349022 | 21.11.2022 | United Kingdom |
| PQ348869 | 09.09.2022 | United Kingdom |
| PQ348861 | 28.08.2022 | United Kingdom |
| PQ348880 | 13.10.2022 | United Kingdom |
| PQ348868 | 09.09.2022 | United Kingdom |
| PQ348865 | 28.08.2022 | United Kingdom |
| PQ348876 | 04.10.2022 | United Kingdom |
| PQ348864 | 14.08.2022 | United Kingdom |
| PQ348859 | 26.07.2022 | United Kingdom |
| PP970058 | 08.12.2022 | Ireland |
| PQ349008 | 19.07.2022 | United Kingdom |
| PQ349014 | 08.08.2022 | United Kingdom |
| PQ349013 | 04.08.2022 | United Kingdom |
| PQ349012 | 01.08.2022 | United Kingdom |
| PQ349009 | 22.07.2022 | United Kingdom |
| PQ349006 | 13.07.2022 | United Kingdom |
| PQ348967 | 18.03.2022 | United Kingdom |
| PQ348965 | 18.03.2022 | United Kingdom |
| PQ348999 | 18.03.2022 | United Kingdom |
| PQ349015 | 09.08.2022 | United Kingdom |
| PQ348879 | 12.10.2022 | United Kingdom |
| PQ348961 | 11.07.2022 | United Kingdom |

| | | |
|---|---|---|
| PQ349004 | 11.07.2022 | United Kingdom |
| PP970056 | 03.11.2022 | Ireland |
| PP970049 | 11.10.2022 | Ireland |
| PP970048 | 11.10.2022 | Ireland |
| PQ349007 | 16.07.2022 | United Kingdom |
| PQ349018 | 18.09.2022 | United Kingdom |
| PQ349001 | 17.05.2022 | United Kingdom |
| PP969950 | 10.09.2022 | Ireland |
| PQ349005 | 13.07.2022 | United Kingdom |
| PQ349017 | 05.09.2022 | United Kingdom |
| PQ349010 | 26.07.2022 | United Kingdom |
| PP781403 | 02.02.2024 | USA |
| PP270232 | 24.10.2023 | USA |
| PP270274 | 06.12.2023 | USA |
| PP781402 | 28.12.2023 | USA |
| PP781388 | 05.12.2023 | USA |
| PP903809 | 16.03.2024 | USA |
| PP270251 | 23.10.2023 | USA |
| PP781418 | 05.02.2024 | USA |
| PP270271 | 28.11.2023 | USA |
| OR882986 | 16.05.2023 | USA |
| PP270253 | 02.10.2023 | USA |
| PP781417 | 14.12.2023 | USA |
| PP781420 | 18.12.2023 | USA |
| PP270272 | 12.12.2023 | USA |
| PP270231 | 21.11.2023 | USA |
| PP781383 | 15.01.2024 | USA |
| PP270270 | 02.12.2023 | USA |
| PP795159 | 03.01.2024 | USA |
| PP882666 | 14.11.2023 | USA |
| PP386365 | 05.11.2023 | USA |
| PP795133 | 05.10.2023 | USA |
| PP882665 | 08.11.2023 | USA |
| PQ762761 | 15.12.2023 | France |
| PQ762897 | 02.11.2023 | France |
| PQ762882 | 03.12.2023 | France |
| OQ171906 | 27.07.2022 | USA |
| OR522529 | 01.01.2023 | USA |
| OR143187 | 26.12.2022 | USA |

| | | |
|---|---|---|
| PP530267 | 29.11.2023 | USA |
| PQ610207 | 11.06.2023 | Argentina |
| PQ390732 | 04.01.2024 | USA |
| PP882664 | 02.11.2023 | USA |
| PQ788217 | 20.11.2024 | USA |
| PQ849796 | 28.11.2024 | USA |
| PQ788215 | 13.11.2024 | USA |
| OR143171 | 01.02.2023 | USA |
| OR143166 | 01.02.2023 | USA |
| PQ610205 | 15.06.2023 | Argentina |
| PQ610204 | 04.06.2023 | Argentina |
| PQ610203 | 23.05.2023 | Argentina |
| PQ762938 | 02.12.2023 | France |
| PQ610209 | 15.05.2023 | Argentina |
| PP934463 | 17.03.2024 | USA |
| PQ762759 | 14.12.2023 | France |
| PQ763158 | 24.11.2023 | France |
| PQ610198 | 22.05.2023 | Argentina |
| PQ610200 | 29.05.2023 | Argentina |
| PP934471 | 02.04.2024 | USA |
| PP957750 | 03.02.2024 | USA |
| PQ762509 | 19.03.2024 | USA |
| PQ762523 | 04.05.2024 | USA |
| PQ610199 | 27.05.2023 | Argentina |
| PQ610197 | 12.05.2023 | Argentina |
| PQ610196 | 25.05.2023 | Argentina |
| PQ610202 | 11.05.2023 | Argentina |
| PQ610201 | 17.05.2023 | Argentina |
| PP790963 | 04.12.2023 | Mexico |
| PQ610208 | 23.06.2023 | Argentina |
| PQ610206 | 22.05.2023 | Argentina |
| PQ834887 | 21.11.2023 | Mexico |
| PP495853 | 31.01.2024 | USA |
| PP770471 | 11.01.2024 | USA |
| PQ834895 | 04.12.2023 | Mexico |
| PQ834890 | 27.11.2023 | Mexico |
| PQ834883 | 14.11.2023 | Mexico |
| PQ834879 | 16.10.2023 | Mexico |
| PQ834901 | 13.12.2023 | Mexico |

| | | |
|---|---|---|
| PQ834899 | 06.12.2023 | Mexico |
| PQ762876 | 28.11.2023 | France |
| PQ762875 | 28.11.2023 | France |
| PP969992 | 29.11.2023 | Ireland |
| PP978501 | 21.02.2024 | USA |
| PP978536 | 11.03.2024 | USA |
| PP978534 | 10.03.2024 | USA |
| PP795148 | 10.11.2023 | USA |
| PQ618038 | 13.10.2023 | Panama |
| PQ610195 | 03.07.2024 | Argentina |
| PQ117649 | 02.01.2024 | USA |
| PP668172 | 05.12.2023 | USA |
| PP709446 | 29.12.2023 | USA |
| PP781425 | 02.01.2024 | USA |
| PP342431 | 13.12.2023 | USA |
| PQ834857 | 28.11.2022 | Mexico |
| PQ834872 | 18.11.2022 | Mexico |
| PQ638684 | 31.12.2022 | USA |
| PQ638670 | 22.11.2022 | USA |
| PQ638676 | 12.11.2022 | USA |
| PP781390 | 29.02.2024 | USA |
| OR872588 | 23.10.2022 | USA |
| OR872637 | 16.10.2022 | USA |
| OR872607 | 25.09.2022 | USA |
| OR975306 | 29.09.2023 | USA |
| PQ638682 | 23.12.2022 | USA |
| PP795156 | 14.12.2023 | USA |
| PP795162 | 19.01.2024 | USA |
| PP795157 | 22.12.2023 | USA |
| PQ834855 | 04.11.2022 | Mexico |
| PQ834858 | 28.11.2022 | Mexico |
| PQ834870 | 16.11.2022 | Mexico |
| PQ834861 | 11.01.2023 | Mexico |
| PQ834907 | 16.01.2024 | Mexico |
| PQ834908 | 24.01.2024 | Mexico |
| PP342419 | 30.11.2023 | USA |
| PP342426 | 05.12.2023 | USA |
| PQ008881 | 11.12.2023 | USA |
| OR872618 | 13.09.2022 | USA |

| | | |
|---|---|---|
| OR883018 | 07.11.2022 | USA |
| OR882991 | 01.11.2022 | USA |
| OR915770 | 15.11.2022 | USA |
| PP781426 | 23.02.2024 | USA |
| PP270243 | 01.11.2023 | USA |
| PP270265 | 06.12.2023 | USA |
| PP781378 | 03.01.2024 | USA |
| PP781406 | 22.12.2023 | USA |
| PP781385 | 30.01.2024 | USA |
| PQ834866 | 27.02.2023 | Mexico |
| PQ834860 | 02.12.2022 | Mexico |
| PQ834868 | 15.11.2022 | Mexico |
| PP084057 | 25.10.2023 | USA |
| PP352384 | 01.12.2022 | USA |
| PQ834862 | 16.01.2023 | Mexico |
| PQ638696 | 11.11.2022 | USA |
| PQ763004 | 04.10.2023 | France |
| PQ762987 | 28.11.2023 | France |
| PQ763013 | 26.12.2023 | France |
| PQ762896 | 05.12.2023 | France |
| PQ762901 | 04.12.2023 | France |
| PQ762887 | 12.12.2023 | France |
| PQ762908 | 01.12.2023 | France |
| PQ834897 | 05.12.2023 | Mexico |
| PQ834900 | 08.12.2023 | Mexico |
| PQ834905 | 14.12.2023 | Mexico |
| PQ638722 | 29.01.2023 | USA |
| PQ834876 | 01.12.2022 | Mexico |
| PQ834854 | 01.11.2022 | Mexico |
| OR975308 | 24.10.2023 | USA |
| PP342441 | 04.01.2024 | USA |
| PP084058 | 02.11.2023 | USA |
| PP970024 | 13.10.2023 | Ireland |
| PP342416 | 19.11.2023 | USA |
| PP970040 | 20.11.2023 | Ireland |
| OR975307 | 22.10.2023 | USA |
| PP342435 | 17.12.2023 | USA |
| PQ008877 | 01.01.2024 | USA |
| PP342438 | 20.12.2023 | USA |

| | | |
|---|---|---|
| PP342422 | 04.12.2023 | USA |
| PQ834863 | 24.01.2023 | Mexico |
| PQ834864 | 24.01.2023 | Mexico |
| PP852062 | 13.02.2024 | USA |
| PP910768 | 26.01.2024 | USA |
| PP847391 | 17.01.2024 | USA |
| PP978537 | 12.03.2024 | USA |
| PQ762526 | 19.06.2024 | USA |
| OR872583 | 01.10.2022 | USA |
| PQ834865 | 15.02.2023 | Mexico |
| PQ834874 | 18.11.2022 | Mexico |
| PQ834871 | 16.11.2022 | Mexico |
| PP352349 | 01.10.2022 | USA |
| PQ834867 | 14.11.2022 | Mexico |
| PP978496 | 31.12.2024 | USA |
| PP342439 | 20.12.2023 | USA |
| PP969987 | 27.11.2023 | Ireland |
| PQ849801 | 20.09.2024 | USA |
| PQ762916 | 31.12.2023 | France |
| PQ638678 | 01.12.2022 | USA |
| PQ762799 | 15.12.2023 | France |
| PQ762798 | 15.12.2023 | France |
| PQ763155 | 12.11.2023 | France |
| MW678370 | 18.09.2019 | Thailand |
| MW678310 | 29.10.2018 | Thailand |
| PP151402 | 16.09.2022 | Kuwait |
| PP151344 | 19.10.2021 | Kuwait |
| PP151368 | 11.05.2022 | Kuwait |
| PP957764 | 22.03.2024 | USA |
| PP709396 | 04.01.2024 | USA |
| OR840708 | 15.04.2023 | China |
| OR840704 | 05.04.2023 | China |
| PP974155 | 10.12.2023 | China |
| PP270249 | 03.11.2023 | USA |
| PP903814 | 19.03.2024 | USA |
| PQ638797 | 08.11.2023 | USA |
| PP760389 | 01.10.2022 | USA |
| OR883017 | 13.04.2023 | USA |
| OQ171910 | 14.10.2022 | USA |

| | | |
|---|---|---|
| PP508181 | 30.05.2022 | Peru |
| OR522477 | 10.11.2022 | USA |
| OR872621 | 16.09.2022 | USA |
| PP352346 | 31.10.2022 | USA |
| PP135031 | 22.09.2023 | USA |
| PP342432 | 14.12.2023 | USA |
| OR143140 | 19.12.2022 | USA |
| PQ834856 | 16.11.2022 | Mexico |
| PP781413 | 31.01.2024 | USA |
| PQ618051 | 07.12.2022 | Panama |
| PQ390730 | 29.12.2023 | USA |
| PQ762990 | 29.11.2023 | France |
| PP135024 | 18.09.2023 | USA |
| PP352386 | 31.12.2022 | USA |
| PQ618052 | 17.10.2022 | Panama |
| PQ638662 | 22.11.2022 | USA |
| OR522493 | 02.10.2022 | USA |
| OR522494 | 27.09.2022 | USA |
| OR143195 | 13.12.2022 | USA |
| PP270255 | 25.10.2023 | USA |
| PQ618048 | 21.06.2024 | Panama |
| PQ618049 | 21.06.2024 | Panama |
| PQ618046 | 28.06.2024 | Panama |
| PQ618045 | 11.07.2024 | Panama |
| PQ618047 | 11.07.2024 | Panama |
| PP795134 | 11.10.2023 | USA |
| PP084059 | 04.11.2023 | USA |
| PQ618050 | 11.07.2024 | Panama |
| PP084060 | 08.11.2023 | USA |
| PP795147 | 07.11.2023 | USA |
| PQ117653 | 12.01.2024 | USA |
| PP969969 | 18.10.2022 | Ireland |
| PQ638666 | 22.11.2022 | USA |
| PQ638699 | 11.11.2022 | USA |
| OR522481 | 06.12.2022 | USA |
| PP530268 | 28.11.2023 | USA |
| PP270234 | 01.11.2023 | USA |
| PP969943 | 04.10.2022 | Ireland |
| PP969951 | 22.09.2022 | Ireland |

| | | |
|---|---|---|
| PP454563 | 16.02.2023 | Saudi Arabia |
| OR162274 | 14.07.2022 | Kenya |
| PQ762676 | 08.10.2023 | France |
| PQ762736 | 04.12.2023 | France |
| OR522470 | 06.11.2022 | USA |
| OR975298 | 06.11.2022 | USA |
| PP970007 | 12.11.2022 | Ireland |
| PP352367 | 30.11.2022 | USA |
| PQ348870 | 11.09.2022 | United Kingdom |
| PQ348871 | 19.09.2022 | United Kingdom |
| OR872581 | 26.09.2022 | USA |
| PQ192488 | 05.10.2023 | Thailand |
| PQ348889 | 14.12.2022 | United Kingdom |
| PQ348874 | 28.09.2022 | United Kingdom |
| PQ348890 | 20.12.2022 | United Kingdom |
| PQ849774 | 07.12.2024 | USA |
| OR872604 | 01.08.2022 | USA |
| OP927864 | 19.01.2022 | Germany |
| OR143218 | 17.12.2022 | USA |
| PQ762495 | 13.03.2024 | USA |
| PQ788236 | 08.11.2024 | USA |
| PQ788244 | 24.10.2024 | USA |
| PQ348888 | 28.12.2022 | United Kingdom |
| PP529967 | 11.02.2023 | Hungary |
| PP970012 | 13.12.2022 | Ireland |
| PP969953 | 26.09.2022 | Ireland |
| PP970045 | 17.11.2023 | Ireland |
| PQ788229 | 13.11.2024 | USA |
| PQ788196 | 22.11.2024 | USA |
| PQ788220 | 19.11.2024 | USA |
| PQ788205 | 23.11.2024 | USA |
| PQ788237 | 05.11.2024 | USA |
| PP454562 | 31.12.2023 | Saudi Arabia |
| PP454570 | 13.10.2023 | Saudi Arabia |
| PP760393 | 31.10.2022 | USA |
| PQ763055 | 03.12.2023 | France |
| PP681261 | 14.12.2022 | USA |
| PP270266 | 05.12.2023 | USA |
| PP270277 | 03.12.2023 | USA |

| | | |
|---|---|---|
| PP847388 | 18.01.2024 | USA |
| PP978538 | 12.03.2024 | USA |
| PP957743 | 01.02.2024 | USA |
| PP978517 | 24.02.2024 | USA |
| PQ849781 | 09.12.2024 | USA |
| PQ008880 | 21.12.2023 | USA |
| OR143144 | 08.12.2022 | USA |
| PP681262 | 14.12.2022 | USA |
| PP495852 | 18.01.2024 | USA |
| PQ762871 | 17.12.2023 | France |
| PQ762741 | 03.12.2023 | France |
| PQ762868 | 02.12.2023 | France |
| PQ763137 | 29.11.2023 | France |
| PQ762991 | 26.11.2023 | France |
| PQ410476 | 01.01.2018 | Belgium |
| OQ248602 | 30.11.2022 | China |
| OR140550 | 01.05.2023 | China |
| OR140549 | 02.04.2023 | China |
| OR140554 | 01.01.2023 | China |
| OQ248606 | 01.12.2022 | China |
| OQ248607 | 31.12.2022 | China |
| OQ024158 | 05.11.2022 | USA |
| PP974164 | 10.12.2023 | China |
| PP974167 | 18.11.2022 | China |
| PQ626971 | 12.08.2024 | China |
| PP833560 | 01.03.2024 | China |
| PQ762685 | 26.09.2023 | France |
| OR840707 | 12.04.2023 | China |
| PP974163 | 18.12.2023 | China |
| PP974168 | 19.12.2023 | China |
| PP974169 | 03.01.2024 | China |
| OR840709 | 21.04.2023 | China |
| OR840706 | 11.04.2023 | China |
| PP970030 | 31.10.2023 | Ireland |
| PP974161 | 08.12.2023 | China |
| PP819400 | 14.01.2024 | China |
| PQ192487 | 28.11.2023 | Thailand |
| PP974162 | 22.12.2023 | China |
| PQ762684 | 24.09.2023 | France |

| | | |
|---|---|---|
| OR872636 | 11.10.2022 | USA |
| PP411979 | 13.08.2023 | Thailand |
| ON152648 | 18.10.2021 | Russia |
| OR143177 | 06.12.2022 | USA |
| PQ618058 | 11.08.2023 | Panama |
| PQ618059 | 27.02.2023 | Panama |
| OQ024138 | 14.11.2022 | USA |
| OQ024143 | 14.11.2022 | USA |
| OR143207 | 30.11.2022 | USA |
| PQ618057 | 12.06.2024 | Panama |
| OQ024157 | 14.11.2022 | USA |
| OQ171900 | 20.07.2022 | USA |
| OQ024135 | 05.11.2022 | USA |
| PP970011 | 29.11.2022 | Ireland |
| OQ024122 | 03.11.2022 | USA |
| MZ961930 | 31.10.2017 | China |
| OQ248601 | 01.11.2022 | China |
| OR140547 | 01.04.2023 | China |
| OR140552 | 31.05.2023 | China |
| PP974156 | 18.12.2023 | China |
| OR140544 | 01.02.2023 | China |
| OR140543 | 01.02.2023 | China |
| PP974159 | 18.11.2022 | China |
| OR140546 | 01.04.2023 | China |
| OR140545 | 01.04.2023 | China |
| OR140551 | 01.05.2023 | China |
| PP668167 | 03.12.2023 | USA |
| PP847377 | 05.01.2024 | USA |
| LC816568 | 10.08.2023 | Japan |
| PQ066217 | 18.12.2023 | USA |
| PP781382 | 06.01.2024 | USA |
| PP781414 | 04.01.2024 | USA |
| PQ762886 | 06.12.2023 | France |
| OR795475 | 07.11.2022 | Germany |
| OR795481 | 05.12.2022 | Germany |
| PP386343 | 27.10.2023 | USA |
| PP709453 | 01.01.2024 | USA |
| PQ762491 | 28.01.2024 | USA |
| PP910764 | 25.01.2024 | USA |

| | | |
|---|---|---|
| PP847395 | 21.01.2024 | USA |
| PQ788247 | 12.01.2024 | USA |
| PP709398 | 01.01.2024 | USA |
| PP594813 | 22.11.2023 | USA |
| PP504628 | 05.11.2023 | USA |
| PP386349 | 28.10.2023 | USA |
| PP386344 | 27.10.2023 | USA |
| PP386337 | 25.10.2023 | USA |
| PP135023 | 12.09.2023 | USA |
| PP847380 | 11.01.2024 | USA |
| PP978507 | 17.02.2024 | USA |
| PQ762940 | 04.12.2023 | France |
| PP352334 | 01.10.2022 | USA |
| PP352363 | 30.11.2022 | USA |
| PP681264 | 12.12.2022 | USA |
| PP342424 | 05.12.2023 | USA |
| PP342425 | 06.12.2023 | USA |
| PP795151 | 22.11.2023 | USA |
| PQ618056 | 23.05.2022 | Panama |
| PQ618055 | 31.10.2022 | Panama |
| OR882972 | 08.05.2023 | USA |
| PQ348895 | 17.10.2022 | United Kingdom |
| PQ348893 | 13.10.2022 | United Kingdom |
| PQ348892 | 10.10.2022 | United Kingdom |
| PP709379 | 22.12.2023 | USA |
| PP770466 | 14.02.2024 | USA |
| PP770461 | 19.01.2024 | USA |
| PQ638796 | 02.11.2023 | USA |
| PP084061 | 15.11.2023 | USA |
| PQ008879 | 10.11.2023 | USA |
| PP342430 | 09.12.2023 | USA |
| PP770452 | 31.01.2024 | USA |
| PP934458 | 11.03.2024 | USA |
| PP342423 | 05.12.2023 | USA |
| PQ638790 | 11.11.2023 | USA |
| PQ638799 | 02.11.2023 | USA |
| PQ638718 | 19.10.2023 | USA |
| PQ638715 | 11.10.2023 | USA |
| OR915772 | 13.08.2023 | USA |

| | | |
|---|---|---|
| PP903815 | 12.04.2024 | USA |
| PQ390740 | 25.11.2023 | USA |
| PQ638720 | 29.10.2023 | USA |
| PP770459 | 22.01.2024 | USA |
| PQ638784 | 12.12.2023 | USA |
| PP776593 | 29.02.2024 | USA |
| PP959040 | 20.02.2024 | USA |
| PP959047 | 10.01.2024 | USA |
| PP530264 | 15.12.2023 | USA |
| PP709370 | 05.12.2023 | USA |
| PQ390742 | 25.11.2023 | USA |
| PP709395 | 03.01.2024 | USA |
| PP709392 | 04.01.2024 | USA |
| PP709369 | 04.12.2023 | USA |
| PP530269 | 03.01.2024 | USA |
| PP847358 | 22.12.2023 | USA |
| PP830387 | 16.12.2023 | USA |
| PP401819 | 28.11.2023 | USA |
| PP709385 | 26.12.2023 | USA |
| LC846999 | 02.09.2019 | Myanmar |
| LC847002 | 12.09.2020 | Myanmar |
| LC847027 | 06.12.2021 | Myanmar |
| OQ024119 | 03.11.2022 | USA |
| OQ024123 | 03.11.2022 | USA |
| OR287942 | 13.01.2020 | USA |
| OR666541 | 01.12.2019 | China |
| LC847003 | 22.08.2020 | Myanmar |
| OQ933826 | 25.11.2021 | China |
| PQ117650 | 11.03.2024 | USA |
| PP974160 | 11.01.2024 | China |
| LC816567 | 26.10.2022 | Japan |
| OR795370 | 31.12.2018 | Germany |
| OR140542 | 31.01.2023 | China |
| OQ248604 | 01.12.2022 | China |
| OQ248603 | 01.12.2022 | China |
| PP352337 | 30.11.2022 | USA |
| OQ024111 | 08.11.2022 | USA |
| PP352378 | 01.11.2022 | USA |
| PQ834853 | 19.10.2022 | Mexico |

| | | |
|---|---|---|
| OR522460 | 26.10.2022 | USA |
| OR522461 | 26.10.2022 | USA |
| PQ638724 | 12.10.2022 | USA |
| PP760397 | 01.11.2022 | USA |
| PP760422 | 01.11.2022 | USA |
| OP890319 | 01.11.2022 | USA |
| OR522474 | 08.11.2022 | USA |
| OR975302 | 30.11.2022 | USA |
| OR522516 | 04.12.2022 | USA |
| OR975301 | 27.11.2022 | USA |
| OR522524 | 01.01.2023 | USA |
| PP401815 | 24.11.2023 | USA |
| PP352385 | 01.12.2022 | USA |
| PP681272 | 05.01.2023 | USA |
| PP352382 | 01.11.2022 | USA |
| OQ024141 | 14.11.2022 | USA |
| LC847044 | 23.07.2022 | Myanmar |
| LC847045 | 28.07.2022 | Myanmar |
| OR162280 | 28.01.2023 | Kenya |
| OR872605 | 30.10.2022 | USA |
| OR162279 | 28.01.2023 | Kenya |
| OP890315 | 01.10.2022 | USA |
| OR795478 | 22.11.2022 | Germany |
| PP760406 | 31.01.2023 | USA |
| PP910780 | 13.02.2024 | USA |
| PP847359 | 06.01.2024 | USA |
| PP504654 | 13.11.2023 | USA |
| PP978521 | 25.02.2024 | USA |
| PQ762520 | 08.05.2024 | USA |
| PQ762483 | 13.01.2024 | USA |
| PP386360 | 05.11.2023 | USA |
| PP957763 | 10.02.2024 | USA |
| PP973780 | 10.02.2024 | USA |
| OP690363 | 31.12.2020 | Taiwan |
| OR795480 | 07.12.2022 | Germany |
| OR162275 | 19.09.2022 | Kenya |
| PP970047 | 20.11.2023 | Ireland |
| PQ762961 | 12.12.2023 | France |
| PP957761 | 08.02.2024 | USA |

| | | |
|---|---|---|
| PP957749 | 01.02.2024 | USA |
| PP910767 | 26.01.2024 | USA |
| PQ762487 | 15.01.2024 | USA |
| PP847363 | 09.01.2024 | USA |
| PP668171 | 05.12.2023 | USA |
| PP594816 | 04.12.2023 | USA |
| PP504647 | 13.11.2023 | USA |
| PP237786 | 09.10.2023 | USA |
| PP237784 | 09.10.2023 | USA |
| PP709447 | 30.12.2023 | USA |
| PP847382 | 13.01.2024 | USA |
| PP668163 | 03.12.2023 | USA |
| PP401817 | 27.11.2023 | USA |
| PP594814 | 26.11.2023 | USA |
| PQ788252 | 15.01.2024 | USA |
| PP910752 | 26.12.2023 | USA |
| PP847378 | 11.01.2024 | USA |
| PQ762679 | 21.01.2024 | France |
| PP974158 | 10.12.2023 | China |
| PP411981 | 13.07.2023 | Thailand |
| PP411984 | 12.08.2023 | Thailand |
| PQ638721 | 01.08.2023 | USA |
| OQ248605 | 01.12.2022 | China |
| OR140553 | 01.05.2023 | China |
| PP974165 | 11.01.2024 | China |
| PP974166 | 20.12.2023 | China |
| PQ763068 | 05.12.2023 | France |
| PQ763039 | 12.11.2023 | France |
| PQ762756 | 05.12.2023 | France |
| PQ763143 | 08.12.2023 | France |
| OQ024128 | 03.11.2022 | USA |
| OR872641 | 10.10.2022 | USA |
| OR872608 | 22.06.2022 | USA |
| OR872623 | 14.07.2022 | USA |
| OQ024115 | 02.11.2022 | USA |
| PP795131 | 03.10.2023 | USA |
| PQ834859 | 30.11.2022 | Mexico |
| PP830388 | 17.12.2023 | USA |
| PP270242 | 14.11.2023 | USA |

| | | |
|---|---|---|
| PP352379 | 31.12.2022 | USA |
| PP781419 | 03.01.2024 | USA |
| PP504630 | 02.11.2023 | USA |
| PP401816 | 23.11.2023 | USA |
| PP709450 | 30.12.2023 | USA |
| PP957780 | 13.04.2024 | USA |
| PQ762512 | 20.04.2024 | USA |
| PP681277 | 10.01.2023 | USA |
| OQ261753 | 02.11.2022 | Austria |
| OP890340 | 31.10.2022 | USA |
| OR143213 | 17.12.2022 | USA |
| OR143139 | 14.12.2022 | USA |
| OR143212 | 08.12.2022 | USA |
| OR143204 | 04.12.2022 | USA |
| PP681274 | 03.01.2023 | USA |
| PQ834906 | 02.01.2024 | Mexico |
| PQ834902 | 13.12.2023 | Mexico |
| PQ834880 | 08.11.2023 | Mexico |
| PQ834881 | 13.11.2023 | Mexico |
| PQ834888 | 22.11.2023 | Mexico |
| PQ834904 | 13.12.2023 | Mexico |
| PQ618043 | 22.07.2024 | Panama |
| PQ618044 | 05.07.2024 | Panama |
| PQ008876 | 13.12.2023 | USA |
| PP342429 | 08.12.2023 | USA |
| PP342433 | 17.12.2023 | USA |
| PP342440 | 26.12.2023 | USA |
| PP342436 | 19.12.2023 | USA |
| PQ008871 | 29.12.2023 | USA |
| PQ008872 | 29.12.2023 | USA |
| OY757830 | 15.09.2022 | Australia |
| PP454569 | 04.06.2023 | Saudi Arabia |
| PP969976 | 06.11.2022 | Ireland |
| PP969974 | 02.11.2022 | Ireland |
| PQ066221 | 17.04.2024 | USA |
| PQ788231 | 08.11.2024 | USA |
| PQ849778 | 06.12.2024 | USA |
| PQ849773 | 07.12.2024 | USA |
| PQ849797 | 01.12.2024 | USA |

| | | |
|---|---|---|
| PQ788227 | 14.11.2024 | USA |
| PQ788200 | 25.11.2024 | USA |
| PQ788212 | 04.11.2024 | USA |
| PQ849782 | 09.12.2024 | USA |
| PQ788232 | 09.11.2024 | USA |
| PQ849771 | 09.12.2024 | USA |
| PQ788235 | 12.11.2024 | USA |
| PQ788210 | 22.11.2024 | USA |
| PQ788211 | 22.11.2024 | USA |
| PQ788209 | 24.11.2024 | USA |
| PQ788224 | 19.11.2024 | USA |
| PQ788238 | 05.11.2024 | USA |
| PQ849779 | 05.12.2024 | USA |
| PQ849799 | 29.11.2024 | USA |
| PQ788234 | 12.11.2024 | USA |
| PQ788228 | 14.11.2024 | USA |
| PQ849775 | 06.12.2024 | USA |
| PQ849785 | 08.12.2024 | USA |
| PQ788241 | 31.10.2024 | USA |
| PQ849794 | 28.11.2024 | USA |
| PQ849792 | 29.11.2024 | USA |
| PQ849776 | 06.12.2024 | USA |
| PQ788197 | 24.11.2024 | USA |
| PQ788233 | 09.11.2024 | USA |
| PQ849788 | 01.12.2024 | USA |
| PQ788239 | 03.11.2024 | USA |
| PQ849800 | 27.11.2024 | USA |
| PQ788226 | 16.11.2024 | USA |
| PQ788222 | 19.11.2024 | USA |
| PQ788198 | 27.11.2024 | USA |
| PQ849793 | 03.12.2024 | USA |
| PQ849786 | 08.12.2024 | USA |
| PQ788207 | 26.11.2024 | USA |
| PQ788221 | 19.11.2024 | USA |
| OR162270 | 12.06.2022 | Kenya |
| OR162262 | 26.04.2022 | Kenya |
| OR162254 | 21.03.2022 | Kenya |
| OR162276 | 19.03.2022 | Kenya |
| PP530261 | 03.01.2024 | USA |

| | | |
|---|---|---|
| PP709373 | 18.12.2023 | USA |
| PP795165 | 11.02.2024 | USA |
| PQ762511 | 20.03.2024 | USA |
| PQ762498 | 13.03.2024 | USA |
| OR143202 | 06.01.2023 | USA |
| OR522468 | 06.11.2022 | USA |
| OR882992 | 28.02.2023 | USA |
| OR882988 | 26.02.2023 | USA |
| PP594804 | 22.11.2023 | USA |
| PQ638725 | 12.10.2022 | USA |
| PQ638723 | 12.10.2022 | USA |
| PQ638717 | 03.11.2022 | USA |
| OR502561 | 30.09.2021 | India |
| PP969973 | 01.11.2022 | Ireland |
| PP970010 | 13.11.2022 | Ireland |
| PP969978 | 13.11.2022 | Ireland |
| PP969975 | 06.11.2022 | Ireland |
| PP969967 | 13.10.2022 | Ireland |
| PP352353 | 31.10.2022 | USA |
| PP352371 | 01.12.2022 | USA |
| PQ788243 | 27.10.2024 | USA |
| PP270237 | 14.11.2023 | USA |
| PQ348968 | 10.09.2022 | United Kingdom |
| PP781405 | 08.01.2024 | USA |
| PP781384 | 03.01.2024 | USA |
| PP270256 | 14.11.2023 | USA |
| PP270239 | 25.10.2023 | USA |
| OQ171918 | 19.07.2022 | USA |
| OQ024125 | 05.11.2022 | USA |
| OQ024150 | 05.11.2022 | USA |
| PP978530 | 05.03.2024 | USA |
| PQ788204 | 23.11.2024 | USA |
| PQ788214 | 11.11.2024 | USA |
| PQ788218 | 20.11.2024 | USA |
| PQ788202 | 04.10.2024 | USA |
| PQ788201 | 26.11.2024 | USA |
| PQ788219 | 20.11.2024 | USA |
| PP135009 | 08.08.2023 | USA |
| OR143210 | 01.01.2023 | USA |

| | | |
|---|---|---|
| PP352355 | 01.10.2022 | USA |
| PP681251 | 08.12.2022 | USA |
| PP352381 | 01.12.2022 | USA |
| OR872627 | 28.10.2022 | USA |
| PP270241 | 10.10.2023 | USA |
| OP890322 | 01.10.2022 | USA |
| PP681281 | 23.11.2022 | USA |
| PP352364 | 30.11.2022 | USA |
| PP934451 | 03.03.2024 | USA |
| PP973771 | 05.02.2024 | USA |
| PP934461 | 12.03.2024 | USA |
| PP852061 | 11.02.2024 | USA |
| PP973779 | 09.02.2024 | USA |
| OR872600 | 12.10.2022 | USA |
| PP781410 | 22.12.2023 | USA |
| PP270257 | 04.12.2023 | USA |
| OR915737 | 31.10.2022 | USA |
| PP903805 | 31.10.2022 | USA |
| OR883010 | 28.10.2022 | USA |
| OR882969 | 18.05.2023 | USA |
| PP903830 | 28.10.2022 | USA |
| OR915760 | 24.10.2022 | USA |
| PP969948 | 28.06.2022 | Ireland |
| OQ261748 | 09.09.2021 | Austria |
| PP386359 | 03.11.2023 | USA |
| PP970059 | 13.11.2022 | Ireland |
| PQ762768 | 15.12.2023 | France |
| PQ762850 | 05.12.2023 | France |
| PQ763159 | 26.11.2023 | France |
| PQ762690 | 26.09.2023 | France |
| PQ762724 | 23.11.2023 | France |
| PQ763139 | 05.12.2023 | France |
| PQ762915 | 01.12.2023 | France |
| PQ762817 | 08.11.2023 | France |
| PQ762864 | 22.12.2023 | France |
| PQ762845 | 07.11.2023 | France |
| PQ762848 | 04.12.2023 | France |
| PQ762983 | 07.01.2024 | France |
| PQ762860 | 18.12.2023 | France |

| | | |
|---|---|---|
| PQ763122 | 13.12.2023 | France |
| PQ762852 | 10.12.2023 | France |
| PQ762846 | 01.12.2023 | France |
| PQ762865 | 20.12.2023 | France |
| PQ762862 | 30.12.2023 | France |
| PQ762861 | 21.12.2023 | France |
| PQ762844 | 06.11.2023 | France |
| PQ762975 | 12.12.2023 | France |
| PQ763073 | 12.12.2023 | France |
| PQ762732 | 27.11.2023 | France |
| PQ762779 | 30.11.2023 | France |
| PQ762974 | 11.12.2023 | France |
| PQ762956 | 05.11.2023 | France |
| PQ762951 | 10.01.2024 | France |
| PQ762953 | 15.10.2023 | France |
| PQ763147 | 29.12.2023 | France |
| PQ762898 | 03.11.2023 | France |
| PQ762905 | 19.12.2023 | France |
| PQ763031 | 09.02.2024 | France |
| PQ762784 | 12.12.2023 | France |
| PQ763168 | 28.11.2023 | France |
| PQ762890 | 18.12.2023 | France |
| PQ763067 | 03.12.2023 | France |
| PQ762906 | 29.12.2023 | France |
| PQ762774 | 28.12.2023 | France |
| PQ762651 | 16.12.2023 | France |
| PQ762879 | 02.11.2023 | France |
| PQ763034 | 25.01.2024 | France |
| PQ762881 | 04.12.2023 | France |
| PQ763146 | 27.12.2023 | France |
| PQ762646 | 07.12.2023 | France |
| PQ762785 | 06.01.2024 | France |
| PQ762877 | 28.11.2023 | France |
| PQ762964 | 09.12.2023 | France |
| PQ762965 | 28.01.2024 | France |
| PQ762999 | 25.11.2023 | France |
| PQ762824 | 02.11.2023 | France |
| PQ762826 | 17.12.2023 | France |
| PQ762667 | 02.12.2023 | France |

| | | |
|---|---|---|
| PQ762757 | 07.12.2023 | France |
| PQ762904 | 08.12.2023 | France |
| PQ762699 | 07.12.2023 | France |
| PQ762836 | 27.11.2023 | France |
| PQ762751 | 10.11.2023 | France |
| PQ762838 | 04.12.2023 | France |
| PQ117652 | 09.01.2024 | USA |
| PQ117648 | 15.12.2023 | USA |
| PQ762707 | 17.12.2023 | France |
| PQ762762 | 19.12.2023 | France |
| PQ762728 | 24.11.2023 | France |
| PQ762710 | 20.12.2023 | France |
| PQ762680 | 03.12.2023 | France |
| PQ762884 | 31.12.2023 | France |
| PQ762926 | 25.11.2023 | France |
| PQ762797 | 15.12.2023 | France |
| PQ763045 | 23.11.2023 | France |
| PQ762970 | 21.11.2023 | France |
| PQ762992 | 28.11.2023 | France |
| KX453505 | 29.04.2015 | Kenya |
| OR795343 | 30.12.2013 | Germany |
| KX765977 | 12.02.2014 | New Zealand |
| MH142228 | 20.03.2014 | Russia |
| PP660602 | 16.03.2021 | South Africa |
| MH182010 | 10.12.2014 | Kenya |
| KX453394 | 19.04.2014 | Kenya |
| OP744445 | 25.03.2022 | Kenya |
| OR162211 | 18.12.2022 | Kenya |
| OR162239 | 04.03.2022 | Kenya |
| OP744443 | 22.03.2022 | Kenya |
| OP744449 | 01.04.2021 | Kenya |
| PQ638686 | 07.01.2023 | USA |
| PQ638701 | 12.01.2023 | USA |
| PQ762856 | 12.12.2023 | France |
| PQ762847 | 04.12.2023 | France |
| PQ762662 | 17.12.2023 | France |
| PQ762855 | 12.12.2023 | France |
| PQ763038 | 20.10.2023 | France |
| PQ762962 | 11.12.2023 | France |

| | | |
|---|---|---|
| PQ762857 | 13.12.2023 | France |
| MZ516118 | 27.12.2019 | Netherlands |
| PQ762808 | 02.12.2023 | France |
| PQ763119 | 01.01.2024 | France |
| PP760414 | 31.12.2022 | USA |
| PP203266 | 10.09.2022 | USA |
| PP203262 | 10.09.2022 | USA |
| OP320398 | 04.02.2020 | Philippines |
| OP320399 | 09.03.2020 | Philippines |
| PP681245 | 05.12.2022 | USA |
| PP594810 | 23.11.2023 | USA |
| OR872615 | 25.10.2022 | USA |
| OR883003 | 19.03.2023 | USA |
| PQ410470 | 31.12.2022 | Belgium |
| PQ348887 | 21.12.2022 | United Kingdom |
| PP969994 | 30.11.2022 | Ireland |
| PP681257 | 07.12.2022 | USA |
| OR795477 | 15.11.2022 | Germany |
| PQ762800 | 26.11.2023 | France |
| PP969972 | 28.10.2022 | Ireland |
| PP969983 | 08.11.2022 | Ireland |
| PQ348878 | 09.10.2022 | United Kingdom |
| PQ348863 | 31.08.2022 | United Kingdom |
| PQ348866 | 30.08.2022 | United Kingdom |
| PQ348964 | 10.10.2022 | United Kingdom |
| PQ348966 | 31.12.2022 | United Kingdom |
| PQ348885 | 21.10.2022 | United Kingdom |
| PQ348873 | 22.09.2022 | United Kingdom |
| PP270238 | 07.11.2023 | USA |
| PQ762818 | 25.11.2023 | France |
| PQ737354 | 30.10.2023 | France |
| PQ762631 | 30.10.2023 | France |
| PQ762903 | 08.01.2024 | France |
| PQ008873 | 03.03.2024 | USA |
| PQ008875 | 25.02.2024 | USA |
| OR601480 | 15.08.2023 | USA |
| PQ638789 | 11.11.2023 | USA |
| PP594805 | 25.11.2023 | USA |
| PP342417 | 28.11.2023 | USA |

| | | |
|---|---|---|
| PQ762675 | 18.12.2023 | France |
| PP454568 | 01.01.2023 | Saudi Arabia |
| OR045924 | 01.01.2021 | United Arab Emirates |
| OQ980512 | 31.12.2021 | United Arab Emirates |
| OR162263 | 03.05.2022 | Kenya |
| OR162277 | 30.08.2022 | Kenya |
| OR162273 | 06.07.2022 | Kenya |
| OR162248 | 10.03.2022 | Kenya |
| OR162264 | 03.05.2022 | Kenya |
| OR162243 | 09.03.2022 | Kenya |
| OR162236 | 01.03.2022 | Kenya |
| OR162241 | 07.03.2022 | Kenya |
| OR162244 | 09.03.2022 | Kenya |
| OR162240 | 06.03.2022 | Kenya |
| OP744446 | 25.03.2022 | Kenya |
| PP957752 | 05.02.2024 | USA |
| PQ762524 | 15.05.2024 | USA |
| PP957775 | 09.04.2024 | USA |
| PP978515 | 23.02.2024 | USA |
| PP973770 | 05.02.2024 | USA |
| PP847389 | 20.01.2024 | USA |
| PP847381 | 12.01.2024 | USA |
| PP847384 | 12.01.2024 | USA |
| PP847362 | 09.01.2024 | USA |
| PP847365 | 08.01.2024 | USA |
| PP830405 | 23.12.2023 | USA |
| PP830402 | 21.12.2023 | USA |
| PP830390 | 16.12.2023 | USA |
| PP830382 | 10.12.2023 | USA |
| PP830381 | 08.12.2023 | USA |
| PP668166 | 30.11.2023 | USA |
| PP594797 | 25.11.2023 | USA |
| PP668162 | 20.11.2023 | USA |
| PP504641 | 11.11.2023 | USA |
| PP504648 | 09.11.2023 | USA |
| PP386361 | 06.11.2023 | USA |
| PP386363 | 05.11.2023 | USA |
| PP504636 | 03.11.2023 | USA |

| | | |
|---|---|---|
| PP504637 | 03.11.2023 | USA |
| PP386364 | 02.11.2023 | USA |
| PP386352 | 28.10.2023 | USA |
| PP386355 | 28.10.2023 | USA |
| PP386353 | 28.10.2023 | USA |
| PP386338 | 25.10.2023 | USA |
| PP237795 | 19.10.2023 | USA |
| PP237792 | 16.10.2023 | USA |
| PP237787 | 05.10.2023 | USA |
| PP237780 | 29.09.2023 | USA |
| PP135034 | 25.09.2023 | USA |
| PP135019 | 11.09.2023 | USA |
| PP135016 | 30.08.2023 | USA |
| PP135013 | 25.08.2023 | USA |
| PP781381 | 03.01.2024 | USA |
| PP270278 | 26.11.2023 | USA |
| PP594815 | 04.12.2023 | USA |
| PP668170 | 02.12.2023 | USA |
| PP237794 | 13.10.2023 | USA |
| PP504631 | 05.11.2023 | USA |
| PP830406 | 24.12.2023 | USA |
| PP847376 | 05.01.2024 | USA |
| PP709454 | 03.01.2024 | USA |
| PP709384 | 28.12.2023 | USA |
| PP504627 | 03.11.2023 | USA |
| PP594799 | 23.11.2023 | USA |
| PP709452 | 01.01.2024 | USA |
| PP504633 | 02.11.2023 | USA |
| PP830393 | 14.12.2023 | USA |
| PP709380 | 24.12.2023 | USA |
| PP830400 | 22.12.2023 | USA |
| PP830413 | 23.12.2023 | USA |
| PP957744 | 01.02.2024 | USA |
| PP910762 | 22.01.2024 | USA |
| PP504626 | 04.11.2023 | USA |
| PP386366 | 05.11.2023 | USA |
| PP830403 | 24.12.2023 | USA |
| PP830409 | 22.12.2023 | USA |
| PQ762497 | 13.03.2024 | USA |

| | | |
|---|---|---|
| PP781380 | 07.02.2024 | USA |
| PQ762484 | 14.01.2024 | USA |
| PP237790 | 13.10.2023 | USA |
| PP594798 | 21.11.2023 | USA |
| PP386346 | 28.10.2023 | USA |
| PP978510 | 22.02.2024 | USA |
| PP504651 | 11.11.2023 | USA |
| PP709459 | 02.01.2024 | USA |
| PP504652 | 10.11.2023 | USA |
| PP386348 | 28.10.2023 | USA |
| PP386351 | 28.10.2023 | USA |
| PP978509 | 22.02.2024 | USA |
| PP237785 | 09.10.2023 | USA |
| PP978511 | 22.02.2024 | USA |
| PP709440 | 25.12.2023 | USA |
| PP957768 | 22.03.2024 | USA |
| PP504645 | 10.11.2023 | USA |
| PP401811 | 03.11.2023 | USA |
| PP237788 | 08.10.2023 | USA |
| PP910774 | 28.01.2024 | USA |
| PQ762503 | 16.03.2024 | USA |
| PQ762482 | 13.01.2024 | USA |
| PP910756 | 11.01.2024 | USA |
| PP668168 | 30.11.2023 | USA |
| PP386341 | 27.10.2023 | USA |
| PP668176 | 16.12.2023 | USA |
| PP847360 | 06.01.2024 | USA |
| PP709394 | 04.01.2024 | USA |
| PP386357 | 04.11.2023 | USA |
| PP910772 | 27.01.2024 | USA |
| PP978528 | 03.03.2024 | USA |
| PP386358 | 04.11.2023 | USA |
| PP504655 | 09.11.2023 | USA |
| PP504635 | 06.11.2023 | USA |
| PP386356 | 02.11.2023 | USA |
| PQ762488 | 30.01.2024 | USA |
| PQ762494 | 12.03.2024 | USA |
| PP668173 | 19.12.2023 | USA |
| PP978514 | 23.02.2024 | USA |

| | | |
|---|---|---|
| PP830391 | 16.12.2023 | USA |
| PP847366 | 08.01.2024 | USA |
| PQ762490 | 30.01.2024 | USA |
| PP401813 | 09.11.2023 | USA |
| PP830399 | 15.12.2023 | USA |
| PP709376 | 21.12.2023 | USA |
| PP237789 | 08.10.2023 | USA |
| PP386342 | 27.10.2023 | USA |
| PP386362 | 04.11.2023 | USA |
| PP594807 | 24.11.2023 | USA |
| PP957755 | 04.02.2024 | USA |
| PP504656 | 14.11.2023 | USA |
| PP594812 | 21.11.2023 | USA |
| PP957746 | 02.02.2024 | USA |
| PQ762492 | 28.01.2024 | USA |
| PP910766 | 26.01.2024 | USA |
| PQ788251 | 13.01.2024 | USA |
| PQ788249 | 12.01.2024 | USA |
| PP847371 | 10.01.2024 | USA |
| PP709457 | 31.12.2023 | USA |
| PQ762480 | 22.12.2023 | USA |
| PP668164 | 04.12.2023 | USA |
| PP504653 | 09.11.2023 | USA |
| PP504629 | 04.11.2023 | USA |
| PP386350 | 28.10.2023 | USA |
| PP386339 | 27.10.2023 | USA |
| PP386345 | 27.10.2023 | USA |
| PP237791 | 12.10.2023 | USA |
| PP237783 | 04.10.2023 | USA |
| PP237781 | 01.10.2023 | USA |
| PP957747 | 02.02.2024 | USA |
| PP978513 | 23.02.2024 | USA |
| MH388041 | 09.02.2015 | Saudi Arabia |
| MK182718 | 11.01.2013 | Saudi Arabia |
| OM256492 | 01.01.2018 | |
| OR143190 | 30.11.2022 | USA |
| OR522515 | 13.11.2022 | USA |
| OP320383 | 14.09.2019 | Philippines |
| PP970009 | 09.12.2022 | Ireland |

| | | |
|---|---|---|
| PQ763033 | 09.02.2024 | France |
| PQ762968 | 20.11.2023 | France |
| PQ762909 | 15.12.2023 | France |
| PQ762854 | 11.12.2023 | France |
| PQ762849 | 04.12.2023 | France |
| PQ763018 | 30.12.2023 | France |
| PQ763021 | 16.12.2023 | France |
| PP508189 | 18.05.2023 | Peru |
| PP969993 | 29.11.2023 | Ireland |
| PP969960 | 27.11.2023 | Ireland |
| PP970001 | 17.11.2023 | Ireland |
| PQ762698 | 06.12.2023 | France |
| PQ762700 | 08.12.2023 | France |
| PP959049 | 12.01.2024 | USA |
| PQ762739 | 30.11.2023 | France |
| PQ762839 | 07.12.2023 | France |
| PQ762842 | 31.12.2023 | France |
| PQ762777 | 08.02.2024 | France |
| PQ762781 | 04.12.2023 | France |
| PQ763062 | 12.11.2023 | France |
| PQ762720 | 29.10.2023 | France |
| PQ762912 | 28.10.2023 | France |
| PQ762780 | 05.12.2023 | France |
| PQ762791 | 02.11.2023 | France |
| PQ762687 | 18.11.2023 | France |
| PQ762716 | 10.01.2024 | France |
| PQ762874 | 28.11.2023 | France |
| PQ762713 | 25.12.2023 | France |
| PQ763053 | 21.12.2023 | France |
| PQ762682 | 24.11.2023 | France |
| PQ762793 | 26.11.2023 | France |
| PQ762835 | 27.11.2023 | France |
| PQ763134 | 05.01.2024 | France |
| PQ762674 | 04.12.2023 | France |
| PQ762830 | 12.12.2023 | France |
| PQ762966 | 29.01.2024 | France |
| PP969947 | 18.06.2022 | Ireland |
| PP770464 | 15.01.2024 | USA |
| PP709780 | 04.04.2022 | Russia |

| | | |
|---|---|---|
| OR522490 | 27.11.2022 | USA |
| PP342421 | 03.12.2023 | USA |
| PP342437 | 19.12.2023 | USA |
| PP352358 | 01.10.2022 | USA |
| OR883005 | 07.11.2022 | USA |
| OR915744 | 05.11.2022 | USA |
| OR915758 | 05.11.2022 | USA |
| OR883000 | 15.10.2022 | USA |
| PP903813 | 11.10.2022 | USA |
| PP903817 | 01.10.2022 | USA |
| OR872620 | 12.09.2022 | USA |
| OR915728 | 03.11.2022 | USA |
| OR915738 | 01.11.2022 | USA |
| PQ638685 | 22.01.2023 | USA |
| PQ117651 | 02.01.2024 | USA |
| PP352329 | 01.10.2022 | USA |
| PP709448 | 30.12.2023 | USA |
| PP795158 | 02.01.2024 | USA |
| OR872624 | 16.07.2022 | USA |
| OR872599 | 10.09.2022 | USA |
| OR872630 | 15.10.2022 | USA |
| PQ638793 | 03.11.2023 | USA |
| PP957777 | 05.04.2024 | USA |
| PQ762514 | 21.04.2024 | USA |
| PQ849791 | 02.12.2024 | USA |
| PQ008878 | 12.02.2024 | USA |
| PP781386 | 08.01.2024 | USA |
| PP781408 | 29.02.2024 | USA |
| PP270240 | 11.10.2023 | USA |
| PP495850 | 11.01.2024 | USA |
| PQ638795 | 07.11.2023 | USA |
| PQ638713 | 14.10.2023 | USA |
| PP970038 | 09.11.2023 | Ireland |
| PQ834893 | 04.12.2023 | Mexico |
| OQ024142 | 05.11.2022 | USA |
| OR872639 | 27.05.2022 | USA |
| OR882999 | 06.12.2022 | USA |
| OQ024112 | 06.11.2022 | USA |
| OQ024153 | 03.11.2022 | USA |

| | | |
|---|---|---|
| PP454565 | 22.06.2023 | Saudi Arabia |
| PQ348896 | 18.10.2022 | United Kingdom |
| OR143194 | 19.12.2022 | USA |
| OR143221 | 18.12.2022 | USA |
| OR143149 | 10.12.2022 | USA |
| PQ349024 | 12.02.2023 | United Kingdom |
| PQ762959 | 24.11.2023 | France |
| PQ762670 | 26.10.2023 | France |
| PQ762669 | 13.10.2023 | France |
| PQ762747 | 28.10.2023 | France |
| PQ762869 | 12.12.2023 | France |
| PQ762867 | 03.12.2023 | France |
| PQ762929 | 01.12.2023 | France |
| PQ762963 | 09.12.2023 | France |
| PQ762717 | 18.01.2024 | France |
| PQ763019 | 27.12.2023 | France |
| PQ762678 | 29.10.2023 | France |
| PQ762788 | 01.01.2024 | France |
| PQ762772 | 29.12.2023 | France |
| PQ762967 | 19.11.2023 | France |
| PQ762737 | 08.12.2023 | France |
| PQ763036 | 21.03.2024 | France |
| PQ762831 | 19.11.2023 | France |
| PQ762660 | 15.01.2024 | France |
| PQ762958 | 13.01.2024 | France |
| PQ762657 | 02.01.2024 | France |
| PQ762775 | 27.12.2023 | France |
| PQ762654 | 22.12.2023 | France |
| PQ762641 | 05.12.2023 | France |
| PQ737362 | 26.11.2023 | France |
| PQ737358 | 18.11.2023 | France |
| PQ762714 | 26.12.2023 | France |
| PQ762989 | 31.12.2023 | France |
| PQ737368 | 09.12.2023 | France |
| PQ762635 | 18.11.2023 | France |
| PQ762770 | 21.12.2023 | France |
| PQ762652 | 18.12.2023 | France |
| PQ762636 | 20.11.2023 | France |
| PQ763020 | 31.12.2023 | France |

| | | |
|---|---|---|
| PQ762647 | 12.12.2023 | France |
| PQ737364 | 05.12.2023 | France |
| PQ762648 | 13.12.2023 | France |
| PQ737359 | 20.11.2023 | France |
| PQ762650 | 14.12.2023 | France |
| PQ762649 | 13.12.2023 | France |
| PQ737365 | 05.12.2023 | France |
| PQ763043 | 13.12.2023 | France |
| PQ762642 | 05.12.2023 | France |
| PQ737363 | 27.11.2023 | France |
| PQ762640 | 27.11.2023 | France |
| PQ762639 | 26.11.2023 | France |
| PQ762645 | 09.12.2023 | France |
| PP970053 | 22.10.2022 | Ireland |
| PP970050 | 14.10.2022 | Ireland |
| PP970057 | 07.11.2022 | Ireland |
| PP970060 | 11.11.2022 | Ireland |
| PQ348894 | 13.10.2022 | United Kingdom |
| PQ349020 | 15.11.2022 | United Kingdom |
| PQ349016 | 16.08.2022 | United Kingdom |
| PQ348891 | 07.10.2022 | United Kingdom |
| PQ349025 | 12.04.2023 | United Kingdom |
| OR522478 | 23.10.2022 | USA |
| PP203261 | 19.09.2022 | USA |
| PQ638707 | 16.11.2022 | USA |
| PQ638677 | 12.11.2022 | USA |
| OR522486 | 12.10.2022 | USA |
| PQ349011 | 30.07.2022 | United Kingdom |
| PQ788230 | 08.11.2024 | USA |
| PQ849802 | 19.08.2024 | USA |
| OK500268 | 30.04.2021 | France |
| PQ762760 | 16.12.2023 | France |
| PQ762840 | 27.12.2023 | France |
| PQ763026 | 27.11.2023 | France |
| PQ762727 | 18.11.2023 | France |
| OR872609 | 11.05.2022 | USA |
| PP970006 | 16.11.2022 | Ireland |
| OR872617 | 22.09.2022 | USA |
| PP970015 | 12.11.2022 | Ireland |

| | | |
|---|---|---|
| OR872638 | 19.09.2022 | USA |
| PP969954 | 26.09.2022 | Ireland |
| PP969979 | 01.12.2022 | Ireland |
| PP957779 | 04.04.2024 | USA |
| PP973773 | 10.02.2024 | USA |
| PP973766 | 06.02.2024 | USA |
| PP973765 | 04.02.2024 | USA |
| PP973763 | 03.02.2024 | USA |
| PP709387 | 28.12.2023 | USA |
| PP882669 | 28.11.2023 | USA |
| PP386354 | 28.10.2023 | USA |
| PP957774 | 03.04.2024 | USA |
| PP973755 | 28.01.2024 | USA |
| PP709378 | 23.12.2023 | USA |
| PP973777 | 16.02.2024 | USA |
| PP910761 | 20.01.2024 | USA |
| PP709401 | 09.01.2024 | USA |
| PP709399 | 05.01.2024 | USA |
| PP934457 | 05.03.2024 | USA |
| PP934462 | 17.03.2024 | USA |
| PP973775 | 10.02.2024 | USA |
| PP709393 | 01.01.2024 | USA |
| PP830386 | 14.12.2023 | USA |
| PP957753 | 02.02.2024 | USA |
| PP709400 | 09.01.2024 | USA |
| PP830380 | 06.12.2023 | USA |
| PP934453 | 29.02.2024 | USA |
| PP910754 | 14.01.2024 | USA |
| PQ762515 | 24.04.2024 | USA |
| PP973772 | 07.02.2024 | USA |
| PP973774 | 10.02.2024 | USA |
| PP830385 | 11.12.2023 | USA |
| PP934454 | 29.02.2024 | USA |
| PP973764 | 04.02.2024 | USA |
| PP709371 | 19.12.2023 | USA |
| PQ638702 | 26.02.2023 | USA |
| PQ638727 | 23.02.2023 | USA |
| OQ024131 | 05.11.2022 | USA |
| OQ024147 | 03.11.2022 | USA |

| | | |
|---|---|---|
| PP508184 | 21.04.2023 | Peru |
| PP508185 | 21.04.2023 | Peru |
| PP508183 | 18.04.2023 | Peru |
| OR882982 | 10.03.2023 | USA |
| PP270273 | 08.12.2023 | USA |
| PP781398 | 26.12.2023 | USA |
| PQ763025 | 24.12.2023 | France |
| PQ762811 | 28.11.2023 | France |
| PQ762949 | 11.12.2023 | France |
| PQ762665 | 01.12.2023 | France |
| PQ763024 | 25.11.2023 | France |
| PQ762812 | 16.12.2023 | France |
| OQ024117 | 03.11.2022 | USA |
| OQ171924 | 26.10.2022 | USA |
| OR915740 | 10.11.2022 | USA |
| OP890330 | 30.11.2022 | USA |
| PP681258 | 12.12.2022 | USA |
| OR872590 | 23.10.2022 | USA |
| OR522531 | 03.01.2023 | USA |
| OR601471 | 09.10.2022 | USA |
| OR522530 | 03.01.2023 | USA |
| OR975304 | 22.12.2022 | USA |
| PP781393 | 30.01.2024 | USA |
| PP882672 | 18.10.2023 | USA |
| PP681266 | 13.12.2022 | USA |
| PP681249 | 07.12.2022 | USA |
| PP681246 | 07.12.2022 | USA |
| PP681271 | 28.12.2022 | USA |
| PP681282 | 27.11.2022 | USA |
| PP681255 | 08.12.2022 | USA |
| PP681244 | 29.11.2022 | USA |
| PP957754 | 05.02.2024 | USA |
| PP352383 | 01.12.2022 | USA |
| OR872586 | 24.06.2022 | USA |
| PQ638687 | 11.11.2022 | USA |
| OR872635 | 25.10.2022 | USA |
| PP352336 | 01.11.2022 | USA |
| OR872594 | 11.07.2022 | USA |
| PP795143 | 29.10.2023 | USA |

| | | |
|---|---|---|
| PP795150 | 15.11.2023 | USA |
| PQ763121 | 12.12.2023 | France |
| PQ638710 | 16.10.2022 | USA |
| PP770462 | 14.01.2024 | USA |
| PQ638712 | 09.08.2023 | USA |
| PQ638785 | 23.11.2023 | USA |
| PQ638794 | 03.11.2023 | USA |
| OR795472 | 14.10.2022 | Germany |
| PQ762806 | 30.11.2023 | France |
| PQ762672 | 01.12.2023 | France |
| PQ618068 | 12.07.2024 | Panama |
| PQ618071 | 30.05.2024 | Panama |
| PQ618070 | 28.06.2024 | Panama |
| PQ618069 | 14.06.2024 | Panama |
| PQ618074 | 27.06.2024 | Panama |
| PQ618073 | 05.07.2024 | Panama |
| PQ618072 | 05.07.2024 | Panama |
| PP270269 | 18.12.2023 | USA |
| PP978508 | 18.02.2024 | USA |
| PP978531 | 06.03.2024 | USA |
| PP530278 | 03.11.2023 | USA |
| PQ618076 | 01.08.2023 | Panama |
| PQ763044 | 12.02.2024 | France |
| PP934467 | 24.03.2024 | USA |
| PP508188 | 18.05.2023 | Peru |
| PQ618075 | 11.12.2023 | Panama |
| PQ618077 | 19.11.2023 | Panama |
| PP969990 | 28.11.2023 | Ireland |
| PP970031 | 29.10.2023 | Ireland |
| PP970027 | 25.10.2023 | Ireland |
| PP681259 | 30.11.2022 | USA |
| PP969981 | 16.11.2022 | Ireland |
| PQ349021 | 21.11.2022 | United Kingdom |
| OQ261752 | 27.10.2022 | Austria |
| PP969971 | 25.10.2022 | Ireland |
| PP969964 | 03.10.2022 | Ireland |
| PP969968 | 16.10.2022 | Ireland |
| PP969965 | 11.10.2022 | Ireland |
| PQ410473 | 10.11.2023 | Belgium |

| | | |
|---|---|---|
| PQ763082 | 20.12.2023 | France |
| PQ762729 | 18.11.2023 | France |
| PQ762955 | 06.11.2023 | France |
| PQ762723 | 03.11.2023 | France |
| PQ763148 | 29.12.2023 | France |
| PQ762960 | 09.01.2024 | France |
| OQ024134 | 14.11.2022 | USA |
| OR522509 | 22.01.2023 | USA |
| OR795476 | 07.11.2022 | Germany |
| OR795474 | 03.11.2022 | Germany |
| OR143215 | 01.12.2022 | USA |
| PP681247 | 06.12.2022 | USA |
| PP681265 | 12.12.2022 | USA |
| OR522465 | 01.11.2022 | USA |
| PQ763035 | 11.03.2024 | France |
| PP270264 | 04.12.2023 | USA |
| OR143134 | 31.01.2023 | USA |
| PP957766 | 27.03.2024 | USA |
| PP847390 | 23.01.2024 | USA |
| PP957769 | 27.03.2024 | USA |
| PQ638679 | 10.12.2022 | USA |
| OP890312 | 31.10.2022 | USA |
| PQ638728 | 07.10.2022 | USA |
| PP970043 | 16.11.2023 | Ireland |
| PQ594188 | 01.01.2023 | China |
| PQ192486 | 17.09.2022 | Thailand |
| PP270259 | 05.12.2023 | USA |
| PQ117647 | 15.12.2023 | USA |
| PP970016 | 31.10.2023 | Ireland |
| PP969955 | 22.11.2023 | Ireland |
| PP969996 | 20.11.2023 | Ireland |
| OR840705 | 08.04.2023 | China |
| PP504649 | 13.11.2023 | USA |
| PQ066215 | 19.11.2023 | USA |
| PP270245 | 20.11.2023 | USA |
| PP781428 | 27.02.2024 | USA |
| LC816571 | 08.08.2023 | Japan |
| PQ788213 | 11.11.2024 | USA |
| PP974157 | 17.12.2023 | China |

| | | |
|---|---|---|
| LC816569 | 14.08.2023 | Japan |
| PQ762972 | 04.12.2023 | France |
| PP270268 | 05.12.2023 | USA |
| PQ788223 | 17.11.2024 | USA |
| PQ192490 | 11.08.2023 | Thailand |
| LC816570 | 31.08.2023 | Japan |
| PQ192489 | 10.10.2023 | Thailand |
| PQ390735 | 22.12.2023 | USA |
| PP504639 | 13.11.2023 | USA |
| PQ762998 | 12.01.2024 | France |
| PQ762863 | 30.12.2023 | France |
| PQ762957 | 26.12.2023 | France |
| PQ763027 | 23.12.2023 | France |
| PQ762621 | 22.11.2023 | France |
| PQ737344 | 22.11.2023 | France |
| OK500265 | 29.04.2021 | France |
| OR522467 | 06.11.2022 | USA |
| OR522459 | 25.10.2022 | USA |
| OR522522 | 11.12.2022 | USA |
| PQ638667 | 25.11.2022 | USA |
| PP495849 | 09.01.2024 | USA |
| PQ638698 | 08.11.2022 | USA |
| PQ638787 | 22.11.2023 | USA |
| PP495845 | 28.12.2023 | USA |
| PQ638798 | 02.11.2023 | USA |
| PP352348 | 05.10.2022 | USA |
| PP352366 | 01.11.2022 | USA |
| OR522528 | 31.01.2023 | USA |
| OR522505 | 27.12.2022 | USA |
| OR975299 | 27.12.2022 | USA |
| PP969966 | 11.10.2022 | Ireland |
| PP203264 | 31.12.2022 | USA |
| PQ638664 | 24.11.2022 | USA |
| PP681254 | 23.11.2022 | USA |
| OP890317 | 22.08.2022 | USA |
| OR143185 | 14.01.2023 | USA |
| OR143192 | 29.12.2022 | USA |
| OR522472 | 08.11.2022 | USA |
| OP890336 | 01.11.2022 | USA |

| | | |
|---|---|---|
| OR882976 | 07.11.2022 | USA |
| OR915768 | 22.11.2022 | USA |
| OP890325 | 01.10.2022 | USA |
| OR915753 | 15.12.2022 | USA |
| OR143146 | 08.12.2022 | USA |
| PP352370 | 01.11.2022 | USA |
| PP681276 | 06.01.2023 | USA |
| OR522469 | 06.11.2022 | USA |
| OR522495 | 04.11.2022 | USA |
| OQ024156 | 03.11.2022 | USA |
| OP890334 | 01.11.2022 | USA |
| OP890331 | 01.10.2022 | USA |
| OP890327 | 01.10.2022 | USA |
| OR522514 | 10.11.2022 | USA |
| OR522476 | 10.11.2022 | USA |
| PP795153 | 01.12.2023 | USA |
| OR522511 | 10.01.2023 | USA |
| OR872584 | 24.08.2022 | USA |
| OP890335 | 01.11.2022 | USA |
| OR522485 | 04.12.2022 | USA |
| OR143156 | 16.12.2022 | USA |
| OR915742 | 15.12.2022 | USA |
| OR143200 | 17.01.2023 | USA |
| OR162281 | 02.03.2023 | Kenya |
| PP934449 | 26.02.2024 | USA |
| PP882678 | 05.12.2023 | USA |
| PP270261 | 11.12.2023 | USA |
| PP970002 | 24.11.2023 | Ireland |
| PP594802 | 24.11.2023 | USA |
| PP969980 | 10.10.2022 | Ireland |
| PP270254 | 25.10.2023 | USA |
| PP957759 | 07.02.2024 | USA |
| PQ618080 | 10.07.2024 | Panama |
| OR795479 | 28.11.2022 | Germany |
| PQ762894 | 05.01.2024 | France |
| PQ763040 | 30.12.2023 | France |
| PQ763100 | 29.12.2023 | France |
| PQ762895 | 19.12.2023 | France |
| PQ762984 | 13.12.2023 | France |

| | | |
|---|---|---|
| PQ762804 | 12.12.2023 | France |
| PQ762893 | 08.12.2023 | France |
| PQ762888 | 06.12.2023 | France |
| PQ762900 | 04.12.2023 | France |
| PQ762937 | 29.11.2023 | France |
| PQ763166 | 28.11.2023 | France |
| PQ763022 | 26.11.2023 | France |
| PQ737348 | 19.11.2023 | France |
| PQ762633 | 29.10.2023 | France |
| PQ737355 | 28.10.2023 | France |
| PQ762632 | 28.10.2023 | France |
| PQ762907 | 27.10.2023 | France |
| PQ737352 | 20.10.2023 | France |
| PQ737350 | 19.10.2023 | France |
| PQ762627 | 19.10.2023 | France |
| PQ737351 | 19.10.2023 | France |
| PQ763065 | 30.01.2024 | France |
| PQ737356 | 29.10.2023 | France |
| PQ762802 | 09.12.2023 | France |
| PQ763142 | 07.11.2023 | France |
| PQ762889 | 15.11.2023 | France |
| PQ763081 | 20.11.2023 | France |
| PQ762880 | 19.12.2023 | France |
| PQ762629 | 20.10.2023 | France |
| PQ762628 | 19.10.2023 | France |
| PQ762885 | 04.12.2023 | France |
| PQ762625 | 19.11.2023 | France |
| PQ763169 | 30.12.2023 | France |
| PQ762878 | 10.12.2023 | France |
| PQ763125 | 15.11.2023 | France |
| PQ762891 | 26.12.2023 | France |
| PQ763103 | 07.01.2024 | France |
| PQ618079 | 11.08.2023 | Panama |
| OR522492 | 07.12.2022 | USA |
| OR522484 | 02.12.2022 | USA |
| OR872634 | 24.06.2022 | USA |
| OR872611 | 24.08.2022 | USA |
| OR915745 | 31.10.2022 | USA |
| OR872613 | 13.10.2022 | USA |

| | | |
|---|---|---|
| PP781396 | 08.01.2024 | USA |
| PP709391 | 03.01.2024 | USA |
| OR915730 | 31.10.2022 | USA |
| OR915751 | 02.10.2022 | USA |
| OR872603 | 26.10.2022 | USA |
| OR872595 | 15.07.2022 | USA |
| PP969970 | 25.10.2022 | Ireland |
| PP969977 | 11.10.2022 | Ireland |
| PP969945 | 06.10.2022 | Ireland |
| PP969946 | 06.10.2022 | Ireland |
| PP969944 | 06.10.2022 | Ireland |
| OR872593 | 16.10.2022 | USA |
| OR872642 | 25.10.2022 | USA |
| OR882993 | 27.11.2022 | USA |
| OR883015 | 27.11.2022 | USA |
| PQ638690 | 11.11.2022 | USA |
| OR872591 | 27.10.2022 | USA |
| OR872598 | 14.09.2022 | USA |
| OR872633 | 11.05.2022 | USA |
| OR882997 | 13.11.2022 | USA |
| OR883006 | 01.05.2023 | USA |
| PP903816 | 19.10.2022 | USA |
| OR915739 | 17.12.2022 | USA |
| OR915733 | 13.10.2022 | USA |
| OR872631 | 08.07.2022 | USA |
| OR872606 | 21.10.2022 | USA |
| PP781404 | 21.12.2023 | USA |
| PP781422 | 26.01.2024 | USA |
| PP882676 | 21.11.2023 | USA |
| PP681252 | 08.12.2022 | USA |
| PQ834846 | 04.11.2022 | Mexico |
| PQ834837 | 11.10.2022 | Mexico |
| OR915750 | 28.11.2022 | USA |
| OR143157 | 05.01.2023 | USA |
| PP903833 | 11.04.2024 | USA |
| PQ849803 | 10.08.2024 | USA |
| PQ638705 | 16.11.2022 | USA |
| PQ638694 | 11.11.2022 | USA |
| OQ171905 | 26.10.2022 | USA |

| | | |
|---|---|---|
| PP910782 | 13.02.2024 | USA |
| PP910781 | 13.02.2024 | USA |
| PP352359 | 01.11.2022 | USA |
| PP352335 | 01.10.2022 | USA |
| PP352339 | 30.11.2022 | USA |
| PP352387 | 01.12.2022 | USA |
| PQ834886 | 21.11.2023 | Mexico |
| PQ834885 | 21.11.2023 | Mexico |
| PQ834882 | 13.11.2023 | Mexico |
| PQ638700 | 11.11.2022 | USA |
| OP890324 | 31.10.2022 | USA |
| OR522482 | 06.12.2022 | USA |
| PP760398 | 01.11.2022 | USA |
| OP890320 | 01.11.2022 | USA |
| OP890328 | 01.11.2022 | USA |
| PP760404 | 01.12.2022 | USA |
| OR522463 | 31.10.2022 | USA |
| OR883016 | 11.11.2022 | USA |
| PQ638691 | 11.11.2022 | USA |
| PQ638665 | 24.11.2022 | USA |
| PQ638671 | 24.11.2022 | USA |
| PQ638660 | 10.10.2022 | USA |
| PP178651 | 04.12.2022 | USA |
| PQ638689 | 11.11.2022 | USA |
| PP760416 | 12.10.2022 | USA |
| OP890321 | 12.10.2022 | USA |
| OR915747 | 11.10.2022 | USA |
| OP890338 | 01.10.2022 | USA |
| OP890318 | 01.10.2022 | USA |
| PP970039 | 10.11.2023 | Ireland |
| PP997255 | 01.04.2024 | USA |
| PQ390721 | 19.12.2023 | USA |
| PQ638729 | 05.09.2023 | USA |
| PP530257 | 18.12.2023 | USA |
| PP530271 | 19.12.2023 | USA |
| PP530275 | 07.12.2023 | USA |
| PQ638786 | 23.11.2023 | USA |
| PP495847 | 04.01.2024 | USA |
| OQ024127 | 03.11.2022 | USA |

| | | |
|---|---|---|
| OR143167 | 17.12.2022 | USA |
| OQ171902 | 18.10.2022 | USA |
| OR882996 | 17.04.2023 | USA |
| OR522508 | 23.01.2023 | USA |
| OR522513 | 15.01.2023 | USA |
| PP352380 | 31.12.2022 | USA |
| OR143151 | 10.12.2022 | USA |
| PP352373 | 31.12.2022 | USA |
| PQ638663 | 24.11.2022 | USA |
| OR872601 | 26.10.2022 | USA |
| OR143205 | 05.12.2022 | USA |
| OQ024113 | 14.11.2022 | USA |
| OQ171908 | 21.10.2022 | USA |
| OQ024129 | 04.11.2022 | USA |
| OQ024144 | 03.11.2022 | USA |
| OP890333 | 01.11.2022 | USA |
| OP965712 | 01.11.2022 | USA |
| OR143150 | 21.12.2022 | USA |
| OR143175 | 25.12.2022 | USA |
| OR143172 | 05.12.2022 | USA |
| PP760388 | 31.10.2022 | USA |
| PP795155 | 11.12.2023 | USA |
| PP270236 | 21.11.2023 | USA |
| OR601478 | 03.11.2022 | USA |
| PQ638711 | 16.10.2022 | USA |
| OR522475 | 09.11.2022 | USA |
| PP903834 | 20.10.2022 | USA |
| OR915749 | 12.12.2022 | USA |
| OR872582 | 25.10.2022 | USA |
| OR883022 | 28.06.2023 | USA |
| OR882980 | 14.11.2022 | USA |
| PP903807 | 24.10.2022 | USA |
| PP903806 | 30.11.2023 | USA |
| PQ066225 | 01.12.2023 | USA |
| PP903828 | 29.10.2022 | USA |
| PP903822 | 06.10.2022 | USA |
| OR882970 | 12.11.2022 | USA |
| OR882995 | 28.02.2023 | USA |
| OR882971 | 24.03.2023 | USA |

| | | |
|---|---|---|
| PP781399 | 17.12.2023 | USA |
| OR883004 | 16.12.2022 | USA |
| OR883008 | 14.12.2022 | USA |
| PP903826 | 22.09.2022 | USA |
| PQ066218 | 12.09.2022 | USA |
| PP903829 | 17.09.2022 | USA |
| PP903835 | 19.10.2022 | USA |
| OR882973 | 07.11.2022 | USA |
| OR915775 | 01.12.2022 | USA |
| PP903825 | 22.11.2022 | USA |
| OR522519 | 17.01.2023 | USA |
| OR143147 | 09.12.2022 | USA |
| OR143155 | 11.12.2022 | USA |
| PP681278 | 10.01.2023 | USA |
| OR522480 | 16.10.2022 | USA |
| OR522483 | 05.12.2022 | USA |
| OR522488 | 29.11.2022 | USA |
| OR522489 | 29.11.2022 | USA |
| OR143199 | 11.03.2023 | USA |
| OR143173 | 17.12.2022 | USA |
| OR143208 | 06.12.2022 | USA |
| OR143219 | 01.12.2022 | USA |
| OR143193 | 31.12.2022 | USA |
| PP760392 | 01.10.2022 | USA |
| OR143203 | 06.12.2022 | USA |
| OR882979 | 23.11.2022 | USA |
| OR883013 | 03.11.2022 | USA |
| OR883001 | 28.10.2022 | USA |
| PP903808 | 03.11.2022 | USA |
| PP903820 | 26.10.2022 | USA |
| PP903831 | 26.10.2022 | USA |
| PP903832 | 26.10.2022 | USA |
| OR882990 | 07.12.2022 | USA |
| OR882987 | 27.11.2022 | USA |
| OR882998 | 25.11.2022 | USA |
| OR883021 | 22.11.2022 | USA |
| PP903818 | 11.10.2022 | USA |
| PP781423 | 15.02.2024 | USA |
| PQ066223 | 18.12.2023 | USA |

| | | |
|---|---|---|
| PQ066226 | 18.12.2023 | USA |
| PP903823 | 19.10.2022 | USA |
| OR882984 | 27.11.2022 | USA |
| OR915764 | 22.11.2022 | USA |
| OR882994 | 04.11.2022 | USA |
| PP781412 | 02.01.2024 | USA |
| OR883011 | 13.12.2022 | USA |
| PP903811 | 09.10.2022 | USA |
| OR882977 | 20.11.2022 | USA |
| OR915766 | 27.11.2022 | USA |
| PP903810 | 20.03.2024 | USA |
| PP781394 | 03.01.2024 | USA |
| OR915759 | 10.11.2022 | USA |
| OR915746 | 02.11.2022 | USA |
| OR915757 | 20.10.2022 | USA |
| OR882983 | 31.10.2022 | USA |
| OR883019 | 31.10.2022 | USA |
| OR915748 | 27.10.2022 | USA |
| OR915736 | 01.01.2023 | USA |
| OR915743 | 01.01.2023 | USA |
| OR915761 | 01.01.2023 | USA |
| PQ762506 | 19.03.2024 | USA |
| PQ762500 | 14.03.2024 | USA |
| PP978497 | 19.02.2024 | USA |
| PP957757 | 07.02.2024 | USA |
| PP748753 | 28.01.2024 | USA |
| PP910770 | 27.01.2024 | USA |
| PP847392 | 17.01.2024 | USA |
| PP847375 | 06.01.2024 | USA |
| PP709458 | 02.01.2024 | USA |
| PP709389 | 31.12.2023 | USA |
| PP709381 | 17.12.2023 | USA |
| PP830396 | 14.12.2023 | USA |
| PP594803 | 26.11.2023 | USA |
| PP504640 | 11.11.2023 | USA |
| PP386336 | 26.10.2023 | USA |
| PP237782 | 02.10.2023 | USA |
| PP830404 | 21.12.2023 | USA |
| PQ762525 | 17.05.2024 | USA |

| | | |
|---|---|---|
| PP910765 | 25.01.2024 | USA |
| PP978506 | 20.02.2024 | USA |
| PP594800 | 24.11.2023 | USA |
| PP830407 | 21.12.2023 | USA |
| PQ762516 | 01.05.2024 | USA |
| PP957770 | 28.03.2024 | USA |
| PP910769 | 26.01.2024 | USA |
| PP847385 | 21.01.2024 | USA |
| PP401818 | 29.11.2023 | USA |
| PP401810 | 05.11.2023 | USA |
| PP504646 | 09.11.2023 | USA |
| PP504638 | 05.11.2023 | USA |
| PP978532 | 11.03.2024 | USA |
| PP830411 | 25.12.2023 | USA |
| PP594808 | 25.11.2023 | USA |
| PP847372 | 04.01.2024 | USA |
| PP957758 | 05.02.2024 | USA |
| PP978524 | 29.02.2024 | USA |
| PQ762489 | 30.01.2024 | USA |
| PP934466 | 24.03.2024 | USA |
| PP978500 | 19.02.2024 | USA |
| PP847374 | 06.01.2024 | USA |
| PP709455 | 04.01.2024 | USA |
| PP709382 | 23.12.2023 | USA |
| PP504642 | 09.11.2023 | USA |
| PP957751 | 03.02.2024 | USA |
| PP910763 | 25.01.2024 | USA |
| PP709390 | 02.01.2024 | USA |
| PP709383 | 25.12.2023 | USA |
| PP957762 | 09.02.2024 | USA |
| PP910777 | 31.01.2024 | USA |
| PP978503 | 20.02.2024 | USA |
| PP978502 | 16.02.2024 | USA |
| PP973761 | 05.02.2024 | USA |
| PQ762519 | 30.04.2024 | USA |
| PQ762521 | 08.05.2024 | USA |
| PQ762518 | 30.04.2024 | USA |
| PQ762481 | 12.01.2024 | USA |
| PP401812 | 07.11.2023 | USA |

| | | |
|---|---|---|
| PP978518 | 24.02.2024 | USA |
| PP668175 | 18.12.2023 | USA |
| PP830383 | 09.12.2023 | USA |
| PP910784 | 11.02.2024 | USA |
| PQ762493 | 13.03.2024 | USA |
| PQ762485 | 14.01.2024 | USA |
| PP594801 | 24.11.2023 | USA |
| PP709377 | 27.12.2023 | USA |
| PP709443 | 26.12.2023 | USA |
| PQ788246 | 13.01.2024 | USA |
| PP709388 | 28.12.2023 | USA |
| PP830412 | 24.12.2023 | USA |
| PP978533 | 11.03.2024 | USA |
| PP709441 | 23.12.2023 | USA |
| PP847373 | 10.01.2024 | USA |
| PP957748 | 03.02.2024 | USA |
| PP795154 | 07.12.2023 | USA |
| PP270267 | 11.12.2023 | USA |
| OR883009 | 19.06.2023 | USA |
| PP795138 | 22.10.2023 | USA |
| PP781401 | 28.12.2023 | USA |
| PP957773 | 08.04.2024 | USA |
| PP978512 | 22.02.2024 | USA |
| PP957745 | 02.02.2024 | USA |
| PP973751 | 15.01.2024 | USA |
| PP709444 | 25.12.2023 | USA |
| PP668174 | 19.12.2023 | USA |
| PP504632 | 04.11.2023 | USA |
| PP386347 | 27.10.2023 | USA |
| PP237793 | 15.10.2023 | USA |
| PP934450 | 29.02.2024 | USA |
| PP934465 | 22.03.2024 | USA |
| PP847367 | 09.01.2024 | USA |
| PP401820 | 03.12.2023 | USA |
| PP847394 | 21.01.2024 | USA |
| PQ762510 | 19.03.2024 | USA |
| PQ788248 | 12.01.2024 | USA |
| PP934459 | 09.03.2024 | USA |
| PP910778 | 13.02.2024 | USA |

| | | |
|---|---|---|
| PP910779 | 13.02.2024 | USA |
| PP910775 | 28.01.2024 | USA |
| PP709442 | 22.12.2023 | USA |
| PP709449 | 03.01.2024 | USA |
| PP978505 | 17.02.2024 | USA |
| PP594811 | 25.11.2023 | USA |
| PP973759 | 29.01.2024 | USA |
| PP830397 | 14.12.2023 | USA |
| PP847364 | 09.01.2024 | USA |
| PP978520 | 25.02.2024 | USA |
| PP978519 | 24.02.2024 | USA |
| PP668169 | 05.12.2023 | USA |
| PQ762502 | 17.03.2024 | USA |
| PQ762496 | 14.03.2024 | USA |
| PQ788253 | 14.01.2024 | USA |
| PP401814 | 20.11.2023 | USA |
| PP504634 | 06.11.2023 | USA |
| PP594817 | 04.12.2023 | USA |
| PP830392 | 14.12.2023 | USA |
| OR143211 | 08.01.2023 | USA |
| PP270233 | 20.11.2023 | USA |
| PP270252 | 20.11.2023 | USA |
| PP270248 | 01.11.2023 | USA |
| PP270263 | 08.12.2023 | USA |
| PP781392 | 29.12.2023 | USA |
| PQ114111 | 29.12.2022 | USA |
| PQ638683 | 27.12.2022 | USA |
| PP781379 | 17.12.2023 | USA |
| PP781421 | 17.01.2024 | USA |
| OR872612 | 20.09.2022 | USA |
| OR872597 | 16.10.2022 | USA |
| PP795135 | 11.10.2023 | USA |
| OQ024126 | 04.11.2022 | USA |
| PQ638661 | 22.11.2022 | USA |
| PP352365 | 01.11.2022 | USA |
| OQ024145 | 14.11.2022 | USA |
| PP352331 | 01.10.2022 | USA |
| OR522466 | 05.11.2022 | USA |
| OR143148 | 10.12.2022 | USA |

| | | |
|---|---|---|
| OR143145 | 08.12.2022 | USA |
| OR143158 | 25.01.2023 | USA |
| OR143169 | 03.12.2022 | USA |
| OR143143 | 26.12.2022 | USA |
| OR143163 | 04.02.2023 | USA |
| PP270244 | 08.11.2023 | USA |
| OR915767 | 16.09.2023 | USA |
| PP781391 | 16.02.2024 | USA |
| OR143160 | 25.01.2023 | USA |
| OR143178 | 18.04.2023 | USA |
| OR143196 | 08.12.2022 | USA |
| PP910773 | 28.01.2024 | USA |
| OR915776 | 17.07.2023 | USA |
| PQ762513 | 20.04.2024 | USA |
| PP342427 | 06.12.2023 | USA |
| PP342428 | 06.12.2023 | USA |
| PP342418 | 29.11.2023 | USA |
| OR915754 | 02.11.2022 | USA |
| OR143165 | 23.02.2023 | USA |
| OR143164 | 22.02.2023 | USA |
| OR143159 | 09.02.2023 | USA |
| OR522525 | 29.12.2022 | USA |
| OR143141 | 18.12.2022 | USA |
| OR883020 | 13.04.2023 | USA |
| OR143201 | 05.12.2022 | USA |
| OR143180 | 30.11.2022 | USA |
| OR143186 | 06.12.2022 | USA |
| OR143176 | 16.12.2022 | USA |
| OR143216 | 13.12.2022 | USA |
| PP270260 | 15.11.2023 | USA |
| PP978516 | 22.02.2024 | USA |
| PP978504 | 17.02.2024 | USA |
| PP709451 | 01.01.2024 | USA |
| PP830401 | 25.12.2023 | USA |
| PP830414 | 22.12.2023 | USA |
| OR872614 | 04.11.2022 | USA |
| OR872629 | 06.11.2022 | USA |
| OR522500 | 20.12.2022 | USA |
| PP681263 | 12.12.2022 | USA |

| | | |
|---|---|---|
| PP760401 | 01.11.2022 | USA |
| PP681248 | 06.12.2022 | USA |
| PP760396 | 30.11.2022 | USA |
| PQ638708 | 16.11.2022 | USA |
| OR522471 | 07.11.2022 | USA |
| PP760408 | 01.01.2023 | USA |
| OQ024110 | 07.11.2022 | USA |
| OQ024140 | 03.11.2022 | USA |
| OQ171925 | 12.10.2022 | USA |
| OQ171929 | 12.10.2022 | USA |
| OQ024146 | 04.11.2022 | USA |
| OQ024151 | 04.11.2022 | USA |
| PP296424 | 02.12.2022 | USA |
| PP278016 | 30.11.2022 | USA |
| PP352372 | 01.11.2022 | USA |
| PP352343 | 01.11.2022 | USA |
| OR915771 | 27.11.2022 | USA |
| OR143153 | 09.12.2022 | USA |
| PQ788242 | 27.10.2024 | USA |
| PP973756 | 01.02.2024 | USA |
| PP973767 | 06.02.2024 | USA |
| PP781395 | 18.01.2024 | USA |
| PP781400 | 11.01.2024 | USA |
| PQ762486 | 15.01.2024 | USA |
| PQ788240 | 31.10.2024 | USA |
| PP270235 | 02.11.2023 | USA |
| PQ834894 | 04.12.2023 | Mexico |
| PQ834891 | 27.11.2023 | Mexico |
| PQ834898 | 05.12.2023 | Mexico |
| PQ834884 | 15.11.2023 | Mexico |
| PQ834892 | 28.11.2023 | Mexico |
| OR872592 | 27.10.2022 | USA |
| OP890313 | 24.10.2022 | USA |
| OP890326 | 24.10.2022 | USA |
| PP781427 | 15.01.2024 | USA |
| PP830384 | 10.12.2023 | USA |
| PQ762754 | 27.11.2023 | France |
| PP910753 | 11.01.2024 | USA |
| PP973768 | 05.02.2024 | USA |

| | | |
|---|---|---|
| PP973762 | 03.02.2024 | USA |
| PQ762499 | 14.03.2024 | USA |
| PQ008874 | 05.03.2024 | USA |
| PP910755 | 12.01.2024 | USA |
| PP847368 | 09.01.2024 | USA |
| PP709372 | 25.12.2023 | USA |
| PP504643 | 10.11.2023 | USA |
| PP903819 | 08.01.2024 | USA |
| PP903824 | 08.01.2024 | USA |
| PP934455 | 28.02.2024 | USA |
| PQ788203 | 26.11.2024 | USA |
| PQ788225 | 17.11.2024 | USA |
| PQ849790 | 01.12.2024 | USA |
| PQ849783 | 09.12.2024 | USA |
| PQ849784 | 08.12.2024 | USA |
| PQ849772 | 07.12.2024 | USA |
| OQ171913 | 28.10.2022 | USA |
| OR915729 | 07.01.2023 | USA |
| OQ171912 | 12.07.2022 | USA |
| PQ762743 | 10.10.2023 | France |
| OR143135 | 11.02.2023 | USA |
| PP203265 | 10.10.2022 | USA |
| PP830379 | 06.12.2023 | USA |
| PP795139 | 26.10.2023 | USA |
| PP795141 | 26.10.2023 | USA |
| PP795146 | 06.11.2023 | USA |
| PP795144 | 30.10.2023 | USA |
| PP795132 | 05.10.2023 | USA |
| OR882989 | 04.11.2022 | USA |
| PP352374 | 30.11.2022 | USA |
| OR143152 | 14.12.2022 | USA |
| PP760412 | 31.12.2022 | USA |
| OR143174 | 30.11.2022 | USA |
| OR143136 | 20.12.2022 | USA |
| OR143217 | 11.12.2022 | USA |
| OR795473 | 20.10.2022 | Germany |
| OR143191 | 04.12.2022 | USA |
| OR143161 | 24.01.2023 | USA |
| OR143181 | 02.12.2022 | USA |

| | | |
|---|---|---|
| OR522532 | 04.01.2023 | USA |
| PP681256 | 08.12.2022 | USA |
| PP760387 | 26.11.2022 | USA |
| OP890323 | 26.11.2022 | USA |
| OR915762 | 26.12.2022 | USA |
| OR915731 | 11.12.2022 | USA |
| OR915755 | 24.10.2022 | USA |
| OR915763 | 11.12.2022 | USA |
| OP890329 | 01.11.2022 | USA |
| PQ638669 | 23.11.2022 | USA |
| PP178652 | 25.11.2022 | USA |
| OP890339 | 01.10.2022 | USA |
| PP760399 | 01.11.2022 | USA |
| PP781411 | 09.01.2024 | USA |
| OR143214 | 04.01.2023 | USA |
| OR883012 | 17.12.2022 | USA |
| PQ834851 | 23.11.2022 | Mexico |
| OR872643 | 18.09.2022 | USA |
| OQ171923 | 21.10.2022 | USA |
| PQ834850 | 24.10.2022 | Mexico |
| OQ171930 | 18.10.2022 | USA |
| OQ171903 | 17.10.2022 | USA |
| OQ171898 | 31.10.2022 | USA |
| PP352332 | 01.10.2022 | USA |
| OQ024118 | 04.11.2022 | USA |
| OQ024132 | 02.11.2022 | USA |
| PP352360 | 30.11.2022 | USA |
| PP352328 | 31.10.2022 | USA |
| OQ024133 | 03.11.2022 | USA |
| PQ834873 | 18.11.2022 | Mexico |
| PP795152 | 27.11.2023 | USA |
| PQ834889 | 24.11.2023 | Mexico |
| PP594809 | 24.11.2023 | USA |
| PP847383 | 11.01.2024 | USA |
| PP709456 | 03.01.2024 | USA |
| PP934464 | 18.03.2024 | USA |
| PP681279 | 02.12.2022 | USA |
| OP890332 | 17.10.2022 | USA |
| OR143162 | 29.01.2023 | USA |

| | | |
|---|---|---|
| OR143182 | 16.01.2023 | USA |
| PP882670 | 19.12.2023 | USA |
| PP795163 | 24.01.2024 | USA |
| PQ834903 | 13.12.2023 | Mexico |
| PQ762504 | 17.03.2024 | USA |
| PQ762501 | 16.03.2024 | USA |
| PP973776 | 14.02.2024 | USA |
| OR915741 | 02.10.2022 | USA |
| PP795164 | 09.02.2024 | USA |
| OR915752 | 16.10.2022 | USA |
| OQ024130 | 03.11.2022 | USA |
| OQ171928 | 25.10.2022 | USA |
| PQ834877 | 06.12.2022 | Mexico |
| PP270258 | 06.12.2023 | USA |
| PP770453 | 27.02.2024 | USA |
| PP770470 | 29.01.2024 | USA |
| PP781424 | 31.01.2024 | USA |
| PQ618078 | 28.07.2023 | Panama |
| PQ834896 | 01.12.2023 | Mexico |
| PP270246 | 06.11.2023 | USA |
| PP504650 | 11.11.2023 | USA |
| PP830394 | 14.12.2023 | USA |
| PQ762985 | 10.12.2023 | France |
| PQ762715 | 04.01.2024 | France |
| PQ763023 | 24.12.2023 | France |
